# Comparative Evaluation of Behavioral-Epidemic Models Using COVID-19 Data

**DOI:** 10.1101/2024.11.08.24316998

**Authors:** Nicolò Gozzi, Nicola Perra, Alessandro Vespignani

## Abstract

Characterizing the feedback linking human behavior and the transmission of infectious diseases (i.e., behavioral changes) remains a significant challenge in computational and mathematical Epidemiology. Existing Behavioral Feedback Models often lack real-world data calibration and cross-model performance evaluation in both retrospective analysis and forecasting. In this study, we systematically compare the performance of three mechanistic behavioral models across nine geographies and two modeling tasks during the first wave of COVID-19, using various metrics. The first model, a Data-Driven Behavioral Feedback Model, incorporates behavioral changes by leveraging mobility data to capture variations in contact patterns. The second and third models are Analytical Behavioral Feedback Models, which simulate the feedback loop either through the explicit representation of different behavioral compartments within the population or by utilizing an effective non-linear force of infection. Our results do not identify a single best model overall, as performance varies based on factors such as data availability, data quality, and the choice of performance metrics. While the data-driven model incorporates substantial real-time behavioral information, the Analytical Compartmental Behavioral Feedback Model often demonstrates superior or equivalent performance in both retrospective fitting and out-of-sample forecasts. Overall, our work offers guidance for future approaches and methodologies to better integrate behavioral changes into the modeling and projection of epidemic dynamics.

## 1 Introduction

During the COVID-19 Pandemic, epidemic models became central tools for providing situational awareness, scenario analysis, and forecasts. Specifically, mathematical and computational models were used to characterize the initial outbreak phases [1–6], assess policy interventions [6–12], evaluate risks from new virus strains [13–17], and estimate outcomes of various vaccination strategies [18–23]. Achieving the required level of realism in these models necessitated incorporating population-level behavioral changes due to epidemic awareness and mandated or recommended non-pharmaceutical interventions (NPIs) [24–26]. However, capturing the feedback loop between the transmission of infectious diseases and human behavior has long been regarded, and still remains, as a major challenge in Epidemiology [24, 27–29].

In this context, we identify two major classes of mechanistic modeling approaches: Data-Driven Behavioral Models and Analytical Behavioral Feedback Models. Data-Driven Behavioral Models integrate real-world data on behavioral changes, such as mobility patterns and social distancing measures into epidemic simulations [13, 20, 30–44]. Hence, these models rely on empirical data to simulate how behaviors change. Analytical Behavioral Feedback Models, instead, use theoretical frameworks to incorporate non-linear mechanisms describing how individual behaviors change in response to the epidemic’s progression [45–52]. These models do not rely on real-world data but rather on mechanistic rules that capture the feedback loop between behavior and epidemic dynamics.

Data-driven approaches have been prevalent in the COVID-19 literature for several reasons. First, using empirical data can drastically reduce the number of free parameters and explicit mechanisms needed to capture human behavior. Additionally, most models in the analytic class were developed before the COVID-19 pandemic and often lacked empirical validation [27]. Using an explicit behavioral model rather than data, however, has the potential to accurately capture the interplay between human behavior and the spread of infectious diseases, enabling more precise projections and forecasts. Furthermore, data-driven models are not necessarily simpler than their analytical counterparts. In fact, they often integrate large amounts of temporal data, relying on methodologies that involve assumptions, may be prone to biases and other data collection issues. These considerations stress the need for a systematic analysis of the performance and calibration of these model classes to understand their reliability and usefulness in informing decision-making processes.

Here, we present a systematic comparison of the performance of different behavioral feedback models during the first wave of the COVID-19 pandemic, spanning nine geographies and two modeling tasks. Specifically, we investigate three mechanistic models: i) the *Data-Driven Behavioral Model* exemplifies data-driven approaches, leveraging mobility data to estimate effective changes in contact patterns; ii) the *Compartmental Behavioral Feedback Model* simulates the feedback loop by explicitly representing different behavioral classes within the population; iii) the *Effective Force of Infection Behavioral Feedback Model* employs an effective non-linear forcing to adjust the infection rate based on the epidemic’s progression and the resulting behavioral changes. We first quantitatively assess the performance of these models through a retrospective analysis of their ability to capture the dynamics of the first COVID-19 wave across nine diverse geographical areas. Using the same datasets, we also evaluate their out-of-sample forecasting performance over a rolling four-week horizon in these regions. Remarkably, in the retrospective analysis the Data-Driven Behavioral Model, which integrates mobility data, does not consistently outperform the Analytical Behavioral Feedback Models. In forecasting performance, similar results are observed, indicating that Analytical Behavioral Feedback Models, though largely neglected during the COVID-19 pandemic, can often provide superior, or comparable, performance by capturing the interplay between human behavior changes and disease progression.

Overall our results pave the way for a broader use of analytical behavioral approaches in epidemic modeling and forecasting contexts. Modeling choices should therefore consider factors such as data availability and quality, target metrics, and geographic scope. It is important to note that, despite similar performances in retrospective and forecasting analyses, the models often offer different characterizations of disease dynamics, exemplified by different effective reproductive numbers time series. This evidence reinforces the indication of interpreting disease dynamics in the context of the model’s structure rather than as intrinsic properties of the pathogen. While our results focus on COVID-19, they hold broad relevance for the analysis and forecasting of respiratory and other transmissible diseases.

## 2 Results

We consider three mechanistic models in which the disease progression is described via an age-structured Susceptible-Exposed-Infected-Recovered (SEIR) disease dynamic with the addition of compartments accounting for COVID-19 deaths and their delayed reporting (see Fig. 1). Each model differs in the way population behavioral changes are integrated into the dynamic:

**Figure 1.**
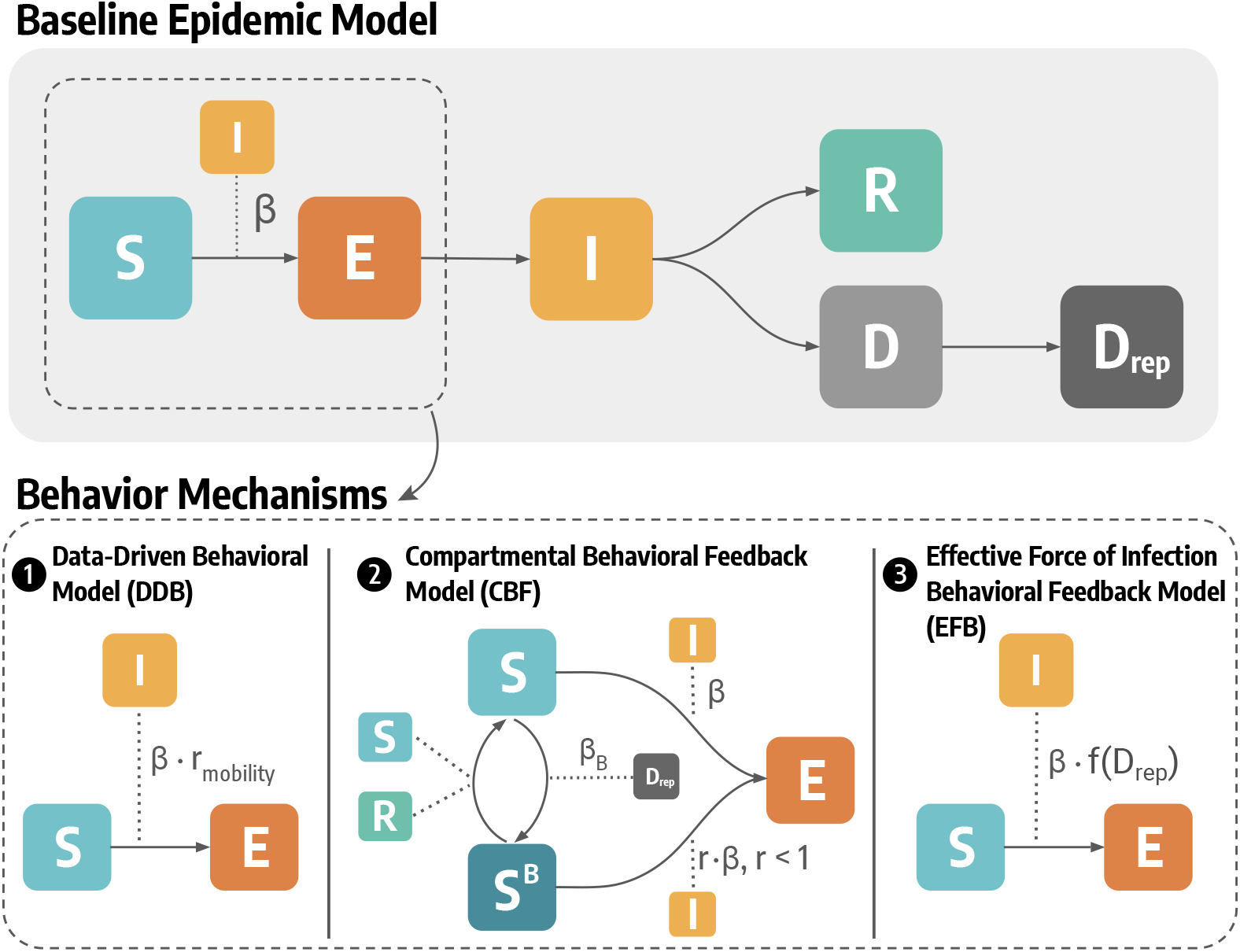
Flow diagrams of the epidemic-behavior models considered. The top row shows the baseline SEIR-like epidemic model. The bottom row shows the epidemic-behavior mechanisms for the three models considered. In particular, *β* indicates the transmission rate, *r*_*mobility*_ is the contact reduction parameter estimated on mobility data, *β*_*B*_ regulates the behavioral transitions in the Compartmental Behavioral Feedback model and *r* is the relative reduction in the force of infection of risk-averse individuals, and *f* (*D*_*rep*_) is a non-linear function of the number of reported deaths that modulates the force of infection in the Effective Force of Infection Behavioral Feedback model.

- *Data-Driven Behavioral* (DDB) model. This model integrates data from the COVID-19 Community Mobility Report published by Google LLC [53] to derive a time-varying contact reduction coefficient.
- *Compartmental Behavioral Feedback* (CBF) model. This model introduces behavioral changes explicitly through a new compartment *S*^*B*^ for Susceptible individuals who are risk averse. These individuals experience a relative reduction *r <* 1 in the force of infection. Additional parameters of the model characterize the transitions to and out of the risk-averse compartment.
- *Effective Force of Infection Behavioral Feedback* (EFB) model. This model integrates the behavior changes in the population with an explicit modulation of the transmissibility [51]. More precisely, we consider a non-linear function (that saturates as the number of reported deaths grows) characterizing the rate at which susceptible individuals acquire infection (i.e., the force of infection).

In Fig. 1 we show a schematic depiction of the compartmental structure and the transitions among compartments of each model. Full details of the models are provided in the Material and Methods section and the Supplementary Information. It is important to note that none of the models distinguishes between spontaneous and mandated behavioral changes [24, 27–29]. In all cases, increased risk aversion among individuals and the resulting reduction in contacts account for all causes leading to behavioral changes.

### Retrospective model inference

We calibrated the three models to fit the initial wave of COVID-19 deaths across nine distinct geographical areas: metropolitan areas such as Bogotá, Chicago, Jakarta, London, Madrid, New York, Rio de Janeiro, and Santiago de Chile, as well as a larger administrative region, such as Gauteng in South Africa. This selection captures a diverse range of epidemiological, socio-demographic, and socioeconomic contexts from both the global North and South. In each region, models are calibrated to weekly deaths using an Approximate Bayesian Computation - sequential Monte Carlo (ABC-SMC) algorithm [54] (details are reported in the Material and Methods section and the Supplementary Information). In Fig. 2, we present the fitted curves (median and 90% predictive intervals) for these nine locations using the three models. Overall, all models successfully replicate the shape of the observed epidemic curves. However, we observe inferior fit quality in specific cases. For instance, the models exhibit lower performance in the epidemic tail for Gauteng and Rio de Janeiro, possibly due to factors such as the completeness and reporting time of epidemiological data in those settings. To quantitatively characterize the model’s performance, in Tab. 1, we report the normalized mean absolute error (nMAE), the normalized weighted interval score (nWIS), and the Bayesian Information Criterion (BIC) weight of each model.

**Figure 2.**
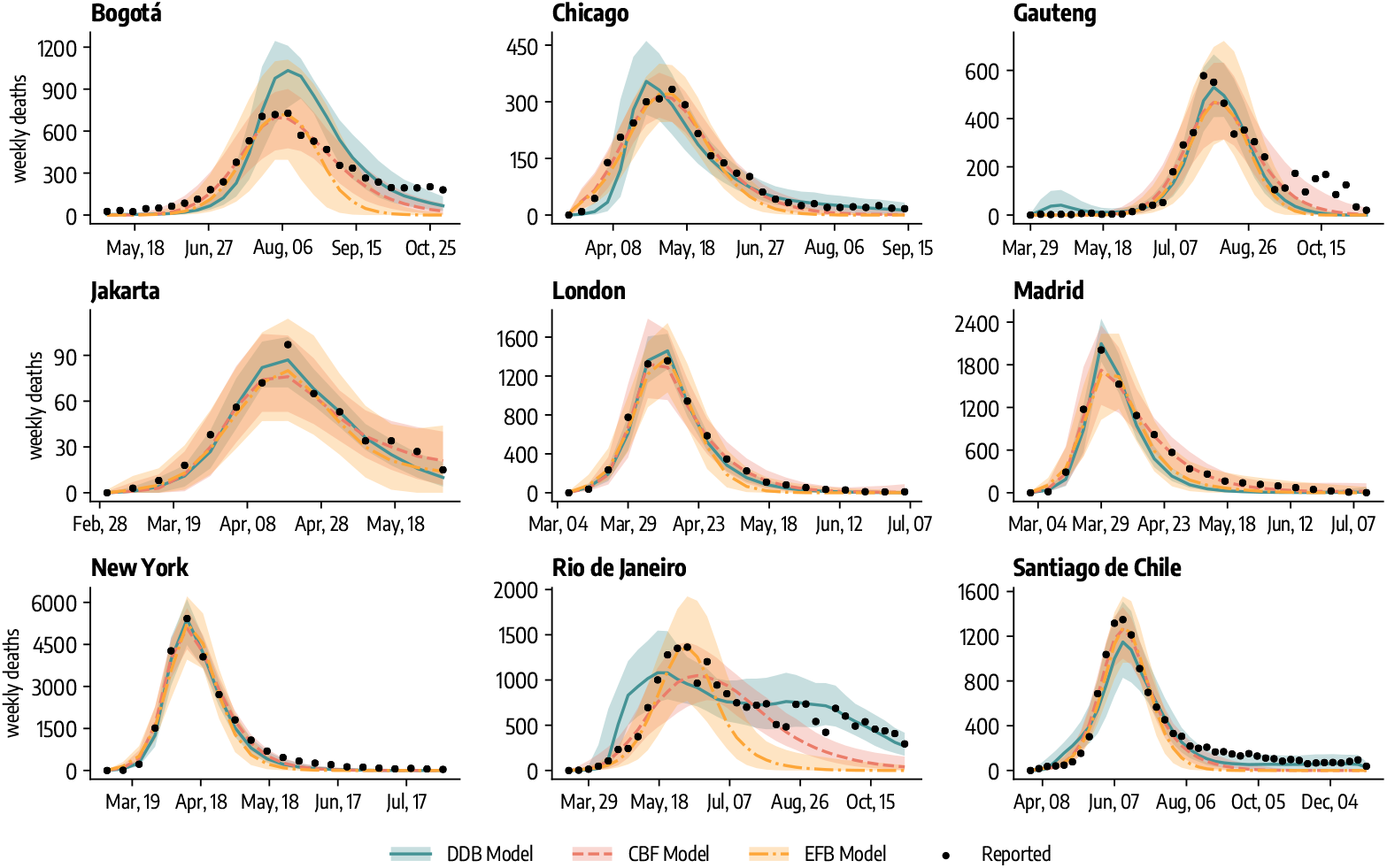
Fitted curves (median, 90% predictive intervals obtained from 1, 000 stochastic trajectories) of weekly deaths during the COVID-19 initial wave across the nine geographies considered and three epidemic-behavior models. DDB stands for Data-Driven Behavioral model, CBF for Compartmental Behavioral Feedback model, and EFB for Effective Force of Infection Behavioral Feedback model.

**Table 1:** Comparison of models performance in retrospective modeling task (** indicates probabilities *<* 0.01%). DDB stands for Data-Driven Behavioral model, CBF for Compartmental Behavioral Feedback model, and EFB for Effective Force of Infection Behavioral Feedback model.

|  | nMAE |  |  | nWIS |  |  | BIC Weights |  |  |
| --- | --- | --- | --- | --- | --- | --- | --- | --- | --- |
|  | DDB | CBF | EFB | DDB | CBF | EFB | DDB | CBF | EFB |
| *Bogotá* | 0.41 | **0.18** | 0.35 | 0.29 | **0.14** | 0.27 | ** | **99.99** | ** |
| *Chicago* | 0.20 | **0.14** | 0.20 | 0.12 | **0.09** | 0.14 | ** | **99.90** | 0.09 |
| *Gauteng* | 0.32 | **0.25** | 0.34 | 0.25 | **0.18** | 0.26 | **94.20** | 5.73 | 0.07 |
| *Jakarta* | 0.14 | **0.13** | 0.14 | **0.09** | **0.09** | 0.11 | **97.92** | 0.35 | 1.73 |
| *London* | 0.12 | **0.07** | 0.18 | 0.07 | **0.06** | 0.11 | 4.57 | **95.42** | ** |
| *Madrid* | 0.28 | **0.10** | 0.24 | 0.21 | **0.08** | 0.14 | ** | **99.99** | ** |
| *New York* | **0.13** | **0.13** | 0.21 | 0.09 | **0.08** | 0.15 | **98.97** | 1.02 | ** |
| *Rio de Janeiro* | **0.23** | 0.33 | 0.52 | **0.16** | 0.25 | 0.43 | **99.99** | ** | ** |
| *Santiago de Chile* | **0.24** | 0.25 | 0.32 | **0.14** | 0.19 | 0.26 | **63.80** | 36.17 | 0.03 |

When considering the nMAE of the median, the Compartmental Behavioral Feedback model is the top performer in all geographies, except for Rio de Janeiro and Santiago de Chile, where the Data-Driven Behavioral model performs better. The WIS measures the effectiveness of predictive intervals in bounding reported data, with normalization enabling comparisons across different geographies. Analyzing the nWIS, the Compartmental Behavioral Feedback model is the top performing model in 6 geographies (Bogotá, Chicago, Gauteng, London, Madrid, New York), although there is more variability in performance. The Data-Driven Behavioral model is the top performer in 3 geographies (Jakarta, Rio de Janeiro, and Santiago de Chile).

It is important to note that the models have different structures and number of free parameters that obfuscate the simple comparison through goodness of fit. To have a more unbiased estimator we consider the Bayesian Information Criterion that discounts the number of estimable parameters, and calculate the BIC weights of each model in each location. The BIC weight can be interpreted as the probability that any given model is the *best* model (i.e., likelihood of the model given the data) among those considered.

According to the BIC weights, the Compartmental Behavioral Feedback model is the most probable in 4 cases (Bogotá, Chicago, London, Madrid), while the Data-Driven Behavioral model is most probably the best model in the remaining 5 (Gauteng, Jakarta, New York, Rio de Janeiro, Santiago de Chile). We note how, in Data-Driven Behavioral models, the contact reduction values estimated on mobility data (i.e., *r*_*mobility*_(*t*)) are discounted from the number of free parameters as they are not subject to calibration. In the Supplementary Information, we also report the models’ accuracy in reproducing both the intensity and timing of epidemic peaks. Interestingly, we find that the Effective Force of Infection model and the Data-Driven model are more accurate than the Behavioral Compartmental model when evaluating these target quantities. This highlights that the definition of the best model often depends on the specific metric being evaluated.

### Estimates of transmission potential

Although the three models offer a similar fit of the epidemic trajectories, it is important to quantify the difference in the disease dynamic emerging from them. First, we considered the posterior distribution of the basic reproductive number (i.e., *R*_0_) provided by the three models across the nine geographies. This quantity is defined as the number of secondary infections, due to a single infectious individual, in an otherwise susceptible population [55]. In all three models, *R*_0_ is defined as 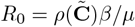, where 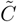 is the contact matrix weighted by age group populations, *ρ*(•) is the spectral radius, *β* is the transmission rate, and *μ* is the inverse of the infectious period. More details on the calculation of *R*_0_ are provided in the Supplementary Information. Notably, in each behavioral model, *R*_0_ remains the same as in the baseline. Indeed, they all converge to the baseline in a fully susceptible population, where behavioral effects are negligible, such as during the epidemic’s early phase.

As shown in Fig. 3, we observe comparable posterior distributions in some instances, as in the case of London, where the three models estimate median and 90%*CI* for *R*_0_ values of 2.54 [2.29, 2.90] (DDB), 2.77 [2.43, 3.20] (CBF), and 2.38 [2.09, 2.68] (EFB). The posterior distributions of the three models are similar (to different extents) also for Jakarta, Madrid, and New York. However, we also find significant variations. For instance, in Santiago de Chile, the Data-Driven Behavioral model estimates a *R*_0_ of 4.20 [3.96, 4.42], whereas the estimates from the Compartmental and the Effective Force of Infection Behavioral Feedback models are notably lower, at 1.87 [1.69, 2.04] and 1.77 [1.59, 1.97], respectively. To quantify the similarity among *R*_0_ distributions, we employ the Wasserstein distance. On average, we find that the distributions projected by the two Analytical Behavioral Feedback Models (CBF and EFB) are closer to each other compared to the distribution projected by the Data-Driven Behavioral model, which tends to exhibit greater dissimilarity. We refer the reader to the Supplementary Information for full details on *R*_0_ distributions and related analysis.

**Figure 3.**
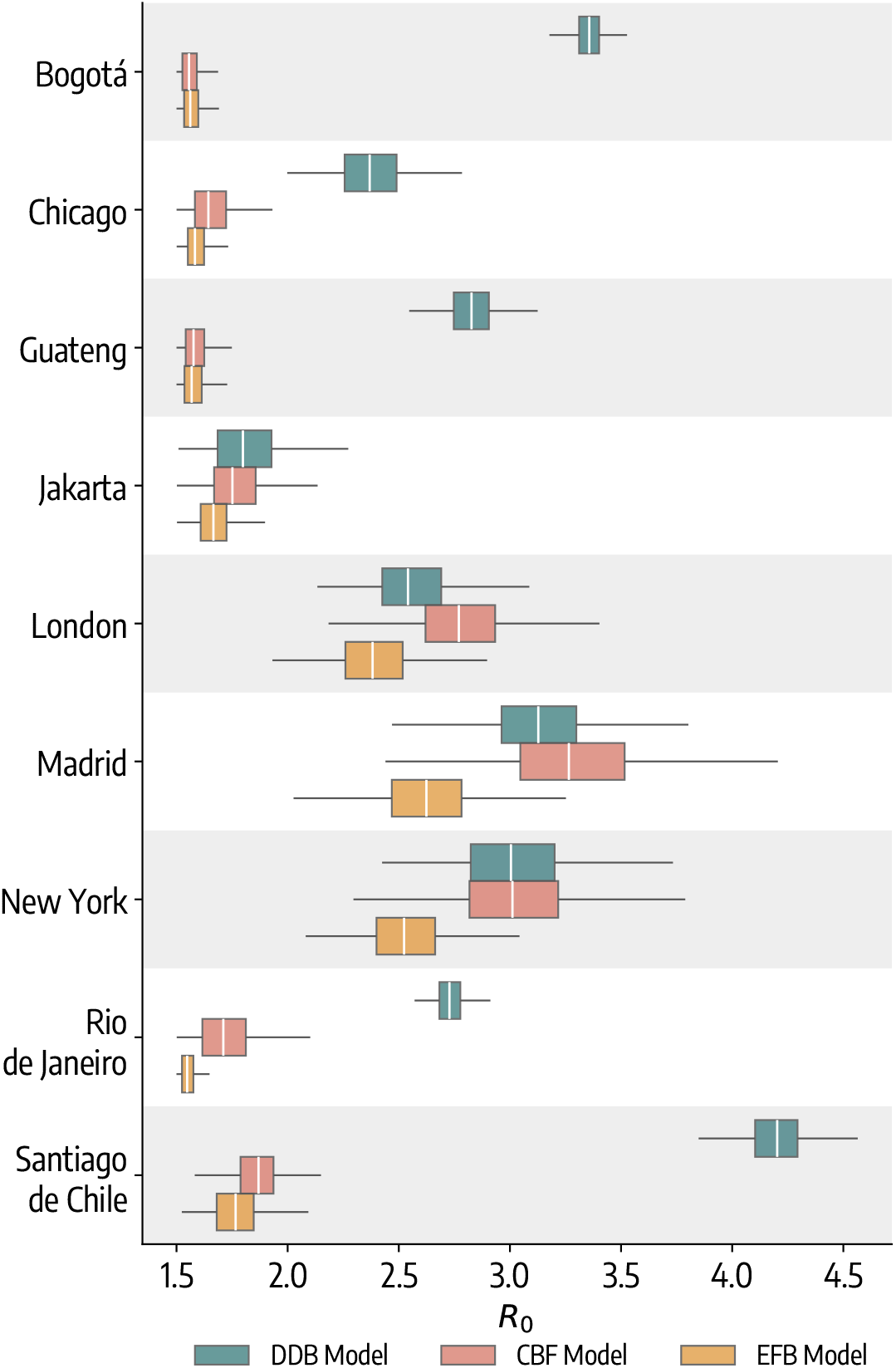
Boxplot of *R*_0_ posterior distributions according to the three models considered across the nine geographies, considering 1, 000 posterior samples. The box boundaries represent the interquartile range (IQR) between the first and third quartiles (Q1 and Q3), the line inside the box indicates the median and the upper (lower) whisker extends to the last datum less (greater) than *Q*3 +1.5*IQR* (*Q*1 − 1.5*IQR*). DDB stands for Data-Driven Behavioral model, CBF for Compartmental Behavioral Feedback model, and EFB for Effective Force of Infection Behavioral Feedback model.

While *R*_0_ is of epidemiological significance, the effective reproductive number *R*_*t*_ emerged as a crucial and closely monitored metric during the COVID-19 pandemic. Unlike *R*_0_, *R*_*t*_ accounts for fluctuations in transmissibility attributed to seasonality, changes in susceptibility, and also behavioral changes. For this reason, in Fig. 4 we compare the *R*_*t*_ estimated using the method described in Ref. [56] from the three models’ data. Generally, we observe a consistent trend in their evolution. Noteworthy is the close alignment of tipping points (i.e., instances where *R*_*t*_ crosses 1) projected by all three models. However, deviations are evident in the cases of Bogotá and Gauteng, where the Data-Driven Behavioral model predicts an early tipping point in March/April 2020. This divergence can be attributed to decreased mobility in those regions during the early months of 2020, prompted by global emergency measures, despite a subsequent rise in cases and deaths. This underscores some limitations of mobility data in accurately estimating the impact of NPIs (see the Discussion section). As an additional analysis, we calculate pairwise correlations between one-step changes in *R*_*t*_ as estimated by the three models (shown in the inset of Fig. 4). We generally observe positive and statistically significant correlations, with a few exceptions. For instance, the evolution of *R*_*t*_ in the Data-Driven Behavioral model does not show a significant correlation with the corresponding quantity estimated by the Compartmental Behavioral Feedback model and the Effective Force of Infection Behavioral Feedback model in Gauteng. Similarly, the *R*_*t*_ of the Data-Driven Behavioral model is not significantly correlated with that of the Compartmental Behavioral and of the Effective Force of Infection Behavioral Feedback model in Santiago de Chile. Similar to the findings regarding *R*_0_ distributions, we note that the evolution of *R*_*t*_ projected by the two Analytical Behavioral Feedback Models generally shows higher correlations.

**Figure 4.**
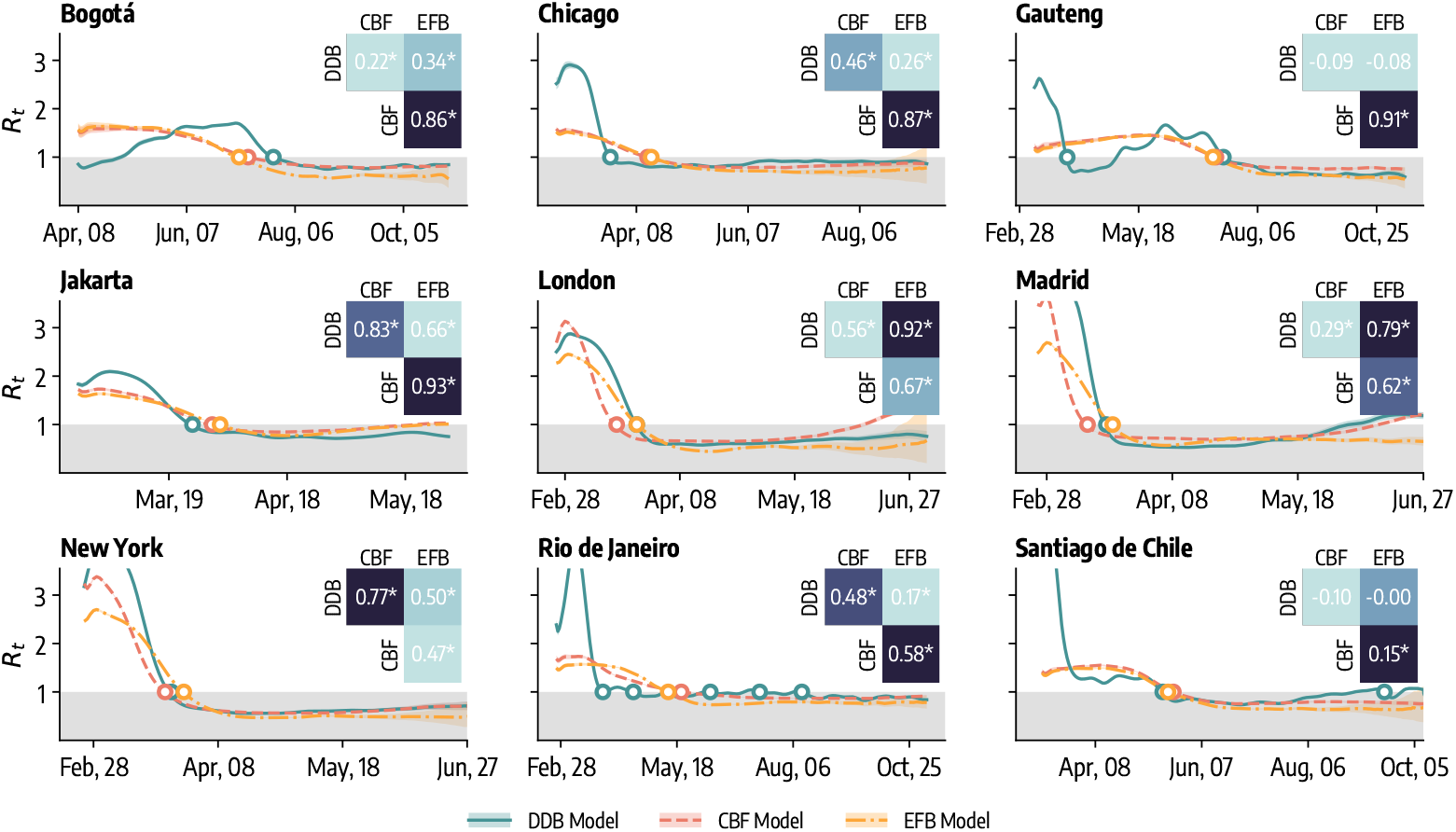
Effective reproductive number (*R*_*t*_) analysis. Effective reproductive number (median, 90% predictive intervals) projected by three epidemic-behavior models across nine geographical regions considered. Shaded grey area indicates *R*_*t*_ *<* 1, while dots indicate points in time where *R*_*t*_ went below 1. In the inset of each figure, we show pairwise Pearson correlation coefficients between one-step changes in *R*_*t*_ as estimated by the three models. Asterisks indicate correlations significant at the 5% significance level. DDB stands for Data-Driven Behavioral model, CBF for Compartmental Behavioral Feedback model, and EFB for Effective Force of Infection Behavioral Feedback model.

Overall, our results highlight an important point. Differences in the estimated values of *R*_0_ and *R*_*t*_ across models are influenced by approaches used to describe the force of infection. Hence, they are affected by different underlying assumptions and approximations. Ensemble approaches, which average across models, might be used to provide more reliable estimations of the real value of such quantities [57]. While these methods are typically used in out-of-sample forecasts, they have been also used to provide in-sample consensus estimates across models for *R*_*t*_ [58].

### Forecasting performance

As a final step in comparing the three models, we use them to forecast the number of weekly deaths during the first wave in the nine geographies under consideration. It is important to note that in assessing forecast performance, the role of model complexity and the number of parameters remains unclear. Especially at the early stage of an epidemic wave, limited available data can disadvantage more complex models, which may be outperformed by more parsimonious approaches. Specifically, we calibrate each model up to time *t*, forecast the subsequent four weeks, then shift the calibration window to *t* + 1 and repeat the process. We assess forecasting performance using two metrics: the Weighted Interval Score (WIS) and the mean absolute error (MAE) of the median. Normalization of metrics is not required in this context, as forecasting performance is assessed relative to a baseline model, as explained below. For simplicity, we present analysis concerning the WIS as a performance metric in the main text, while MAE results are reported in the Supplementary Information. The main findings remain consistent across both metrics.

In Fig. 5A, we present the ratio, for all forecasting rounds, between the average WIS of each model over the 4-week horizon and the average WIS of a baseline model. The latter is defined as a model that consistently predicts, as median value, the last data point within the calibration period and whose predictive intervals are estimated on past data. Similar baseline models have become a standard neutral benchmark providing a simple reference for all models in the context of collaborative forecasting hubs, such as the US and the European COVID-19 Forecast Hub [57, 59]. We refer the reader to the Material and Methods for more details and the definition of the baseline model. It follows that, values below (above) 1 indicate superior (inferior) performance compared to the baseline. We observe heterogeneous forecasting performance among the geographies under consideration. Notably, in Chicago, Gauteng, London, Madrid, New York, and Santiago de Chile, all models statistically outperform the baseline. However, in other locations, some models significantly underperform compared to the baseline; for instance, the Effective Force Behavioral Model in Rio de Janeiro and the Data-Driven Behavioral Model in Bogotá. We use the Wilcoxon signed-rank test (with outliers removal) to statistically compare the performance of different models. The null hypothesis of this test is that the two groups come from the same distribution. In Fig. 5A we report the statistical significance of the tests comparing different pairs of models as follows: ****: *p*_*value*_ ≤ 10^−4^, ***: 10^−4^ *< p*_*value*_ ≤ 10^−3^, **: 10^−3^ *< p*_*value*_ ≤ 10^−2^, *: 10^−2^ *< p*_*value*_ ≤ 0.05, and otherwise blank if *p*_*value*_ *>* 0.05. The Data-Driven Behavioral Model performs best in terms of median relative WIS in Chicago, Jakarta, London, New York, Rio de Janeiro and Santiago de Chile. However, its performance distribution is statistically different from that of the Compartmental Behavioral Model only in three cases (Chicago, London, and New York). The Compartmental Behavioral Model is the median top performer in the remaining three locations. Its performance distribution is statistically different from that of the Data-Driven model in all these three cases, namely Bogotá, Gauteng, and Madrid.

**Figure 5.**
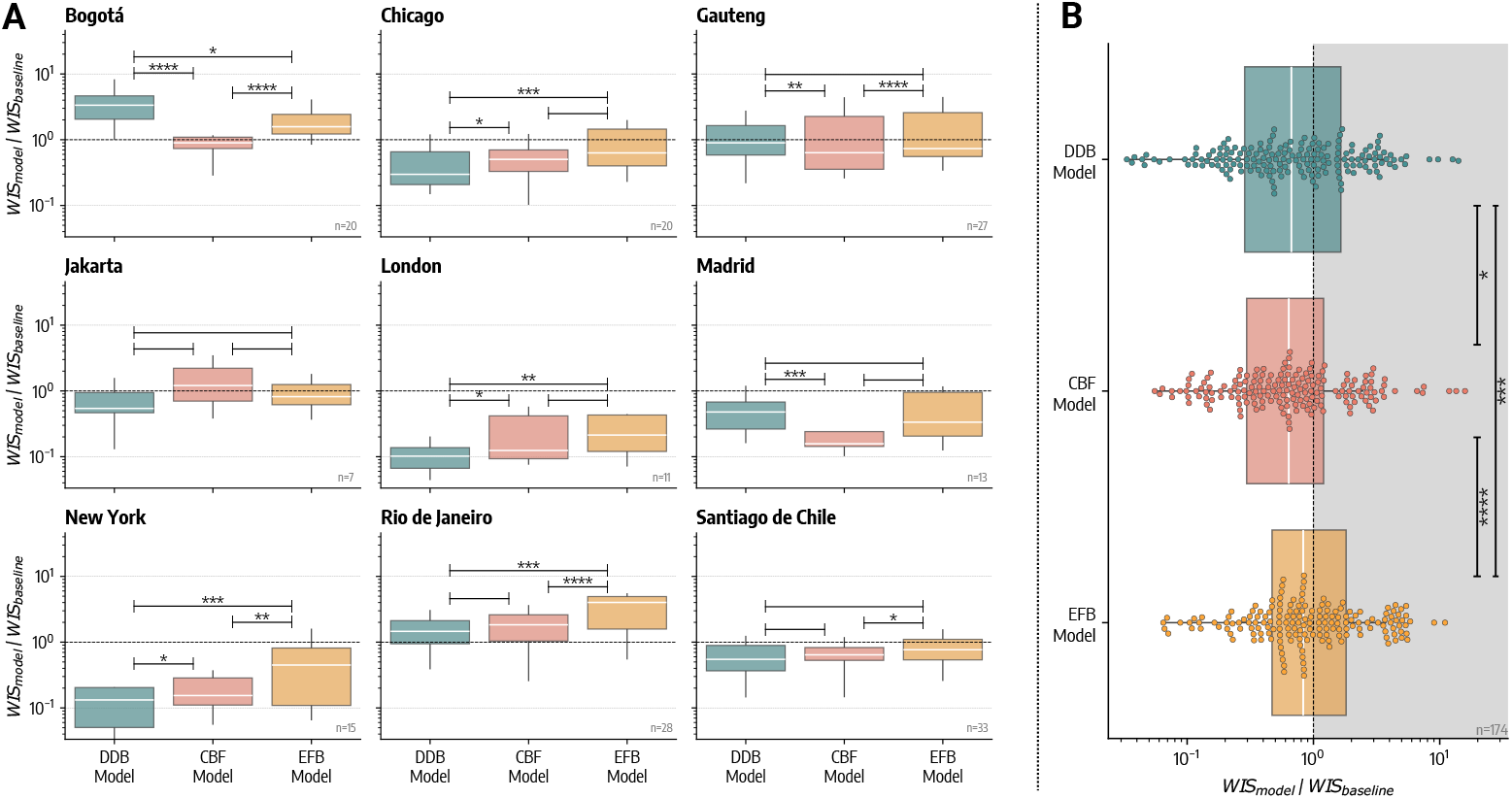
Forecasting performance (WIS). A) Relative WIS computed over all forecasting rounds for the three epidemic-behavior models across the nine geographical regions considered. Values below 1 indicate better performance with respect to baseline forecasting model. Each data point underlying the boxplot represents the relative WIS averaged over the four-week horizon of the corresponding forecasting round. In the bottom right of each plot, we report the number of forecasting rounds for each location. B) Boxplot and swarmplot of relative WIS for different models pooling together results from all rounds and geographies. The box boundaries represent the interquartile range (IQR) between the first and third quartiles (Q1 and Q3), the line inside the box indicates the median and the upper (lower) whisker extends to the last datum less (greater) than *Q*3 + 1.5*IQR* (*Q*1 − 1.5*IQR*). DDB stands for Data-Driven Behavioral model, CBF for Compartmental Behavioral Feedback model, and EFB for Effective Force of Infection Behavioral Feedback model. In both panels we report the statistical significance of the Wilcoxon test comparing different forecasting performances as follows: ****: *p*_*value*_ ≤ 10^−4^, ***: 10^−4^ *< p*_*value*_ ≤ 10^−3^, **: 10^−3^ *< p*_*value*_ ≤ 10^−2^, *: 10^−2^ *< p*_*value*_ ≤ 0.05, and otherwise blank if *p*_*value*_ *>* 0.05.

To provide an overall performance assessment, in Fig. 5B, we show the distribution of the relative WIS with respect to the baseline model for the three models. To provide an overall view of models’ performance, we combine results from all geographies and all forecasting points. The analysis shows that the Compartmental Behavioral Feedback model is statistically the top performer with an overall median relative WIS of 0.64, closely followed by the Data-Driven Behavioral model (0.67) and the Effective Force of Infection Behavioral Feedback model (0.83). The reported significance of the Wilcoxon test confirms the statistical difference in performance between the Compartmental Behavioral Model and the other two models. Furthermore, for this model, nearly 70% of the forecasts are better than the baseline compared to only the 60% and 58% for the Data-Driven and the Effective Force of Infection Behavioral Feedback model, respectively.

In the Supplementary Information, we present the forecasting performance analysis for two ensemble models that combine the forecasts of individual epidemic-behavior models. Ensemble forecasts have consistently demonstrated greater accuracy and reliability over time in various epidemiological forecasting contexts [57, 59]. In the first ensemble approach, all models are weighted equally, while in the second, weights are proportional to past forecasting performance. Interestingly, we find that ensemble models outperform individual epidemic-behavior models in most cases across different metrics. Although the performance-weighted ensemble shows slight improvements over the equally weighted ensemble, these improvements are generally marginal and not statistically significant.

## 3 Discussion

Modeling the interplay between human behavior and the spread of infectious diseases is still considered a *hard problem* in epidemiology [24, 27, 28, 60]. One of the main obstacles to solving this challenge has been the lack of data to validate the theoretical models developed. A review of studies published between 2010 and 2015 found that only about 15% utilized data to parameterize and/or validate the proposed epidemic-behavior mechanisms [27]. The COVID-19 pandemic has significantly altered this landscape. The abundance of novel data sources and the scale of the emergency made incorporating behavioral data into modeling studies not only possible but essential. Most computational models, however, have leveraged data to include behavioral changes as an exogenous factor, as seen in the Data-Driven Behavioral Feedback Model used here. While this approach benefits from a transparent data integration process, it has limited the use and validation of general classes of models that explicitly simulate the feedback loop between the spread of infectious diseases and human behaviors.

Interestingly, our results show that Analytical Behavioral Feedback Models, developed well before the COVID-19 pandemic, often provide comparable or superior performance to data-driven approaches in both retrospective analysis and forecasting. Although we observe variability depending on the task, geographical context, and the metrics considered, our findings suggest that purely data-driven methodologies to model behavior change may not always represent the best modeling solution.

While it may seem counter-intuitive to suggest that data-driven approaches based on mobility data are less reflective of the actual dynamics of an epidemic, several factors contribute to this conclusion. In many cases, there is limited knowledge about the underlying population or the data generation process, complicating the assessment of data representativeness and potential biases. Additionally, for forecasting purposes, real data can only be used under specific assumptions about the future —often with a status quo assumption— which may not accurately capture the true dynamics of the population. Additionally, there is limited mobility data available for many low and middle-income countries, and when available, the data quality can be poor [61–64]. This can easily deteriorate the modeling results [65]. These issues can be mitigated by using analytical behavioral feedback models, which derive the epidemic-behavior interplay in a self-consistent way through calibration and parameterization of behavioral mechanisms. However, purely model-based approaches can misinterpret errors in reporting, the emergence of more transmissible virus strains, or other factors influencing the epidemic’s progression as changes in behavior. Moreover, the high-dimensional space characterizing complex analytical behavioral feedback models can lead to challenges such as parameter identifiability, interpretability, and overfitting.

Our results stress the critical importance of systematically evaluating different modeling approaches and considering the use of ensemble modeling techniques. Indeed, as shown in the Supplementary Information, by integrating the three models, ensemble methods generally improve forecast accuracy and generate a broader range of potential epidemic trajectories. Since no single model can completely capture the feedback mechanisms of epidemic dynamics and human behavior, ensembling multiple models allows for the integration of different mechanisms and factors, potentially resulting in a more reliable representation of the epidemic-behavior interplay [66, 67].

As with all modeling approaches, this study is not without limitations. First, we could not consider all the possible approaches used to model COVID-19 epidemic-behavior interplay. For instance, we did not test the performance of survey-driven models [68–71] and semi-mechanistic models [72, 73].

Furthermore, translating mobility changes into contact reductions remains an open challenge. Therefore, the performance of the data-driven model may vary depending on different approaches to effective contacts rescaling [74–78]. Finally, our forecasts are based on data reported as of today, not addressing the challenge of retrospective data adjustments (i.e., backfilling) very common for epidemiological datasets. While this could impact forecast performance, our primary goal is to compare models. Therefore, assuming that all models would be equally affected, this issue may not significantly impact our comparative analysis.

In light of the assumptions and limitations, our study offers a clear path forward for the application of analytic feedback behavioral models in both retrospective analysis and epidemic forecasting [59, 79, 80]. The strong performance of the Compartmental Behavioral Feedback model in both tasks suggests that the epidemic-behavior interplay can be mechanistically captured in a parsimonious way, substantially improving accuracy. Moreover, our study paves the way for more systematic use of analytic feedback behavioral models in operational forecasting efforts. By demonstrating their capability to account for the dynamic relationship between human behavior and disease spread, these models can improve epidemic forecasts and projections without relying on explicit assumptions about the possible evolution of population behavior. This approach can be particularly valuable in anticipating the effects of public health interventions and supporting more informed decision-making.

## 4 Materials and methods

### 4.1 Epidemic models

#### 4.1.1 Compartmental and age structure

In all models studied, we adopt a SEIR compartmentalization setup. Healthy and susceptible individuals are placed in the compartment *S*. Through interactions with infectious, they transition to the compartment of the exposed *E*. Individuals in the *E* compartment get infectious only after the latent period (*ϵ*^−1^) when they transition to the compartment *I*. Finally, after the infectious period (*μ*^−1^) individuals in the *I* compartment transition to the compartment of the recovered *R*. We assume the population to be stratified into 10 different age groups ([0−9, 10−19, 20−24, 25−29, 30−39, 40−49, 50−59, 60−69, 70−79, 80+]). We introduce the contact matrix **C** ∈ ℝ^*K×K*^, whose element *C*_*i,j*_ is the average number of daily contacts that an individual in age group *i* has with individuals in age group *j* [81]. The rate at which susceptible individuals acquire infection, namely the force of infection, is 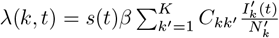 (where *s*(*t*) is a seasonality modulation term and 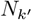 is the number of individuals in age group *k*′ such that 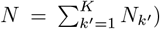, and the basic reproductive number for this model is 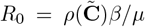, where 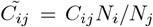 and 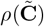 is the spectral radius of the matrix. Our implementation of the model is stochastic and the number of individuals transitioning among compartments is simulated via chain binomial processes. Additionally, we model COVID-19 deaths by applying the age-stratified infection fatality rates [82] to the number of individuals transitioning from *I*_*k*_ to *R*_*k*_ and accounting for a lag Δ between such transition and actual death due to isolation, hospitalization, and reporting delays. More details on the model definition are provided in the Supplementary Information.

#### 4.1.2 Data-Driven Behavioral Model

In the Data-Driven Behavioral model, we use the COVID-19 Community Mobility Report published by Google LLC [53] to modulate the force of infection. This dataset reports percentage changes in mobility to specific locations on a given day and geography. Our models do not consider multiple locations, so we derive an overall mobility change percentage *m*(*t*) as the average of mobility changes towards all locations (excluding mobility towards parks due to its anomalous behavior). Finally, *m*(*t*) is turned into a contacts reduction parameter as follows: *r*_*mobility*_(*t*) = (1 −|*m*(*t*)|*/*100)^2^. The intuition is that, under the homogeneous mixing assumption, the number of contacts will be proportional to the square of the number of individuals. Then, we use *r*_*mobility*_(*t*) to modulate the rate at which susceptible becomes infected as a consequence of behavior change, namely, we modify the force of infection as *λ*′(*k, t*) = *r*_*mobility*_(*t*)*λ*(*k, t*). In short-term forecasting, we assume that future *r*_*mobility*_(*t*) will be equal to the last observed contacts reduction parameter in the calibration window (i.e., status quo assumption).

#### 4.1.3 Compartmental Behavioral Feedback Model

In the Compartmental Behavioral Feedback model, we introduce an additional compartment 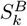 of susceptible individuals that adopt behavior change and thus get infected at a lower rate *rλ*(*k, t*), where *r <* 1 is a parameter that describes the efficacy of preventive measures. The transition from *S*_*k*_ to 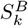 happens at rate 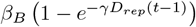, where the *β*_*B*_ and *γ* regulate the behavioral response, and *D*_*rep*_(*t*−1) is the total number of reported deaths in the previous day. We also assume that 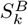 individuals can relax their behavior and transition back to *S*_k_ at a rate 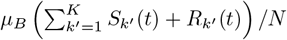, where *μ*_B_ set the tendency of susceptibles to drop safer behaviors [83].

#### 4.1.4 Effective Force of Infection Behavioral Feedback Model

In the Effective Force of Infection Behavioral Feedback model, we consider the following function

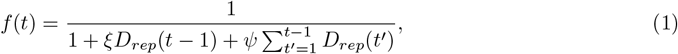

where, as above, *D*_*rep*_ (*t* − 1) is the number of reported deaths at time 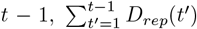) is the cumulative number of reported deaths up to *t* − 1, *ξ* and *ψ*are parameters that set the behavioral reactivity of individuals [51]. This function multiplies the force of infection and serves as a proxy for the modulating effect of behavioral changes. Specifically, it takes into account new reported deaths in the last time step, capturing short-term effects of recent epidemiological conditions on behavior, as well as cumulative reported deaths, capturing the long-term effects of past epidemiological conditions on current behavior.

### 4.2 Models calibration

Models are calibrated using an Approximate Bayesian Computation - sequential Monte Carlo (ABC-SMC) algorithm [54, 84]. The ABC-SMC is an extension of the more simple rejection algorithm, which works as follows. The modeler needs to choose prior distribution *π*(***θ***) for the free parameters ***θ*** of the model, a distance metric *d*(*·*), a tolerance *δ*, and a population size *P*. Then, the model is run iteratively sampling at each step a parameters set ***θ***_*i*_ from the prior distribution *π*(***θ***). At each iteration an output quantity produced by the model *y*_*i*_ (i.e., simulated deaths) is compared to the corresponding real quantity *y*_*obs*_ using the distance metric *d*(*y*_*i*_, *y*_*obs*_). If *d*(*y*_*i*_, *y*_*obs*_) *< δ* then ***θ***_*i*_ is accepted, otherwise it is rejected. This process continues until *P* parameter sets are accepted. The main limitation of this approach is that the acceptance criterion remains fixed, causing slow convergence. Additionally, finding an appropriate tolerance value *δ* beforehand is challenging, especially with multiple models and geographies. The ABC-SMC algorithm addresses these issues by using a sequence of *T* rejection steps (i.e., *generations*) with decreasing tolerance. Each generation’s prior distribution is the posterior distribution from the previous one perturbed via a kernel function. This method starts with high error tolerances and broad prior distributions, progressively refining the parameter space. The final generation’s accepted ***θ***_*i*_ distribution approximates the true posterior distribution of the parameters. Here, we consider 10 generations, 1, 000 parameter sets accepted at each step, weekly deaths as output quantity, and the weighted mean absolute percentage error (wMAPE) as a distance metric, defined as

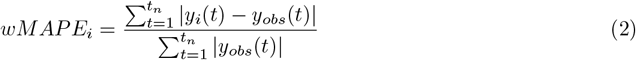

where *y*_*obs*_ is the vector of actuals and *y*_*i*_ of model estimates, and *t*_*n*_ is the number of weeks considered. We use the ABC-SMC implementation of the *pyabc* Python package [85]. In the Supplementary Information, we report additional information on the calibration, including prior distributions.

### 4.3 Performance metrics

The Mean Absolute Error (MAE) of the median is defined as:

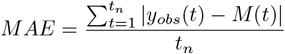

Where *y*_*obs*_ is the vector of actuals and *M* the model’s medians. Its normalized version is simply defined as *nMAE* = *MAE/mean*(*y*_*obs*_).

The Weighted Interval Score (*WIS*) is a score that approximates the continuous ranked probability score (*CRPS*) [86]. For a given a prediction interval (1 − *α*) *×* 100% (i.e., 90% interval) of a model’s estimate, the interval score (*IS*_*α*_) is defined as:

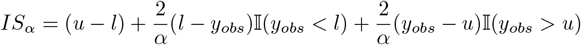

Where *u* (*l*) is the upper (lower) limit of the prediction interval, *y*_*obs*_ is the actual outcome, and 𝕀(*c*) is an indicator function that equals 1 if condition *c* is met and 0 otherwise. Looking at *IS*_*α*_ we see that its first term captures how wide is the prediction interval, while the second and third terms are the penalization for under and over-prediction. Indeed, they are different from 0 only if the actual data point *y*_*obs*_ is below or above the interval limits. The *WIS* is an extension of the *IS* and takes into account multiple prediction intervals at once. It is defined as:

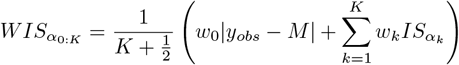

Where *K* is the number of prediction intervals considered, *M* is the model’s median, and *w*_*k*_ are the non-negative weights of the different intervals. We can see that, the first term in parenthesis measures how much the model’s central estimate *M* differs from the actual data *y*_*obs*_, while the second term is a weighted sum of the different interval scores 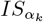. Following a common approach, we set *w*_0_ = 1*/*2, *w*_*k*_ = *α*_*k*_*/*2, and we consider 11 prediction intervals (*α*_*k*_ = 0.02, 0.05, 0.10, 0.20, 0.30, 0.40, 0.50, 0.60, 0.70, 0.80, 0.90). Its normalized version is simply defined as *nWIS* = *WIS/mean*(*y*_*obs*_).

The Bayesian Information Criterion (*BIC*) is a metric that evaluates the model’s estimates based on the accordance with real data and on the model complexity [87]. For a model *q* it is defined as:

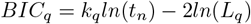

Where *k*_*q*_ is the number of model’s free parameters, and *L*_*q*_ is the model’s likelihood which we define it here as

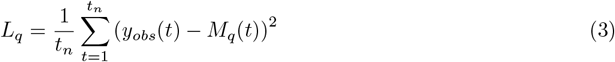

the mean squared error between actual (*y*_*obs*_) and median predicted values (*M*_*q*_). Intuitively, the model reaching the lowest BIC is the best, since it guarantees the minimum deviation from observed data with the minimum number of parameters. In this sense, *BIC* favors both accordance with real data and the model’s parsimony, however, its values lack an immediate interpretation. For this reason, we consider BIC weights defined as:

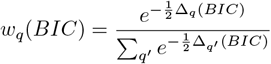

Where 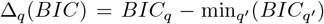. These weights express the relative probability of a model over the others.

### 4.4 Forecasting

In each *forecasting round*, we calibrate the three models using data up to time *t* and we forecast weekly deaths in the next four weeks. In the next round, we move our window up to *t* + 1 and we repeat the calibration and forecasting procedures. This process is performed iteratively until the end of the epidemic curve, starting with at least 4 data points for model calibration. Instead of the ABC-SMC algorithm, for forecasting, we adopt a modified version of the rejection algorithm where, instead of setting a predefined tolerance, we calibrate models by selecting top 1, 000 simulations out of a total of 1*M* simulations obtained through sampling from the prior distributions. In the case of forecasting, we also consider as distance metric a generalized version of the wMAPE which gives more importance to more recent data points defined as 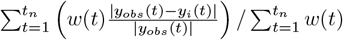, where *w*(*t*) = 1*/*((*t*_*n*_ + 1) − *t*).

### 4.4.1 Baseline forecasting model

We employ a baseline forecasting model that consistently predicts the median value as the last data point within the calibration period. To compute predictive intervals, we consider the previous 1-step increments. Specifically, we compute 1-step differences up to time *t*: *δ* = (*d*_2_, *d*_3_, …, *d*_*t*_). To ensure the median forecast aligns with the last calibration point, we symmetrize *δ* by considering *δ*′ = (*δ*, −*δ*). If the maximum horizon is *H*, we sample *H* differences from *δ*′. Finally, predictions at horizon *h* are computed as: 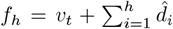, where *v*_*t*_ represents the last observed data points. Following this process, we generate 10, 000 trajectories from which we compute quantiles and predictive intervals.

## Data Availability

All data produced in the present study are available upon reasonable request to the authors

## Acknowledgments

N.G. acknowledge support from the Lagrange Project of the Institute for Scientific Interchange Foundation (ISI Foundation) funded by Fondazione Cassa di Risparmio di Torino (Fondazione CRT). A.V. acknowledges support from the HHS/CDC-5U01IP000113 and the CDC-RFA-FT-23-0069 cooperative agreement from the CDC’s Center for Forecasting and Outbreak Analytics. The findings and conclusions in this study are those of the authors and do not necessarily represent the official position of the funding agencies. Any use of trade, firm, or product names is for descriptive purposes only and does not imply endorsement by the U.S. Government.

