## Supplementary Information for "Comparative Evaluation of Behavioral-Epidemic Models Using COVID-19 Data"

**2 Model calibration 7**

**3 Comparison of models performance in retrospective modeling task: peak intensity and timing 10**

**4 $R_0$ posterior distributions 11**

**5 Forecasting performance: Mean Absolute Error 11**

**6 Forecasting performance by Horizon 13**

**7 Forecasting performance of ensemble models 14**

**8 Posterior distributions in time** **18**

**9 N-week ahead forecasts** **20**

### 1 Epidemic-behavior models

#### 1.1 Baseline epidemic model

We adopt a SEIR compartmentalization setup to model COVID-19 disease progression. We assume the population to be stratified into 10 different age groups $k$ ($[0-9, 10-19, 20-24, 25-29, 30-39, 40-49, 50-59, 60-69, 70-79, 80+]$), each with population $N_k$ representing the demographic structure of the population of interest (see Tab. 1 for sources of demographic and epidemiological in different locations). For each geographical locations we also consider the contact matrix $\mathbf{C} \in \mathbb{R}^{K \times K}$, whose element $C_{i,j}$ is the average number of daily effective contacts that an individual in age group $i$ has with individuals in age group $j$ [1].

The rate at which healthy and susceptible individuals $S$ transition to the exposed $E$ state, namely the force of infection, is:

$$\lambda(k,t) = \beta s(t) \sum_{k'=1}^{K} C_{kk'} \frac{I_{k'}(t)}{N_{k'}}, \tag{1}$$

where the rate of infection is assumed to be proportional to the fraction of infectious individuals in each age group and $\beta$ is the infection trasmissibility. The expression also accounts seasonality modulation $s(t)$ to account for variations in humidity, temperature, and other factors that may impact both transmissibility and contact patterns [2, 3]. This term is equal to:

$$s_i(t) = \frac{1}{2}\left[\left(1 - \frac{s_{min}}{s_{max}}\right) \sin\left(\frac{2\pi}{365}(t - t_{max,i}) + \frac{\pi}{2}\right) + 1 + \frac{\alpha_{min}}{\alpha_{max}}\right] \tag{2}$$

where $i$ refers to the hemisphere, and $t_{max,i}$ is the time which corresponds to the maximum of $s_i(t)$. In the northern hemisphere, we set it to January $15^{th}$, and six months later in the southern hemisphere. We assume no typical seasonality in the tropical region. We set $s_{max} = 1$ and consider $s_{min}$ as a free parameter [3, 4].

Exposed individuals in the state $E$ progress to the infectious stage $I$ at a rate $\epsilon$ inversely proportional to the latent period, and infectious individuals progress to the removed stage $R$ at a rate $\mu$ inversely proportional to the infectious period. Removed individuals are those who can no longer infect others. We simulate the disease progression by using stochastic chain binomial processes with the number of individuals transitioning on day $t$ from compartment $X_k$ to $Y_k$ is sampled from a binomial distribution $Bin(X_k(t), p_{X_k \to Y_k, t})$, where $p_{X_k \to Y_k, t}$ is the transition probability at time $t$. This yields the following stochastic processes that can be conveniently iterated computationally:

$$
\begin{aligned}
S_k(t+\delta t) &= S_k(t) - Bin(S_k(t), \lambda_r(k,t)) \\
E_k(t+\delta t) &= E_k(t) + Bin(S_k(t), \lambda_r(k,t)) - Bin(E_k(t), \epsilon_r) \\
I_k(t+\delta t) &= I_k(t) + Bin(E_k(t), \epsilon_r) - Bin(I_k(t), \mu_r) \\
R_k(t+\delta t) &= R_k(t) + Bin(I_k(t), \mu_r),
\end{aligned}
\tag{3}
$$

where $\lambda_r(k,t)$, $\epsilon_r$, and $\mu_r$, are the transition probability obtained by transforming rates into risk with $risk = 1 - \exp(-rate \times \delta t)$. We set the unitary time scale $\delta t$ equal to 1 day and we run simulations with a smaller integration step of $\delta t = 1/12$ (approximately 2 hours) to have that $risk \simeq rate$.

For the above contagion process, the basic reproduction number is $R_0 = \rho(\tilde{\mathbf{C}})\beta/\mu$, where $\tilde{C_{ij}} = C_{ij}N_i/N_j$, $\rho(\cdot)$ is the spectral radius [5], and the generation time of the disease is $T_G = \epsilon^{-1} + \mu^{-1}$. It is important to note that, in the settings considered here, the $R_0$ in all behavioral models studied is the same as in the baseline which does not include behavioral changes.

In order to model the outcome of the disease and more specifically COVID-19 deaths, we consider the daily transition from $I_k$ to $R_k$. A fraction of this, regulated by age-stratified estimates of the Infection Fatality Rate (IFR) from Ref. [6], transitions to the compartment $D_k$ that accounts for individuals that die. To account for delays between the transition $I_k \rightarrow R_k$ and actual death due to isolation, hospitalization, and reporting delays we introduce the parameter $\Delta$. Deaths computed on the number of recovered of day $t$ are recorded at day $t+\Delta$ in our simulations. In practice, we use $\Delta$ to move individuals from compartment $D_k$ to a new compartment $D_{k,rep}$ (where the superscript *rep* stands for *reported*) that accounts for deaths on the day of reporting. Finally, we also consider a deaths under-reporting factor $\alpha$. This implies that the number of simulated deaths is multiplied by $\alpha$ to account for the fraction of deaths that are reported. In the following, when we refer to a compartment by name without the age subscript $k$, we denote the total count across all age groups. For example $D_{rep} = \sum_{k'=1}^{K} D_{k',rep}$.

#### 1.2 Data-Driven Behavioral (DDB) model

In the Data-Driven Behavioral (DDB) model we integrate Community Mobility Report published by Google LLC [7] to define an effective parameter that we use to modulate the force of infection. This dataset reports percentage changes in mobility/visits to specific locations on a given day and geography. Our models do not consider multiple locations (i.e., contexts), so we introduce an overall parameter $m(t)$ defined as the average percent reduction of visits towards all locations (we only exclude mobility towards parks due to its anomalous behavior). Finally, $m(t)$ is turned into an effective contact reduction parameter $r_{mobility}(t) = (1 - |m(t)|/100)^2$. The intuition behind this formula is that, under the homogeneous mixing assumption, the number of potential contacts $C$ is proportional to the square of the number of

| Location | Demographic Data Source | Epidemiological Data Source |
|---|---|---|
| *Bogotá* | Observatorio de Salud de Bogotá, Población de Bogotá [8] | Gov.co Datos Abiertos, Casos positivos de COVID-19 en Colombia [9] |
| *Chicago* | Census Reporter, ACS 2022 1-year, Total Population [10] | Chicago Data Portal, Daily Chicago COVID-19 Cases, Deaths, and Hospitalizations - Historical [11] |
| *Gauteng* | Coronavirus COVID-19 (2019-nCoV) Data Repository for South Africa, Provincial projection by sex and age [12] | Coronavirus COVID-19 (2019-nCoV) Data Repository for South Africa [13] |
| *Jakarta* | Population by Age Group and Sex in DKI Jakarta Province, 2020 [14] | Daily Update Data Agregat Covid-19 Jakarta [15] |
| *London* | Office for National Statistics, Estimates of the population for the UK, England, Wales, Scotland, and Northern Ireland [16] | Coronavirus (COVID-19) Weekly Update, Greater London Authority (GLA) [17] |
| *Madrid* | Instituto Nacional de Estadistica, Población por comunidades, edad (grupos quinquenales), Españoles/Extranjeros, Sexo y Año [18] | Ministerio de Sanidad, COVID-19 Deaths [19] |
| *New York* | United States Census Bureau, Age and Sex [20] | NYC Health COVID-19 Data [21] |
| *Rio de Janeiro* | Instituto Brasileiro de Geografia e Estatística, Population Projection [22] | Ministério da Saúde, Coronavirus Brazil [23] |
| *Santiago de Chile* | Instituto Nacional de Estadisticas, Proyecciones de población [24] | Departamento de Estadísticas e Información de Salud, COVID-19 Open Data [25] |

Table 1: Demographic and epidemiological data sources in the nine geographical areas considered in the modeling study.

individuals; i.e. $C = N(N-1)/2 \sim N^2$. The reduction $N'(t) = (1 - m(t)/100)N$ of individuals in a location thus lead to a number of contacts be $C'(t) = N'(t)(N'(t)-1)/2 \sim N'(t)^2 = (1-m(t)/100)^2N^2$ that define the contact reduction parameter $r_{mobility}(t) = C'(t)/C$. The contact reduction parameter is acting on the the force of infection as a factor accounting for the change of contact rate as:

$$\lambda^{DDB}(k,t) = r_{mobility}(t)\beta s(t)\sum_{k'=1}^{K} C_{kk'}\frac{I_{k'}(t)}{N_{k'}}. \tag{4}$$

The DDB model thus uses the very same set of equations described above (see Eq. 3) with mobility data to modulate in time the rate at which susceptible becomes infected, as a consequence of behavior change.

#### 1.3 Effective Force of Infection Behavioral Feedback (EFB) model

In the Effective Force of Infection Behavioral Feedback (EFB) model, we follow an approach similar to the one presented in Ref. [26]. More precisely, We consider the function

$$f(t) = \frac{1}{1 + \xi D_{rep}(t-1) + \psi\sum_{t'=1}^{t-1} D_{rep}(t')}, \tag{5}$$

where $D_{rep}(t-1)$ is the number of new reported deaths at time $t-1$, $\sum_{t'=1}^{t-1} D_{rep}(t')$ is the cumulative number of reported deaths up to $t-1$, and $\xi$, $\psi$ are parameters that set the behavioral reactivity of individuals. This function modulates as a multiplier the force of infection and serves as a proxy for the effects of behavior change, yielding

$$\lambda^{EFB}(k,t) = f(t)\beta s(t) \sum_{k'=1}^{K} C_{kk'} \frac{I_{k'}(t)}{N_{k'}}. \tag{6}$$

More precisely, $f(t)$ accounts for both recent reported deaths, capturing the impact of current epidemiological conditions on behavior (i.e., short-term effect), and cumulative reported deaths, reflecting the influence of past epidemiological conditions on current behavior (i.e., long-term effect). This results in an asymmetry in individuals' behavioral responses, where the same current number of reported deaths may trigger different behaviors depending on the historical context. Also this model uses the very same set of equations reported above (see Eq. 3) with the only difference that the force of infection includes the $f(t)$ term.

#### 1.4 Compartmental Behavioral Feedback (CBF) model

In the Compartmental Behavioral Feedback (CBF) model we introduce an additional state $S_k^B$ identifying susceptible individuals that have decided to adopt a risk aversion behavior. The force of infection acting on individuals in this state is therefore reduced by a factor $r < 1$ which encapsulates the effects of the behavioral changes [27], so that

$$\lambda^{CBF}(k,t) = r\beta s(t) \sum_{k'=1}^{K} C_{kk'} \frac{I_{k'}(t)}{N_{k'}}. \tag{7}$$

The transition from $S_k$ to $S_k^B$ can be modeled through different mechanisms that can provide more importance to local or global information processes [27]. Local mechanisms are modeled via a mass-action law assuming that susceptibles adopt protective behaviors proportionally to the fraction of people affected by the disease. Global mechanisms are instead modelled via a pseudo mass-action law. Hence even a small number of infections or reported deaths can influence the behavior. This mimic effects due to information acquired through media or mandated government changes. Here, we adopt a global mechanism where the transition $S_k \to S_k^B$ happens at rate $\omega(t) = \beta_B \left(1 - e^{-\gamma D_{rep}(t-1)}\right)$, where the two parameters $\beta_B$ and $\gamma$ govern the rate and time scale of behavioral changes, and $D_{rep}(t-1)$ is the number of new reported deaths at time $t-1$. Finally, we can imagine that $S_k^B$ individuals can relax their risk aversion behavior and transition back to $S_k$, once the epidemic conditions improve. This can be achieved in practice by introducing a transition $S_k^B \to S_k$ regulated by the number of non-infected individuals through the following rate $\omega^B(t) = \mu_B \left(\sum_{k'=1}^{K} S_{k'}(t) + R_{k'}(t)\right)/N$, where $\mu_B$ set the rate for susceptibles to return to the regular behavior. In this general setting, the equations describing the epidemic model can be solved using the same chain binomial process approach by extending the equations reported above (see Eq. 3) to include the compartmental transitions related to $S_k^B$, obtaining

$$\begin{aligned}
S_k(t+\delta t) &= S_k(t) - Mult_1(S_k(t), \lambda_r(k,t), \omega_r(t)) - Mult_2(S_k(t), \lambda_r(k,t), \omega_r(t)) + Mult_2(S_k^B, \lambda_r^{CBF}(k,t), \omega_r^B(t)) \\
S_k^B(t+\delta t) &= S_k(t) + Mult_2(S_k(t), \lambda_r(k,t), \omega_r(t)) - Mult_1(S_k^B, \lambda_r^{CBF}(k,t), \omega_r^B(t)) - Mult_2(S_k^B, \lambda_r^{CBF}(k,t), \omega_r^B(t)) \\
E_k(t+\delta t) &= E_k(t) + Mult_1(S_k(t), \lambda_r(k,t), \omega_r(t)) + Mult_1(S_k^B, \lambda_r^{CBF}(k,t), \omega_r^B(t)) - Bin(E_k(t), \epsilon_r) \\
I_k(t+\delta t) &= I_k(t) + Bin(E_k(t), \epsilon_r) - Bin(I_k(t), \mu_r) \\
R_k(t+\delta t) &= R_k(t) + Bin(I_k(t), \mu_r),
\end{aligned} \tag{8}$$

where $\lambda_r(k,t)$, $\lambda_r^{CBF}(k,t)$, $\omega_r(t)$, $\omega_r^B(t)$, $\epsilon_r$, and $\mu_r$, are the transition probability obtained by transforming rates into risk with a small $\delta t$ integration step. Furthermore, $Mult_1(m, p_1, p_2)$ and $Mult_2(m, p_1, p_2)$ indicate a draw from the random variable 1 occurring with probability $p_1$ and the random variable 2 occurring with probability $p_2$, respectively, when there are $m$ trials. It is worth remarking that the multinomial distribution has to be normalized by considering the events with probability $1 - p_1 - p_2$.

##### 1.5 Calibration parameters and priors

The above models share a common set of parameters. In addition, the CBF and EFB models have additional parameters that characterize their dynamic. In the calibration procedure, for each of these parameters, we explore a flat prior informed by the natural history of the COVID-19 disease. In the Table 2 we report the full set of parameters that are used in the calibration of the models together with the assumed priors. We note how we consider two separate prior distributions for deaths detection rate. Based on data from Ref. [28] we categorize regions into high (Bogotá, Chicago, London, Madrid, New York, Rio de Janeiro, Santiago de Chile) and low (Jakarta and Gauteng) reporting ones. We provide a detailed description of the calibration procedure in the following section.

#### 2 Model calibration

The models are calibrated using an Approximate Bayesian Computation Sequential Monte Carlo (ABC-SMC) method [29, 30]. The ABC-SMC algorithm is an extension of the simpler ABC rejection algorithm. In the rejection algorithm, the modeler needs to choose prior distribution $\pi(\boldsymbol{\theta})$ for the set of free parameters $\boldsymbol{\theta}$ of the model, a distance metric $d(\cdot)$, a tolerance $\delta$, and a population size $P$. Then, the model is run iteratively sampling at each step a parameters set $\boldsymbol{\theta}_i$ from the prior distribution $\pi(\boldsymbol{\theta})$.

| | Parameter | Symbol | Prior Distribution |
|---|---|---|---|
| Common parameters | Basic reproductive number | $R_0$ | $Unif(1.5, 6.0)$ |
| | Deaths delay | $\Delta$ | $\hat{Unif}(7, 35)$ |
| | Initial infected | $I_0$ | $\hat{Unif}(10, 10000)$ |
| | Detection rate (High) | $\alpha_H$ | $Unif(50\%, 100\%)$ |
| | Detection rate (Low) | $\alpha_L$ | $Unif(5\%, 70\%)$ |
| | Seasonality parameter | $s_{min}$ | $Unif(0.6, 1.0)$ |
| *CBF model* | Behavior change adoption rate | $\beta_B$ | $Unif(0, 3)$ |
| | Behavior change relaxation rate | $\mu_B$ | $Unif(0, 3)$ |
| | Behavioral sensitivity to infected | $\gamma$ | $LogUnif(-3, 3)$ |
| | Efficacy of behavior change | $r$ | $Unif(0.1, 0.7)$ |
| *EFB model* | Short-term behavior coefficient | $\xi$ | $LogUnif(10^{-4}, 10^0)$ |
| | Long-term behavior coefficient | $\psi$ | $LogUnif(10^{-5}, 10^0)$ |

Table 2: **Prior distributions**. We report the prior distributions for the free parameters used in the ABC-SMC calibration. $Unif(\cdot)$ indicates a uniform prior for continuous variables, $\hat{Unif}(\cdot)$ for discrete ones, and $LogUnif(\cdot)$ indicates a logarithmic uniform prior. We present both the parameters shared across all models and those specific to each model. The Data-Driven Behavioral (DDB) model does not have any additional parameters beyond the common ones.

An output quantity produced by the model $y_i$ (i.e., simulated deaths) at each iteration is compared to the corresponding real quantity $y_{obs}$ using the distance metric $d(y_i, y_{obs})$. According to the rejection algorithm, if $d(y_i, y_{obs}) < \delta$ then $\boldsymbol{\theta}_i$ is accepted, otherwise it is rejected. This process continues until $P$ parameter sets are accepted. The distribution of accepted $\boldsymbol{\theta}_i$ will approximate the true posterior distribution of the parameters. This approach has the advantage of being straightforward and easily parallelizable, nonetheless, it also has several limitations. Indeed, we often do not know, a priori, what is a *good* tolerance $\delta$. Small tolerance values will lead to more accurate results but the calibration will be very slow. On the other hand, higher tolerance values will lead to faster convergence but less accurate results. Furthermore, the prior distribution is never updated to reflect the knowledge acquired from previous iterations. The ABC-SMC algorithm extends the rejection framework to overcome these issues. We report in Algorithm 1 the pseudocode for the ABC-SMC algorithm.

The algorithm consists of $T$ generations. The first one is equal to the rejection algorithm with a very high tolerance value. At the next generation, the tolerance value is decreased and the prior distribution will be constituted by the parameter sets accepted in the previous generation perturbed through a kernel function. This process is repeated for the following generations using increasingly lower tolerances and updating the prior distribution based on parameters accepted in the previous step. The particle sets accepted in the last generation will be the approximation of the posterior of the parameters. This approach has clear advantages, since modelers do not need to define the right tolerance a priori, and

**Algorithm 1** ABC-SMC Algorithm

**Input:**
  $P$: Number of particles
  $\delta$: Sequence of tolerance levels $\delta_1 > \delta_2 > \cdots > \delta_T$
  $\pi(\boldsymbol{\theta})$: Prior distribution
  $f(\cdot|\boldsymbol{\theta})$: Simulator model
  $d(\cdot, \cdot)$: Distance function
  $y_{\text{obs}}$: Observed data
**Output:** Posterior distribution approximations $\{\boldsymbol{\theta}_i^{(t)}\}_{i=1}^{P}$ for each $t = 1, \ldots, T$
**for** $t = 1$ to $T$ **do**
  **if** $t = 1$ **then**
    **for** $i = 1$ to $P$ **do**
      **repeat**
        Sample $\boldsymbol{\theta}_i^{(1)} \sim \pi(\boldsymbol{\theta})$
        Simulate $y_i \sim f(\cdot|\boldsymbol{\theta}_i^{(1)})$
      **until** $d(y_i, y_{\text{obs}}) \leq \epsilon_1$
      Set weight $w_i^{(1)} = \frac{1}{P}$
    **end for**
  **else**
    **for** $i = 1$ to $P$ **do**
      **repeat**
        Sample $\boldsymbol{\theta}_i^{(t-1)} \sim \{\boldsymbol{\theta}_j^{(t-1)}\}_{j=1}^{N}$ with weights $w_j^{(t-1)}$
        Perturb $\boldsymbol{\theta}_i^{(t)} \sim K(\boldsymbol{\theta}|\boldsymbol{\theta}_i^{(t-1)})$
        Simulate $y_i \sim f(\cdot|\boldsymbol{\theta}_i^{(t)})$
      **until** $d(y_i, y_{\text{obs}}) \leq \epsilon_t$
      Compute weight $w_i^{(t)} = \frac{\pi(\boldsymbol{\theta}_i^{(t)})}{\sum_{j=1}^{N} w_j^{(t-1)} K(\boldsymbol{\theta}_i^{(t)}|\boldsymbol{\theta}_j^{(t-1)})}$
    **end for**
  **end if**
  Normalize weights: $w_i^{(t)} \leftarrow \frac{w_i^{(t)}}{\sum_{j=1}^{N} w_j^{(t)}}$ for $i = 1, \ldots, P$
**end for**

information from past iterations is included in the sampling process to speed up the calibration and lead to more accurate results.

Here, we calibrate all models using the ABC-SMC with $T = 10$ generations, a generation population size $P = 1000$, the weighted mean absolute percentage error (wMAPE) as distance metric, and weekly deaths as output quantity. For continuous parameters, we consider a multivariate normal transition kernel with a covariance matrix estimated on previous generation particles, while we consider a discrete transition jump with a transition probability of 0.3 for discrete parameters. Instead of predefining a sequence of tolerances, we begin with $\delta_1 = \infty$. The tolerance for subsequent generations is then dynamically set as the median of the distances of the accepted particles from the previous generation. In Table 2 we report the prior distributions used for different models. We use the python library *pyabc* to implement the ABC-SMC calibration [31].

For forecasting, instead of the ABC-SMC algorithm, we adopt a modified version of the ABC rejection algorithm where, instead of setting a predefined tolerance, we calibrate models by selecting top 1000 simulations out of a total of $1M$ simulations obtained through sampling from the prior distributions. In the case of forecasting, we also consider as distance metric a generalized version of the wMAPE which gives more importance to more recent data points defined as $\sum_{t=1}^{t_n} \left( w(t) \frac{|y_{obs}(t) - y_i(t)|}{|y_{obs}(t)|} \right) / \sum_{t=1}^{t_n} w(t)$, where $w(t) = 1/((t_n + 1) - t)$.

### 3 Comparison of models performance in retrospective modeling task: peak intensity and timing

In Fig. 1, we compare the simulated peak intensity of weekly deaths from the three epidemic-behavior models with the reported data. Interestingly, we find that the Data-Driven model most accurately reproduces the peak intensity in 4 regions, the Effective Force of Infection Behavioral Feedback model in 3, and the Behavioral Compartmental model in the remaining 2. This highlights how the definition of the best model is a function of the metric under scrutiny.

In Fig. 2, we compare the simulated peak timing of weekly deaths from the three epidemic-behavior models against the reported data. Overall, all models demonstrate accuracy in predicting peak timing, with a median deviation of no more than 2 weeks. In Santiago de Chile and New York all models perform equally well in terms of median timing. In Bogotá, Gauteng, Madrid the Data-Driven and the Compartmental Behavioral Feedback model are tied, while in Jakarta and London the Data-Driven and the Effective Force of Infection Behavioral Feedback model are similarly accurate. Finally, in Chicago and Rio de Janeiro the Effective Force of Infection Behavioral Feedback model most closely replicates the observed peak timing.

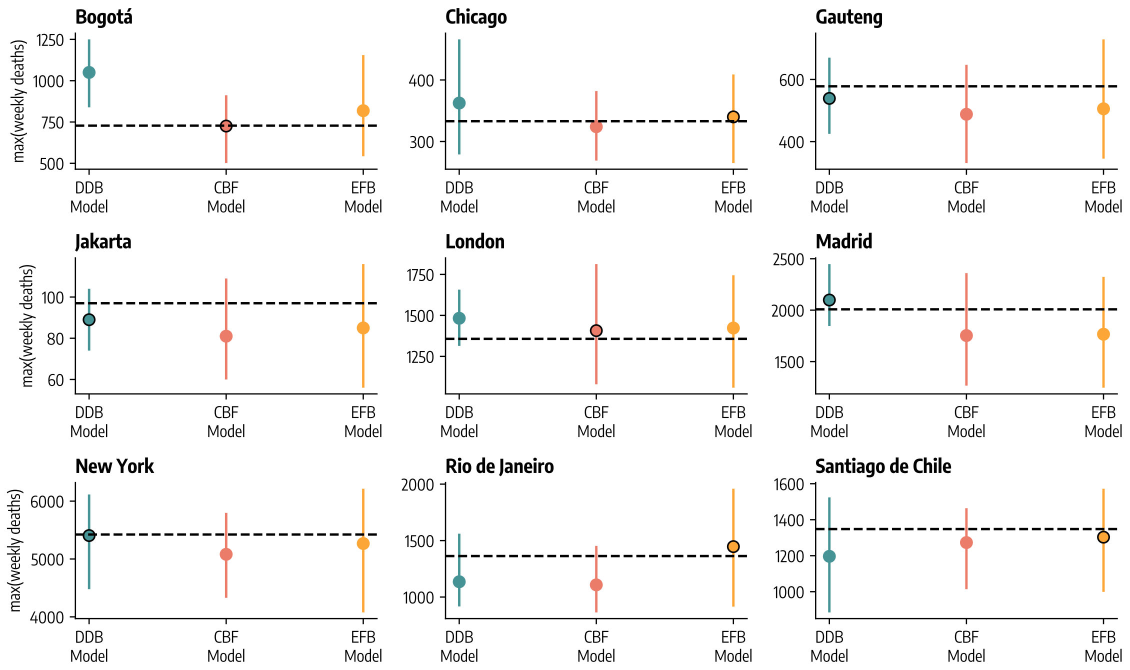

Figure 1: Median and 90% predictive intervals of weekly death peak intensity, estimated from 1,000 stochastic trajectories across three epidemic-behavior models. The horizontal dashed lines represent the observed peak intensity in reported weekly deaths. The model with the median peak intensity closest to the observed data is marked with a dot outlined in black. As described in the text, DDB stands for Data-Driven Behavioral model, CBF for Compartmental Behavioral Feedback model, and EFB for Effective Force of Infection Behavioral Feedback model.

### 4 $R_0$ posterior distributions

In Fig. 3A we show the posterior distributions for the basic reproductive number $R_0$ estimated via ABC-SMC calibration for the three models across the nine geographies considered during the retrospective analysis. Additionally, we report median and 90% predictive intervals of these posterior distributions in Tab. 3

In Fig. 3B we show the Wasserstein distance among $R_0$ posterior distributions estimated by each pair of models. We display, the average distance across the nine geographies, for a given pair of models. Overall, we observe that the posterior distributions derived from the Compartmental and the Effective Force of Infection Behavioral Feedback models exhibit greater similarity to each other (i.e., lower distance) compared to those estimated by the Data-Driven Behavioral model utilizing mobility data.

### 5 Forecasting performance: Mean Absolute Error

Here, we report the forecasting performance results considering the Mean Absolute Error (MAE) of the median instead of the Weighted Interval Score as the evaluation metric. The findings are consistent with the analysis presented in the main text. We use the Wilcoxon signed-rank test to statistically compare the performance of different models. The null hypothesis of this test is that the two groups come from

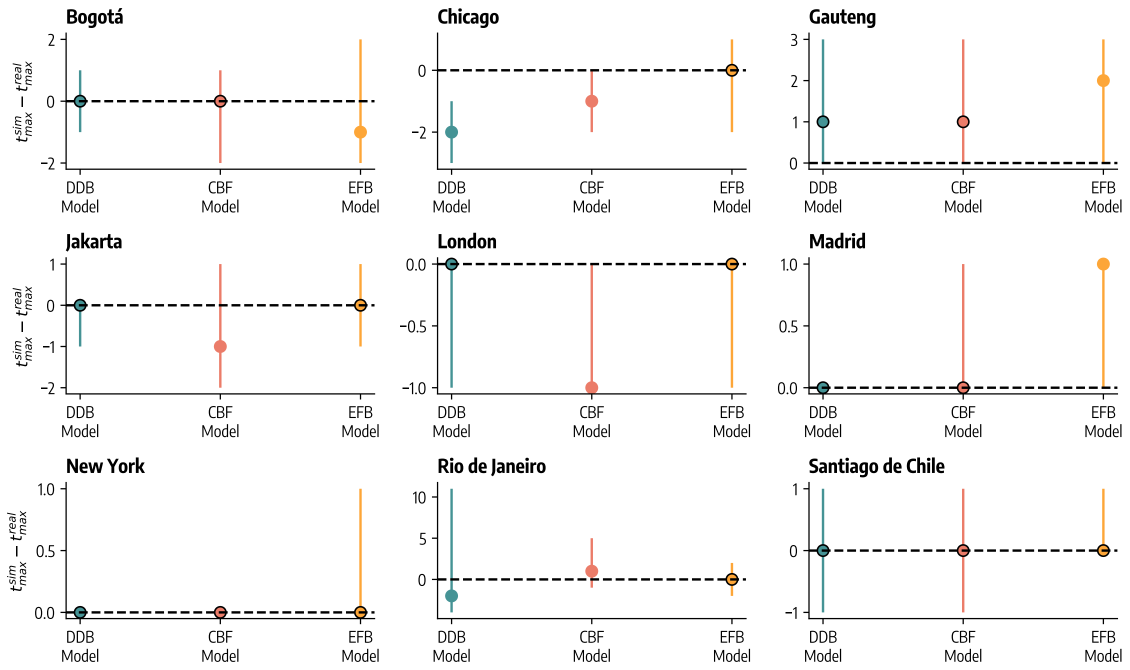

Figure 2: Median and 90% predictive intervals of the difference between simulated and reported weekly death peak timing, based on 1,000 stochastic trajectories from three epidemic-behavior models. The horizontal dashed line at 0 indicates perfect alignment with the observed peak timing. Models with peak timing closest to the reported data are highlighted with a dot outlined in black. As detailed in the text, DDB refers to the Data-Driven Behavioral model, CBF to the Compartmental Behavioral Feedback model, and EFB to the Effective Force of Infection Behavioral Feedback model.

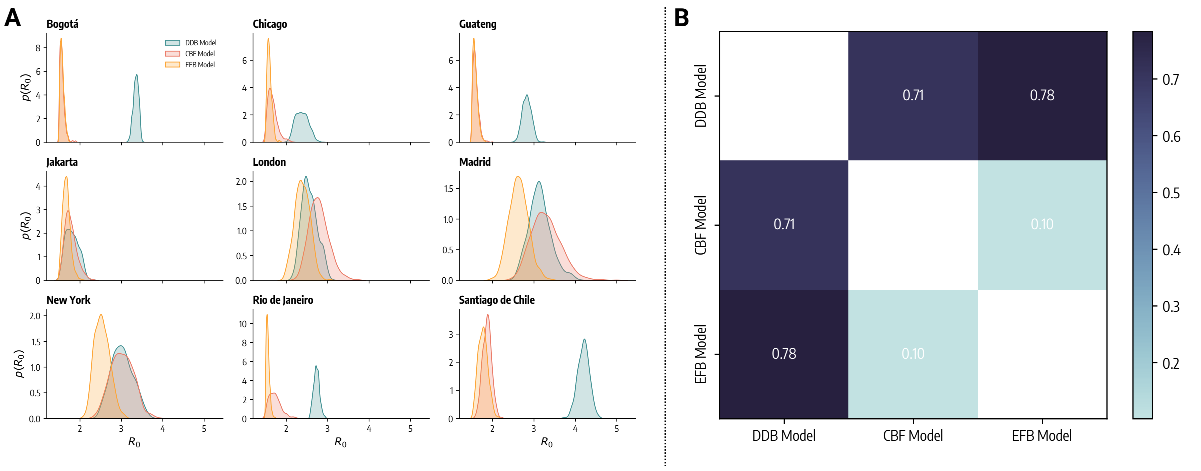

Figure 3: A) Posterior distributions for $R_0$ estimated via Approximate Bayesian Computation calibration for the three models across the nine geographies considered (1,000 stochastic posterior samples). B) Pairwise Wasserstein distance among $R_0$ posterior distributions estimated by different models. For a given pair of models, the average distance across the nine geographies is displayed.

the same distribution. In Fig. 4 we report the statistical significance of the tests comparing different pairs of models as follows: ****: $p_{value} \leq 10^{-4}$, ***: $10^{-4} < p_{value} \leq 10^{-3}$, **: $10^{-3} < p_{value} \leq 10^{-2}$, *: $10^{-2} < p_{value} \leq 0.05$, and otherwise blank if $p_{value} > 0.05$

From Fig. 4A we observe heterogeneous forecasting performance among the geographies under consid-

Table 3: Posterior distributions (median and 90% confidence intervals from $1,000$ posterior samples) for basic reproductive number $R_0$ estimated by the three models across nine geographies considered.

| **Region** | **DDB Model** | **CBF Model** | **CBF Model** |
|---|---|---|---|
| Bogotá | 3.36 [3.24, 3.45] | 1.56 [1.51, 1.67] | 1.56 [1.51, 1.66] |
| Chicago | 2.37 [2.15, 2.63] | 1.64 [1.52, 1.89] | 1.58 [1.52, 1.70] |
| Gauteng | 2.83 [2.65, 3.01] | 1.58 [1.51, 1.71] | 1.57 [1.51, 1.69] |
| Jakarta | 1.80 [1.58, 2.08] | 1.75 [1.58, 2.05] | 1.67 [1.54, 1.84] |
| London | 2.54 [2.29, 2.90] | 2.77 [2.43, 3.20] | 2.38 [2.09, 2.68] |
| Madrid | 3.13 [2.73, 3.63] | 3.27 [2.76, 3.91] | 2.62 [2.26, 3.01] |
| New York | 3.00 [2.60, 3.46] | 3.01 [2.57, 3.46] | 2.52 [2.26, 2.85] |
| Rio de Janeiro | 2.73 [2.62, 2.85] | 1.71 [1.53, 2.01] | 1.55 [1.50, 1.63] |
| Santiago de Chile | 4.20 [3.96, 4.42] | 1.87 [1.69, 2.04] | 1.77 [1.59, 1.97] |

eration. Notably, in London, Madrid, and New York all models exhibit a median relative MAE smaller than 1, indicating better performance compared to the baseline. On the other hand, we note lower performance in other cases such as the Data-Driven Behavioral model in Bogotá and all three models, to different extents, in Rio de Janeiro, which generally have worse performance with respect to the baseline. In terms of median MAE, the Data-Driven model outperforms the others in 5 locations, with a statistically significant difference from the second-best model in only 2 of those locations. The Compartmental Behavioral model outperforms the others in the remaining 4 locations, with a statistically significant difference from the Data-Driven model in 3 of them.

Similar to what we found for the WIS, in Fig. 4B we see that, among the three epidemic-behavior models considered in this study, the Compartmental Behavioral Feedback model generally emerges as the top performer, followed by the Data-Driven and the Effective Force of Infection Behavioral Feedback model.

#### 6 Forecasting performance by Horizon

We extend the forecasting performance analysis by dividing the forecasts by horizon, ranging from 1 to 4 weeks ahead. In Fig. 5A, we present the relative WIS by horizon for the three epidemic-behavior models across the nine locations. In Fig. 5B, we show the overall relative WIS by horizon for the three models, pooling all locations together.

Generally, we observe that forecasting performance improves with longer horizons (i.e., the relative WIS decreases for higher horizons). While this may seem counterintuitive at first, it is important to note that this performance is relative to a baseline model. Although absolute performance may decline over longer horizons, relative performance can still improve. However, as seen in Fig. 5A, this pattern does not hold uniformly across all location/model combinations.

Regarding model comparison, Fig. 5B shows that the Compartmental Behavioral Feedback model consistently outperforms the others across all forecasting horizons, confirming the findings from the

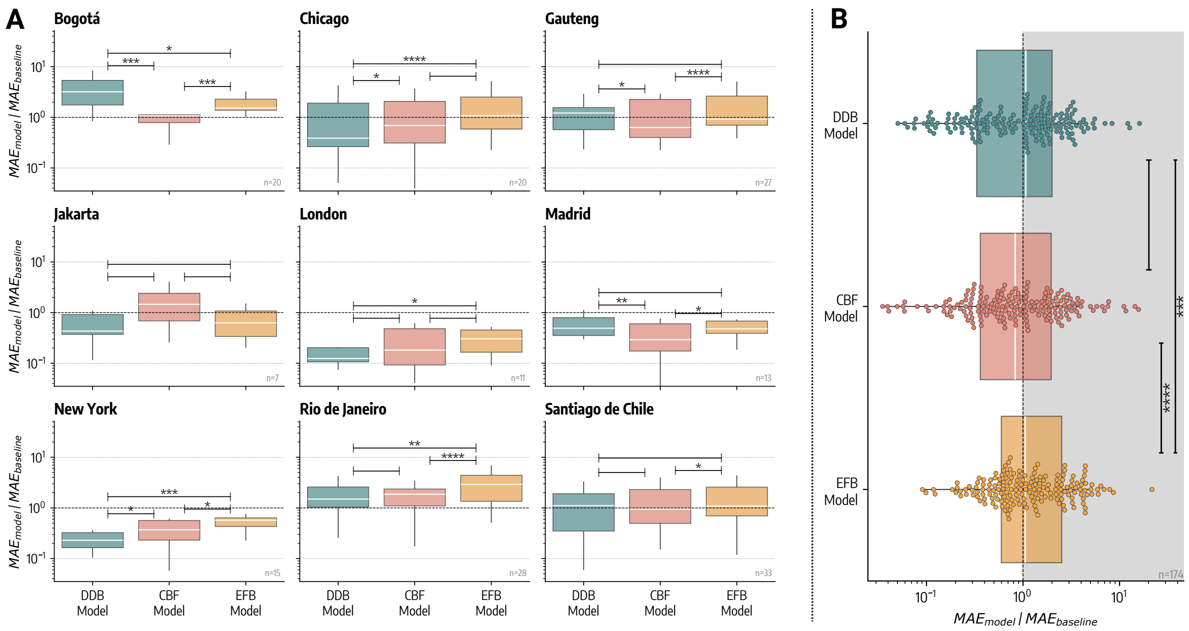

Figure 4: Forecasting performance (MAE). A) Relative MAE computed over all forecasting rounds for the three epidemic-behavior models across the nine geographical regions considered. Values below 1 indicate better performance with respect to baseline forecasting model. Each data point underlying the boxplot represents the relative MAE over the four-week horizon of the corresponding forecasting round. In the bottom right of each plot, we report the number of forecasting rounds for each location. B) Boxplot and swarmplot of relative MAE for different models pooling together results from all rounds and geographies. The box boundaries represent the interquartile range (IQR) between the first and third quartiles (Q1 and Q3), the line inside the box indicates the median and the upper (lower) whisker extends to the last datum less (greater) than $Q3 + 1.5IQR$ ($Q1 - 1.5IQR$). DDB stands for Data-Driven Behavioral model, CBF for Compartmental Behavioral Feedback model, and EFB for Effective Force of Infection Behavioral Feedback model. In both panels we report the statistical significance of the Wilcoxon test comparing different forecasting performances as follows: ****: $p_{value} \leq 10^{-4}$, ***: $10^{-4} < p_{value} \leq 10^{-3}$, **: $10^{-3} < p_{value} \leq 10^{-2}$, *: $10^{-2} < p_{value} \leq 0.05$, and otherwise blank if $p_{value} > 0.05$.

main text.

In Fig. 6, we repeat the analysis using the Absolute Error (AE) as the evaluation metric by horizon, finding similar overall results.

### 7 Forecasting performance of ensemble models

We consider two ensemble models that combine the forecasts of each individual epidemic-behavior model. In several epidemiological forecasting contexts, ensemble forecasts have consistently demonstrated greater accuracy and reliability over time compared to individual models [32, 33]. In both approaches discussed here, the prediction intervals of the ensemble are calculated as a weighted average of the intervals from individual models. That is, the $n^{th}$ quantile of the ensemble is defined as: $q_n^{ens.} = \sum_m w_m q_n^m$, where $q_n^m$ is the $n^{th}$ quantile of individual model $m$ and $w_m$ is its weight. In the first approach, referred to as the simple ensemble, we assign $w_m^{simple} = 1/3$ to each of the three epidemic-behavior models $m$, meaning

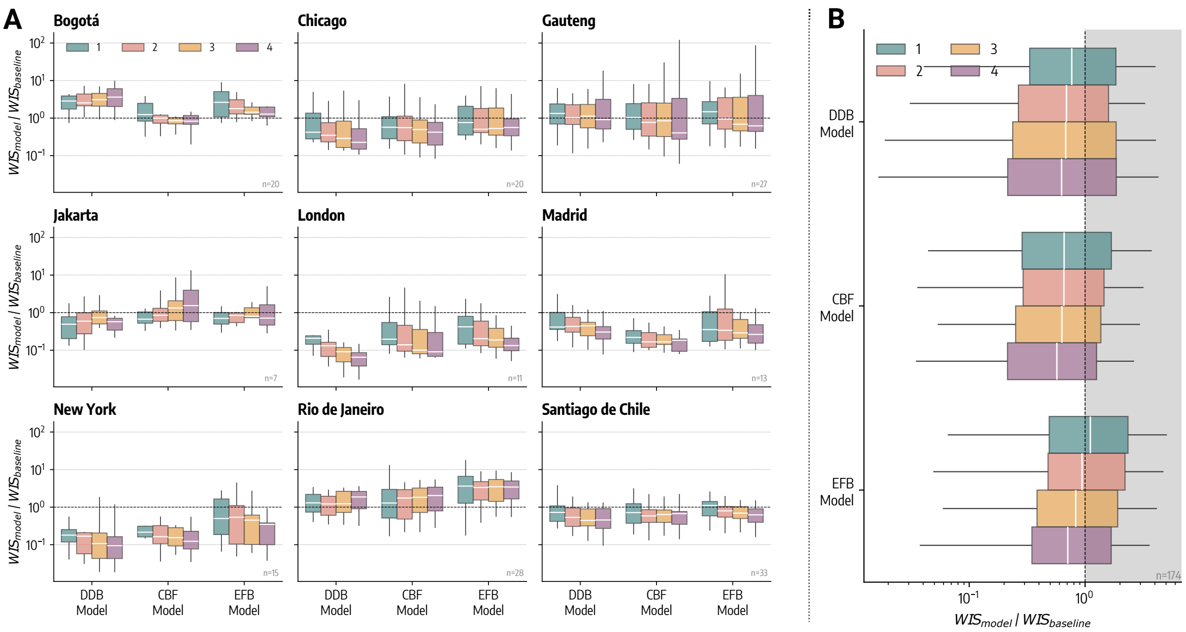

Figure 5: Forecasting performance (WIS) by horizon. Relative WIS computed over all forecasting rounds for the three epidemic-behavior models across the nine geographical regions considered divided by horizon (1 to 4 weeks ahead). Values below 1 indicate better performance with respect to the baseline forecasting model. Each data point underlying the boxplot represents the relative WIS for a given horizon of the corresponding forecasting round. B) Boxplot of relative WIS for different models pooling together results from all rounds and geographies divided by horizon (1 to 4 weeks ahead). The box boundaries represent the interquartile range (IQR) between the first and third quartiles (Q1 and Q3), the line inside the box indicates the median and the upper (lower) whisker extends to the last datum less (greater) than $Q3 + 1.5IQR$ ($Q1 - 1.5IQR$). As described in the text, DDB stands for Data-Driven Behavioral model, CBF for Compartmental Behavioral Feedback model, and EFB for Effective Force of Infection Behavioral Feedback model.

all models are equally weighted. Consequently, the weighted average simplifies to a basic arithmetic mean. In the second approach, referred to as the weighted ensemble, models are weighted based on their past forecasting performance. Specifically, the weight assigned to model $m$ at time $t$ is the inverse of its average forecasting performance $\gamma$, computed over the last three forecasting rounds. This means that the weight for model $m$ is computed as $w_m^{weighted} = \left(\frac{\sum_{k=1}^{3} \gamma_{t-k}^m}{3}\right)^{-1}$. Forecasting performance is assessed using either the WIS or the MAE of the median, depending on the metric used to evaluate the weighted ensemble. In Fig. 7 and Fig. 8, we present the weights of the epidemic-behavior models used to construct the weighted ensemble across different forecasting rounds, with the past performance of individual models evaluated using either the WIS or the MAE.

In Fig. 9A, we present the ratio, for all forecasting rounds, between the average WIS (over the 4-week horizon) of each model epidemic-behavior and the two ensembles and the average WIS of a baseline model. Across the board, we observe that the weighted ensemble consistently outperforms the baseline at all locations in terms of the median (i.e., median relative WIS is less than 1). The simple ensemble generally performs better than the baseline at all locations, except for Rio de Janeiro, where

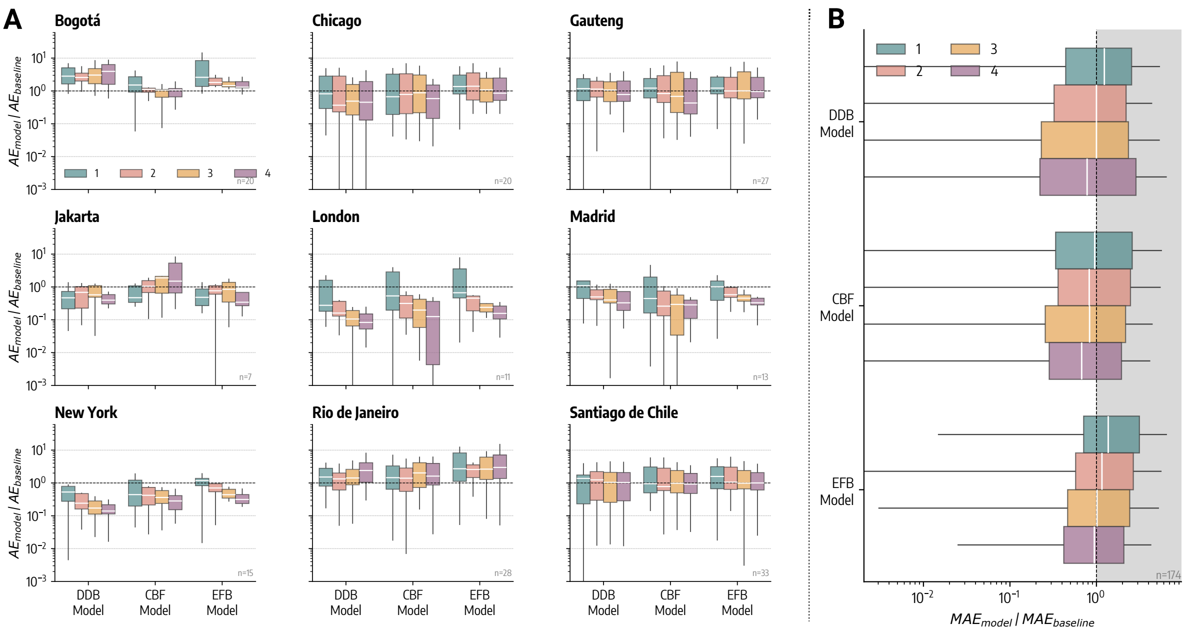

Figure 6: Forecasting performance (AE) by horizon. Relative AE computed over all forecasting rounds for the three epidemic-behavior models across the nine geographical regions considered divided by horizon (1 to 4 weeks ahead). Values below 1 indicate better performance with respect to the baseline forecasting model. Each data point underlying the boxplot represents the relative AE for a given horizon of the corresponding forecasting round. B) Boxplot of relative AE for different models pooling together results from all rounds and geographies divided by horizon (1 to 4 weeks ahead). The box boundaries represent the interquartile range (IQR) between the first and third quartiles (Q1 and Q3), the line inside the box indicates the median and the upper (lower) whisker extends to the last datum less (greater) than $Q3 + 1.5IQR$ ($Q1 - 1.5IQR$). As described in the text, DDB stands for Data-Driven Behavioral model, CBF for Compartmental Behavioral Feedback model, and EFB for Effective Force of Infection Behavioral Feedback model.

its relative WIS median is slightly greater than 1. The weighted ensemble achieves the lowest median relative WIS in Gauteng, Rio de Janeiro, and Santiago de Chile, while the simple ensemble only in Bogotá. In Fig. 9B we show the distribution of the relative WIS with respect to the baseline model for the three epidemic-behavior models and the two ensembles, combining results from all geographies and all forecasting points to provide an overall view of models' performance. Interestingly, both ensembles achieve an overall relative WIS median significantly lower than that of the individual models, specifically 0.45 for the simple ensemble and 0.44 for the weighted ensemble. The performance distributions of the two ensembles are also statistically different ($p_{val} < 0.05$) according to the Wilcoxon signed-rank test. For the weighted ensemble, nearly 80% of forecasts outperform the baseline, compared to around 75% for the simple ensemble.

In Fig. 10, we repeat the analysis of ensemble performance using the MAE of the median as the performance metric. The findings obtained with the WIS are confirmed. The weighted ensemble outperforms the baseline in median terms at all locations, while the simple ensemble does so at all locations except Rio de Janeiro. The weighted ensemble achieves the lowest relative MAE median in three locations

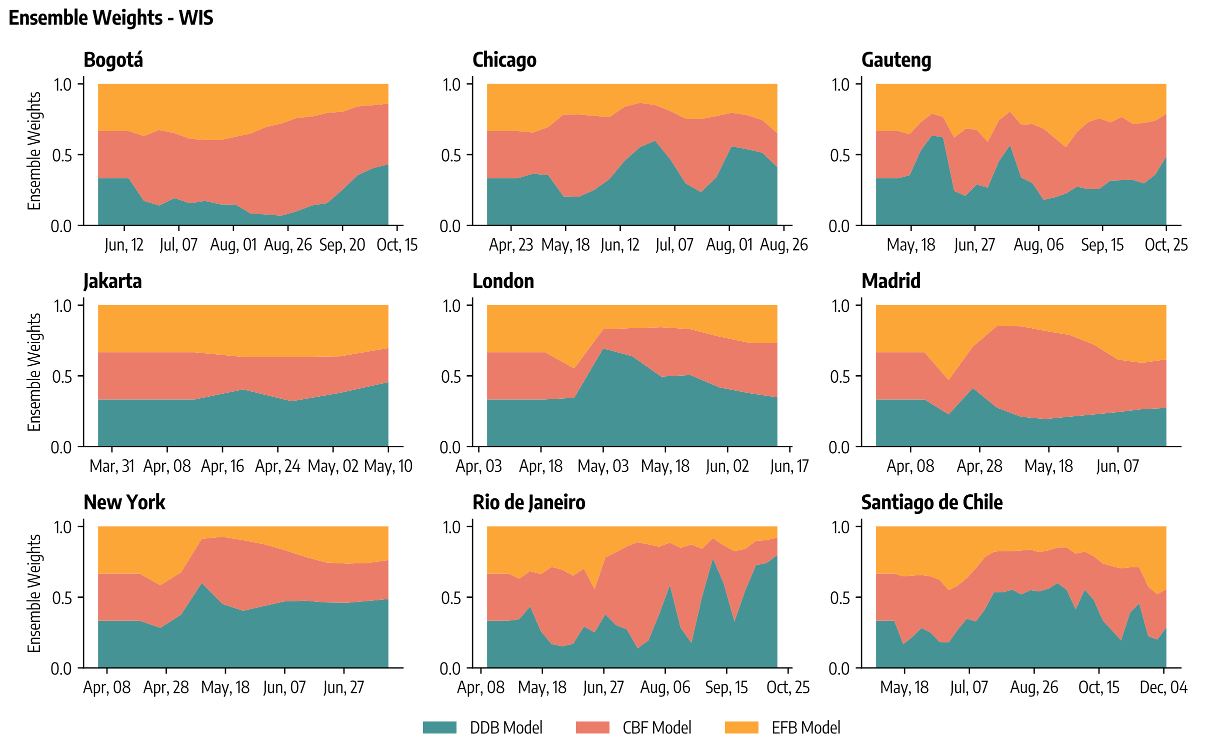

Figure 7: Weights of the epidemic-behavior models used to construct the weighted ensemble across different forecasting rounds, based on the WIS to evaluate the past performance of individual models.

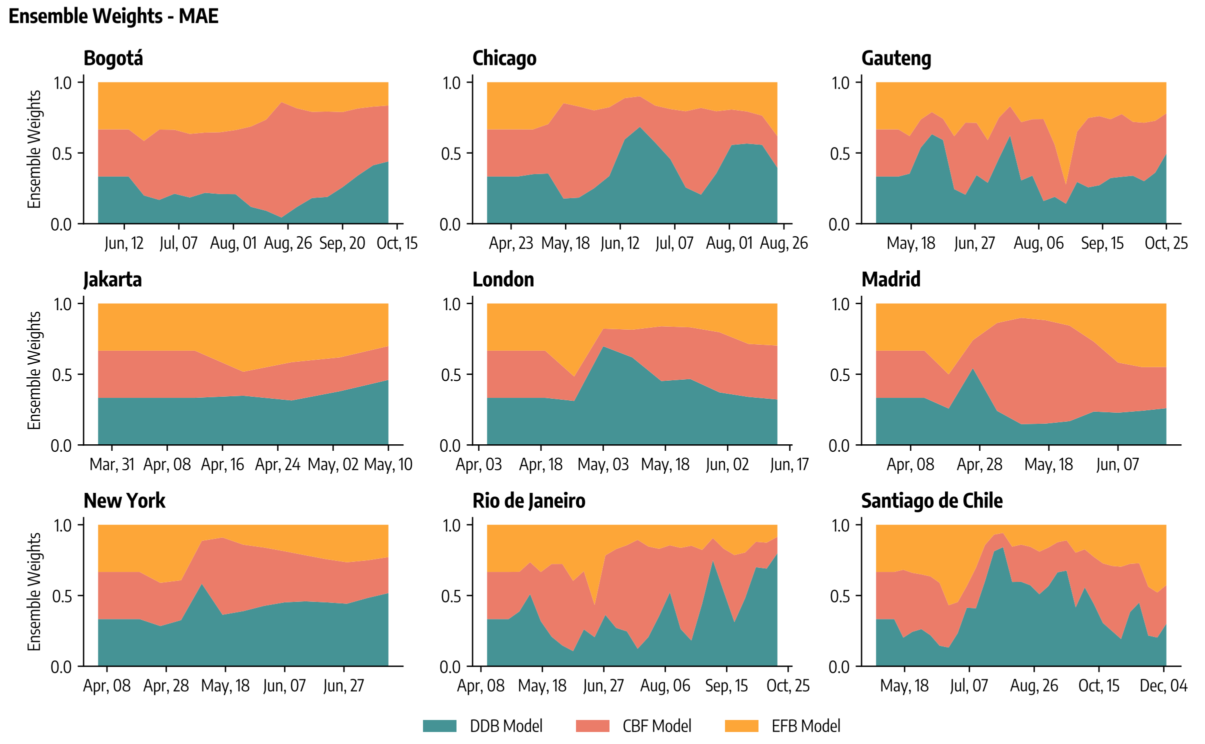

Figure 8: Weights of the epidemic-behavior models used to construct the weighted ensemble across different forecasting rounds, based on the MAE to evaluate the past performance of individual models.

(Bogotá, Rio de Janeiro, and Santiago de Chile), while the simple ensemble does so in two (Gauteng and London). When examining overall model performance across all locations in Fig. 10B, both the weighted and simple ensembles achieve a median relative MAE of 0.57. In this case, their performance distributions are not statistically different, according to the Wilcoxon signed-rank test ($p_{val} > 0.05$).

Finally, in Tab. 4, we present the differences in forecasting performance between the ensemble and individual forecasting models. Specifically, we report the percentage difference between the median ensemble model performance and that of each individual model, both in terms of WIS and MAE. Negative percentages indicate that the ensemble improves performance (i.e., achieves a lower median WIS or MAE), while positive values indicate poorer performance relative to the individual model (i.e., achieves a higher median WIS or MAE). The table also includes the statistical significance of the differences in forecasting performance, as measured by the Wilcoxon signed-rank test. For the WIS, both the simple and weighted ensembles lead to improved forecasting performance in 78% of cases (location/model combinations). When considering the MAE, the simple ensemble improves forecasting performance in 81% of cases, while the weighted ensemble does so in 74%. When examining the overall difference between the ensemble and individual models, we find that both the simple and weighted ensembles significantly outperform each individual model across both metrics. However, the improvements achieved by the weighted ensemble are very similar to those of the simple ensemble.

### 8 Posterior distributions in time

We report the posterior distributions of key behavioral parameters obtained in different forecasting rounds. More detail, Fig. 11, 12, 13, 14 show posterior distribution of behavioral parameters ($r$, $\beta_B$, $\mu_B$, $\gamma$) for the Compartmental Behavioral Feedback model, and 15, 16 for the Effective Force of Infection Behavioral Feedback model ($\xi$, $\psi$). The Data-Driven Behavioral model does not have any additional free parameters beyond the ones common to all models. In each figure, we also report the posterior distribution (median and 95% predictive intervals) obtained in the retrospective modeling exercise as a horizontal grey line and shaded area. We observe that posterior distributions resulting from forecasting are similar to those resulting from retrospective modeling, especially for the last forecasting rounds, where models were trained on more data.

To further analyze the evolution and stability of the estimated posterior distributions over time, we track the cosine similarity of the median posterior distribution. For two forecasting rounds, $t_1$ and $t_2$, this similarity is defined as:

$$S_C(t_1, t_2) = \frac{\hat{\boldsymbol{\theta}}(t_1) \cdot \hat{\boldsymbol{\theta}}(t_2)}{||\hat{\boldsymbol{\theta}}(t_1)|| \; ||\hat{\boldsymbol{\theta}}(t_2)||} \tag{9}$$

where $\hat{\boldsymbol{\theta}}(t)$ is a vector representing the median posterior distribution of each free parameter estimated

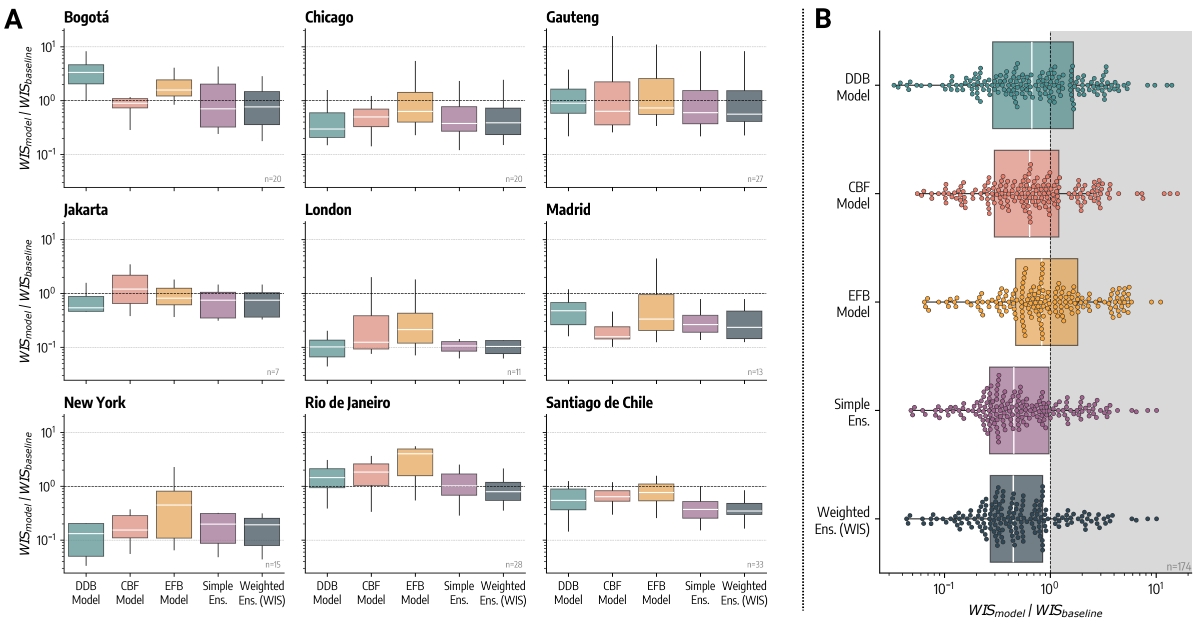

Figure 9: Forecasting performance (WIS) of ensemble and individual epidemic-behavior models. Relative WIS computed over all forecasting rounds for the three epidemic-behavior models and the two ensemble models across the nine geographical regions considered. Values below 1 indicate better performance with respect to baseline forecasting model. Each data point underlying the boxplot represents the relative WIS over the four-week horizon of the corresponding forecasting round. In the bottom right of each plot, we report the number of forecasting rounds for each location. B) Boxplot and swarmplot of relative WIS for different epidemic-behavior and ensemble models pooling together results from all rounds and geographies. The box boundaries represent the interquartile range (IQR) between the first and third quartiles (Q1 and Q3), the line inside the box indicates the median and the upper (lower) whisker extends to the last datum less (greater) than $Q3+1.5IQR$ ($Q1-1.5IQR$). DDB stands for Data-Driven Behavioral model, CBF for Compartmental Behavioral Feedback model, and EFB for Effective Force of Infection Behavioral Feedback model.

during forecasting round $t$. To prevent the scale of some parameters from dominating the similarity, each parameter is normalized to its maximum value.

In Fig. 17, we show the evolution of $S_C(t, t-1)$, which represents the cosine similarity between the median posterior distributions estimated in round $t$ and the previous round. Across all locations, the posterior distributions in successive steps are highly similar, with the lowest values exceeding 0.9, observed mainly at the beginning of the iterative forecasting process when less data is available for calibration.

In Fig. 18, we show the evolution of $S_C(t, t_0)$, representing the cosine similarity between the posterior distributions estimated in round $t$ and the initial round. Here, we observe a decrease in similarity early on, although the values never fall below 0.7. This indicates that the posterior distributions remain stable even with respect to the initial estimates, despite the limited data available initially.

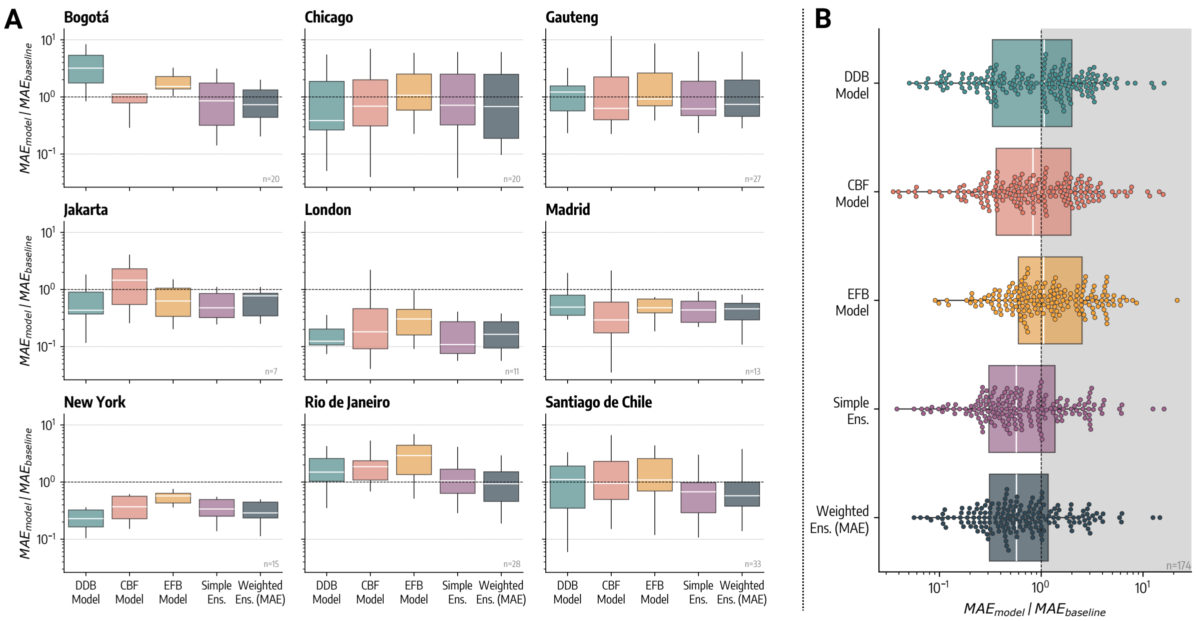

Figure 10: Forecasting performance (MAE) of ensemble and individual epidemic-behavior models. Relative MAE computed over all forecasting rounds for the three epidemic-behavior models and the two ensemble models across the nine geographical regions considered. Values below 1 indicate better performance with respect to baseline forecasting model. Each data point underlying the boxplot represents the relative MAE over the four-week horizon of the corresponding forecasting round. In the bottom right of each plot, we report the number of forecasting rounds for each location. B) Boxplot and swarmplot of relative MAE for different epidemic-behavior and ensemble models pooling together results from all rounds and geographies. The box boundaries represent the interquartile range (IQR) between the first and third quartiles (Q1 and Q3), the line inside the box indicates the median and the upper (lower) whisker extends to the last datum less (greater) than $Q3 + 1.5IQR$ $(Q1 - 1.5IQR)$. DDB stands for Data-Driven Behavioral model, CBF for Compartmental Behavioral Feedback model, and EFB for Effective Force of Infection Behavioral Feedback model.

### 9 N-week ahead forecasts

Here, we show one to four week ahead forecasts for the DDB model (Fig. 19, 25, 31, 37), the CBF model (Fig. 20, 26, 32, 38), the EFB model (Fig. 21, 27, 33, 39), the simple ensemble (Fig. 22, 28, 34, 40), the ensemble weighted according to past WIS performance (Fig. 23. 29, 35, 41), and the ensemble weighted according to past MAE performance in the nine locations (Fig. 24. 30, 36, 42).

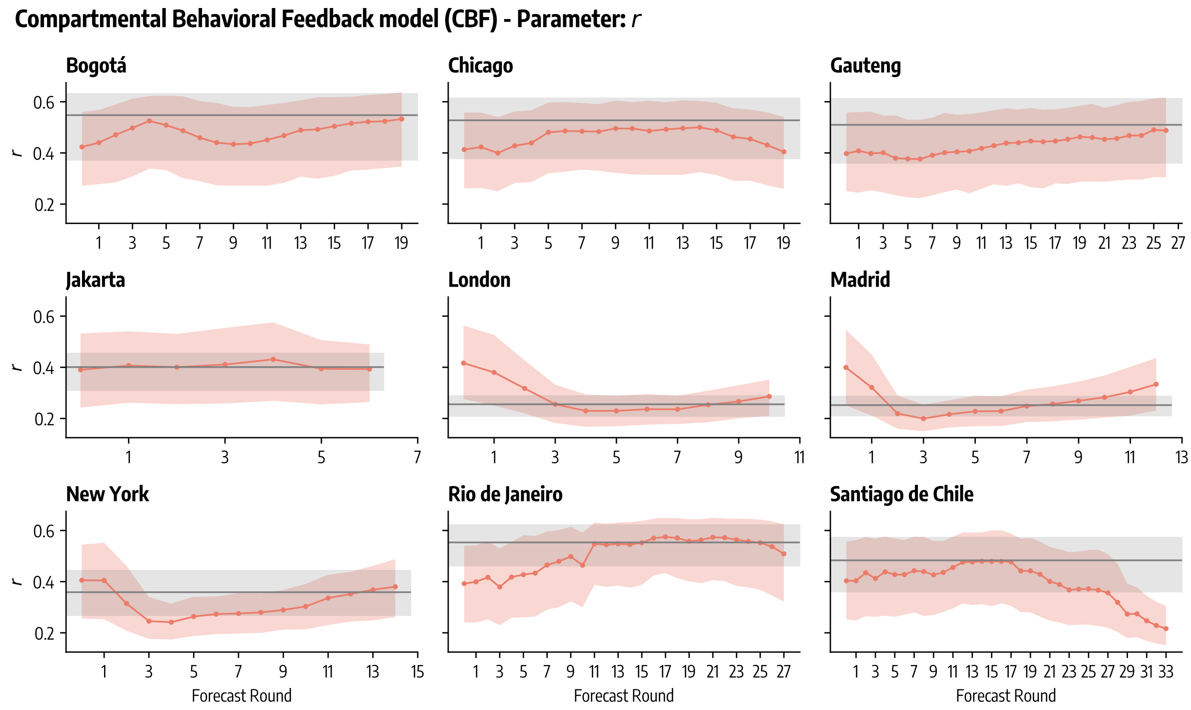

Figure 11: Posterior distributions in time. Compartmental Behavioral Feedback model - $r$. Median and 95% predictive intervals in different forecasting rounds obtained from $1,000$ posterior samples. Posterior distribution (median and 95% predictive intervals) obtained from retrospective modeling are also reported in grey.

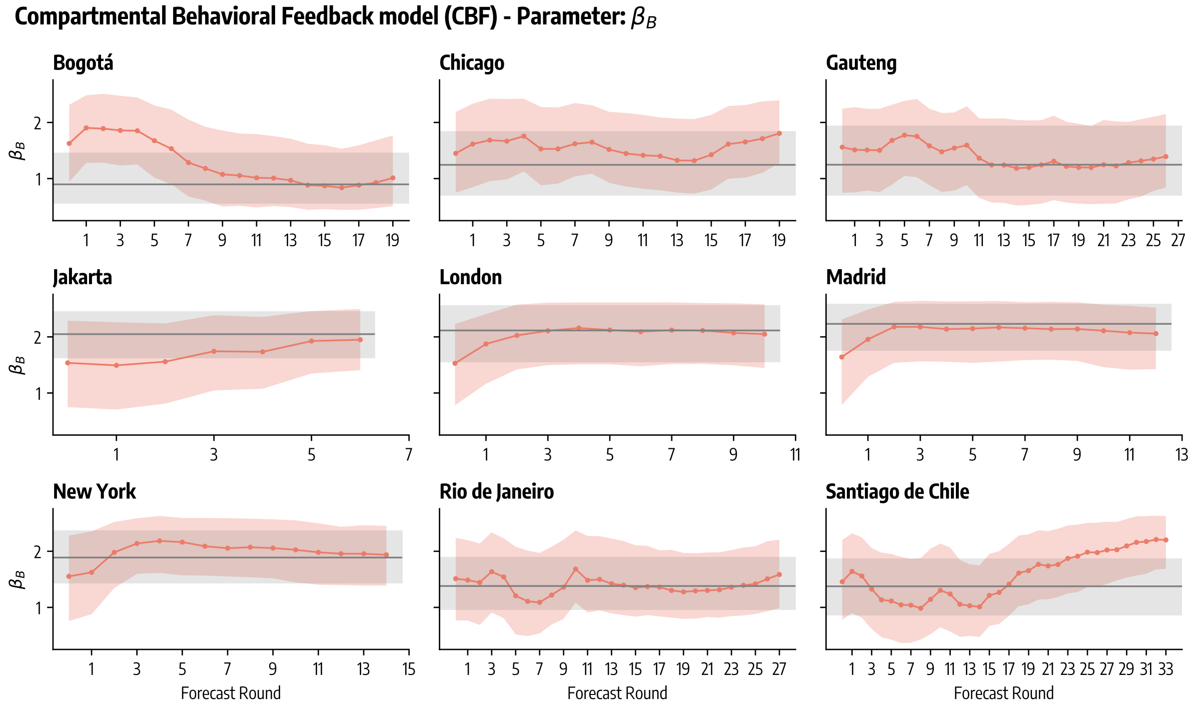

Figure 12: Posterior distributions in time. Compartmental Behavioral Feedback model - $\beta_B$. Median and 95% predictive intervals in different forecasting rounds obtained from $1,000$ posterior samples. Posterior distribution (median and 95% predictive intervals) obtained from retrospective modeling are also reported in grey.

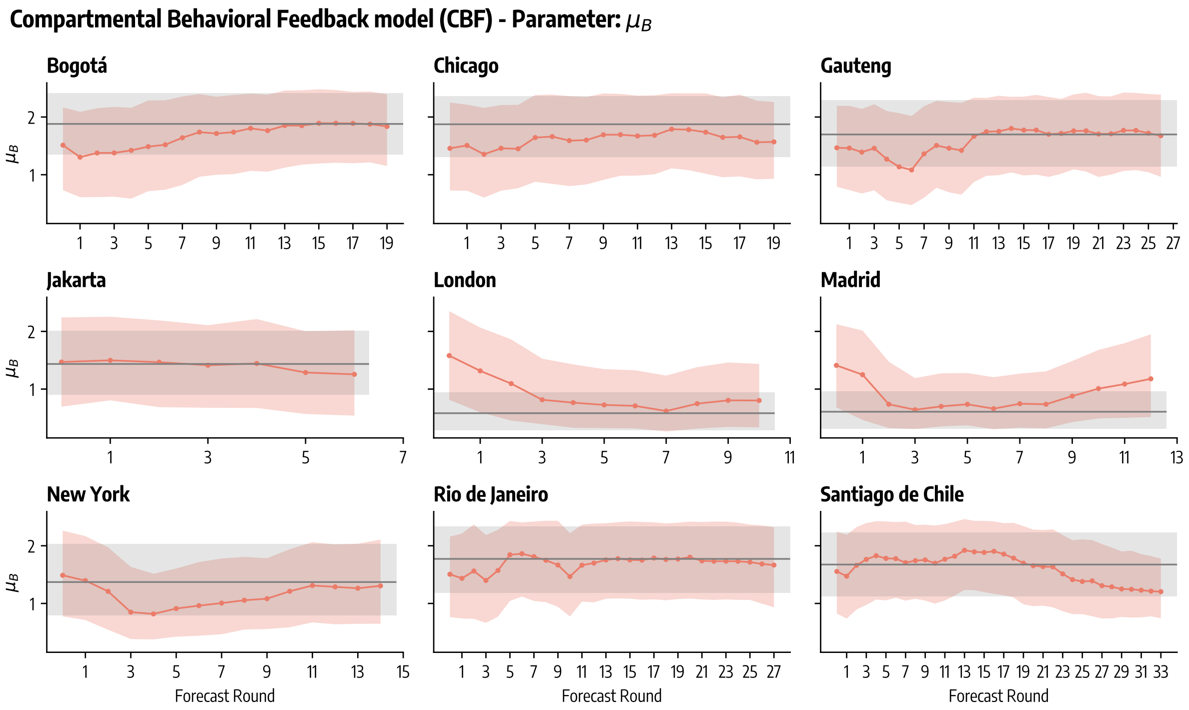

Figure 13: Posterior distributions in time. Compartmental Behavioral Feedback model - $\mu_B$. Median and 95% predictive intervals in different forecasting rounds obtained from 1,000 posterior samples. Posterior distribution (median and 95% predictive intervals) obtained from retrospective modeling are also reported in grey.

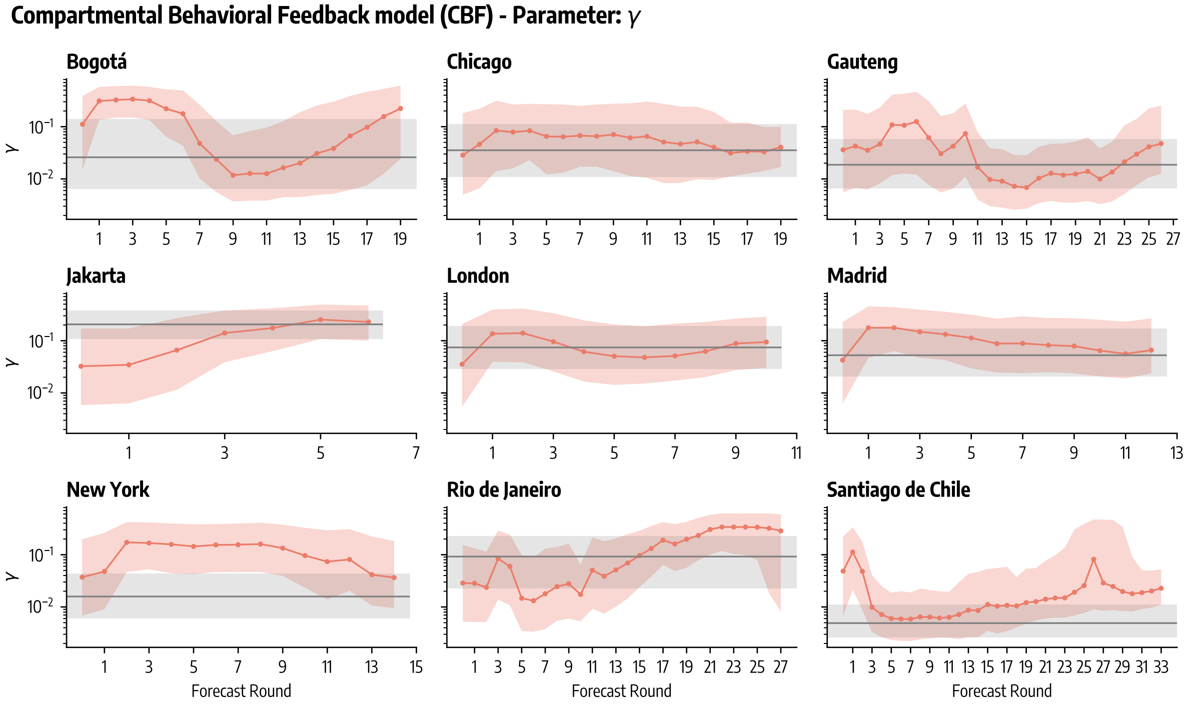

Figure 14: Posterior distributions in time. Compartmental Behavioral Feedback model - $\gamma$. Median and 95% predictive intervals in different forecasting rounds obtained from 1,000 posterior samples. Posterior distribution (median and 95% predictive intervals) obtained from retrospective modeling are also reported in grey.

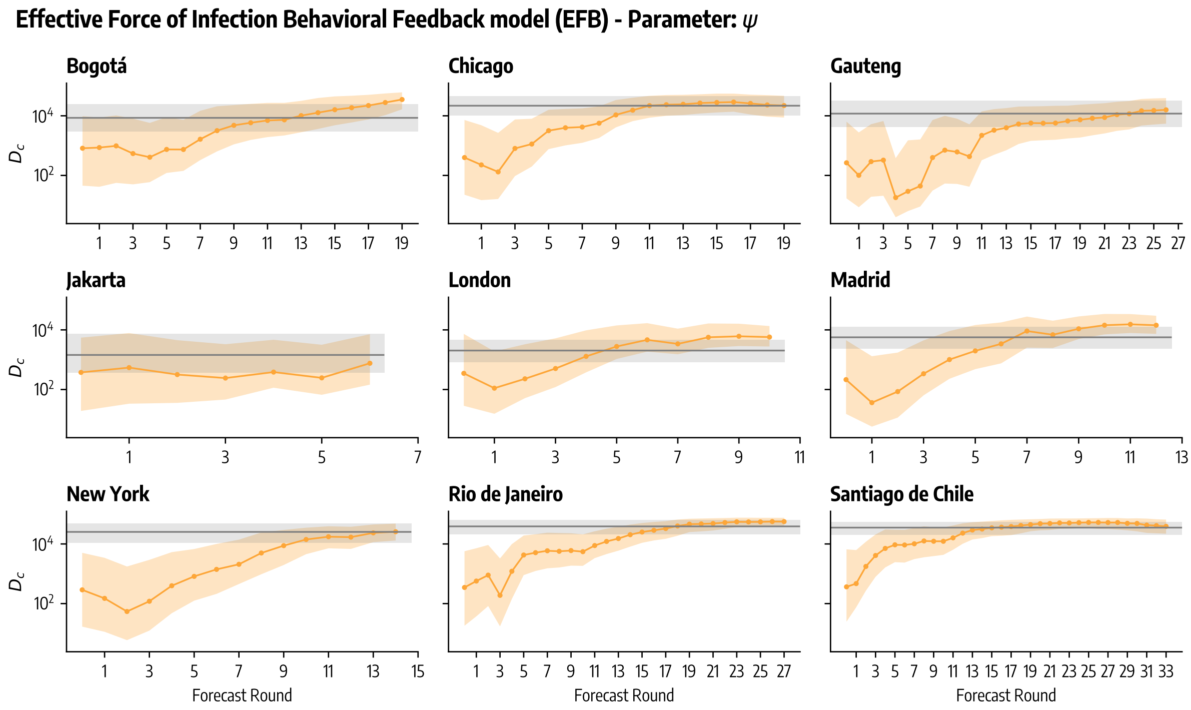

Figure 15: Posterior distributions in time. Effective Force of Infection Behavioral Feedback model - $\psi$. Median and 95% predictive intervals in different forecasting rounds obtained from 1,000 posterior samples. Posterior distribution (median and 95% predictive intervals) obtained from retrospective modeling are also reported in grey.

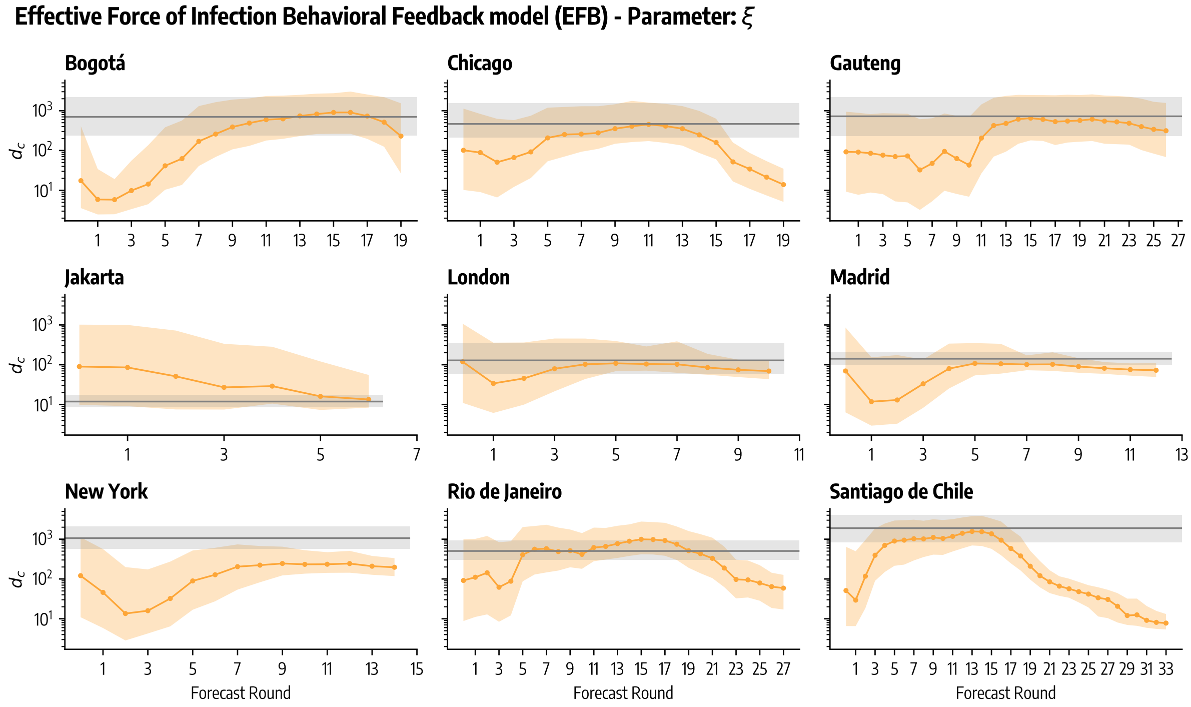

Figure 16: Posterior distributions in time. Effective Force of Infection Behavioral Feedback model - $\xi$. Median and 95% predictive intervals in different forecasting rounds obtained from 1,000 posterior samples. Posterior distribution (median and 95% predictive intervals) obtained from retrospective modeling are also reported in grey.

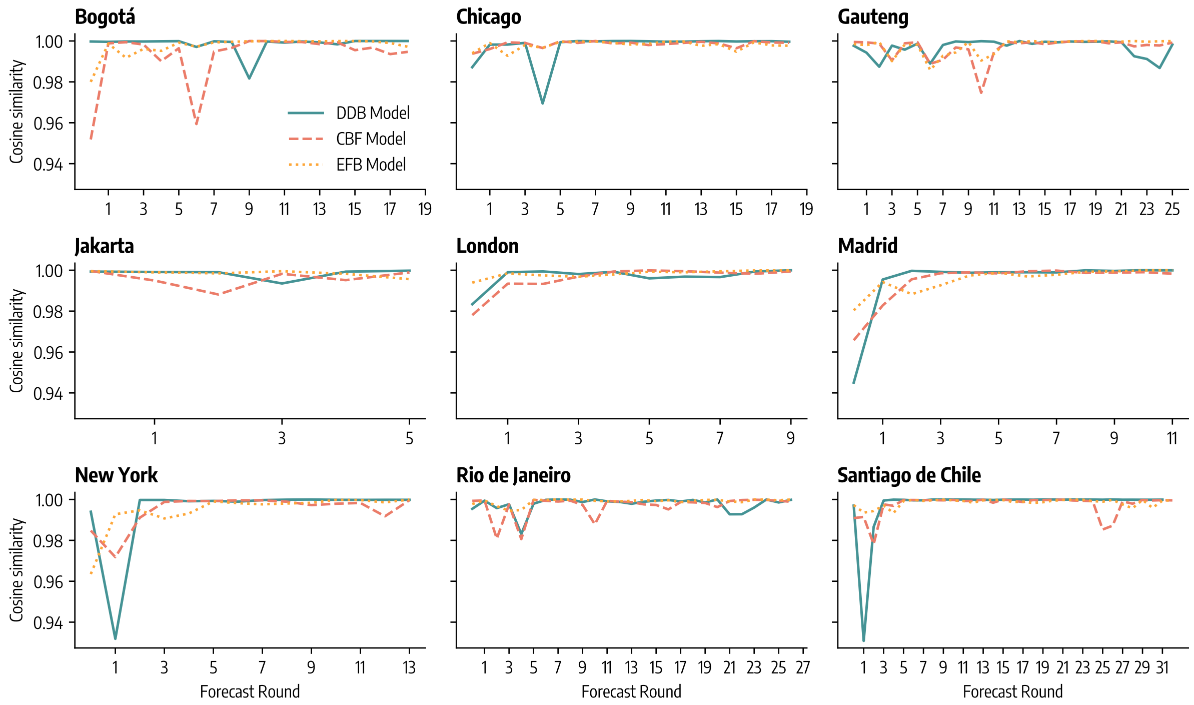

Figure 17: Cosine similarity between median posterior distributions estimated at forecasting round $t$ with respect to previous forecasting round $t-1$. DDB stands for Data-Driven Behavioral model, CBF for Compartmental Behavioral Feedback model, and EFB for Effective Force of Infection Behavioral Feedback model.

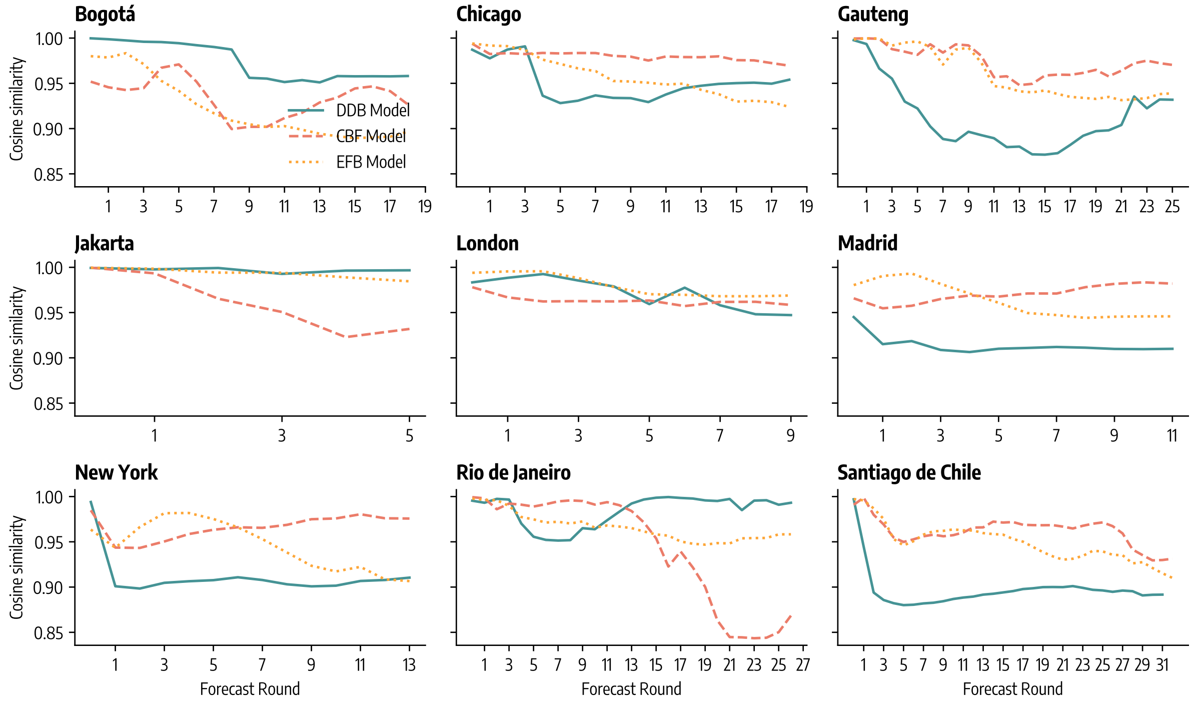

Figure 18: Cosine similarity between median posterior distributions estimated at forecasting round $t$ with respect to first forecasting round $t_0$. DDB stands for Data-Driven Behavioral model, CBF for Compartmental Behavioral Feedback model, and EFB for Effective Force of Infection Behavioral Feedback model.

| | | Δ% MAE | | | Δ% WIS | | |
|---|---|---|---|---|---|---|---|
| | | DDB | CBF | EFB | DDB | CBF | EFB |
| *Bogotá* | Simple Ensemble | -73.2% (**) | -20.4% () | -43.9% (****) | -78.9% (***) | -21.3% () | -55.2% (****) |
| | Weighted Ensemble | -77.1% (**) | -31.9% () | -52.0% (****) | -77.1% (***) | -14.5% () | -51.4% (****) |
| *Chicago* | Simple Ensemble | 85.2% () | 4.2% (**) | -32.7% (**) | 26.8% () | -24.9% (**) | -40.2% (**) |
| | Weighted Ensemble | 77.2% () | -0.2% (***) | -35.6% (**) | 31.3% () | -22.3% (**) | -38.0% (**) |
| *Gauteng* | Simple Ensemble | -49.2% () | -1.8% () | -32.8% (****) | -33.2% (***) | -5.6% () | -18.2% (***) |
| | Weighted Ensemble | -39.1% () | 17.6% () | -19.5% (***) | -37.4% (***) | -11.5% () | -23.3% (**) |
| *Jakarta* | Simple Ensemble | 10.5% () | -67.2% () | -23.9% () | 38.4% () | -37.8% (*) | -8.3% () |
| | Weighted Ensemble | 79.1% () | -46.8% () | 23.2% () | 38.4% () | -37.8% (*) | -8.3% () |
| *London* | Simple Ensemble | -12.4% () | -40.1% () | -64.3% (*) | 4.2% () | -14.6% () | -50.3% (**) |
| | Weighted Ensemble | 32.1% () | -9.7% () | -46.2% (*) | 2.0% () | -16.5% (*) | -51.4% (**) |
| *Madrid* | Simple Ensemble | -11.0% (*) | 49.5% (*) | -8.6% () | -44.6% (***) | 68.2% () | -21.0% () |
| | Weighted Ensemble | -6.8% () | 56.4% () | -4.3% () | -50.8% (***) | 49.3% () | -29.8% () |
| *New York* | Simple Ensemble | 47.4% (***) | -8.6% () | -40.9% (***) | 50.2% (***) | 28.8% () | -55.8% (***) |
| | Weighted Ensemble | 25.8% (**) | -22.1% () | -49.6% (***) | 45.9% (***) | 25.1% () | -57.1% (**) |
| *Rio de Janeiro* | Simple Ensemble | -29.8% () | -43.5% (****) | -63.9% (****) | -30.1% () | -44.5% (****) | -74.6% (****) |
| | Weighted Ensemble | -37.5% (**) | -49.7% (****) | -67.9% (****) | -45.6% (**) | -56.8% (****) | -80.2% (****) |
| *Santiago de Chile* | Simple Ensemble | -39.1% () | -28.8% (***) | -38.0% (****) | -32.1% (*) | -42.0% (****) | -51.5% (****) |
| | Weighted Ensemble | -48.0% () | -39.2% (****) | -47.1% (****) | -36.6% (*) | -45.8% (****) | -54.7% (****) |
| *Overall* | Simple Ensemble | -46.7% (*) | -31.4% (****) | -46.1% (****) | -32.4% (***) | -29.1% (****) | -45.5% (****) |
| | Weighted Ensemble | -46.6% (**) | -31.1% (****) | -45.9% (****) | -33.2% (****) | -29.9% (****) | -46.2% (****) |

Table 4: **Forecasting performance improvements of ensemble models with respect to individual epidemic-behavior models**. The table shows the percentage improvements in forecasting performance metrics (MAE and WIS) with respect to each individual model's performance in each location and over all of them. Negative percentages denote an improvement in forecasting performance, while positive values indicate a decline in performance. We consider a simple ensemble where each model is weighted equally, and a weighted ensemble where each model is weighted according to its past forecasting performance. Table also reports the statistical significance of the difference in performance of ensembles' and individual models' performance measured via Wilcoxon signed rank test. In particular: ****: $p_{value} \leq 10^{-4}$, ***: $10^{-4} < p_{value} \leq 10^{-3}$, **: $10^{-3} < p_{value} \leq 10^{-2}$, *: $10^{-2} < p_{value} \leq 0.05$, and otherwise blank if $p_{value} > 0.05$.

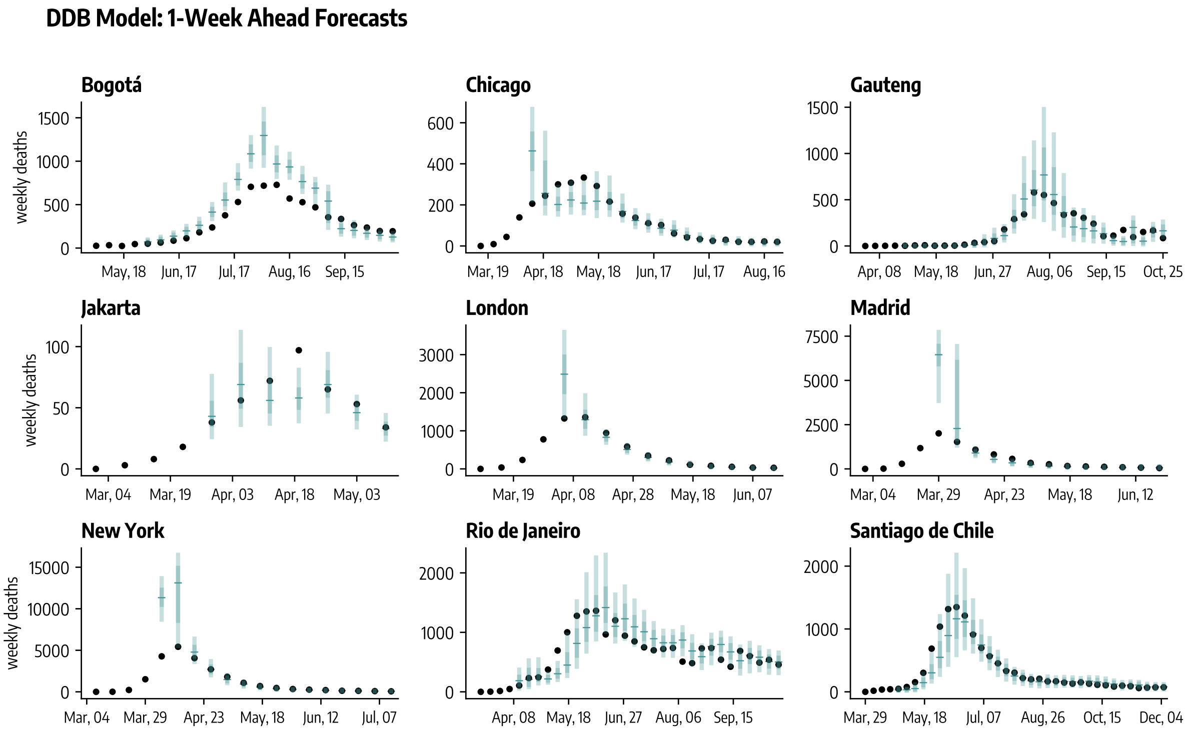

Figure 19: One-week ahead forecasts (median, 50%, and 90% predictive intervals obtained from 1,000 stochastic trajectories) of weekly deaths during the COVID-19 initial wave across nine geographies for the Data-Driven Behavioral model (DDB).

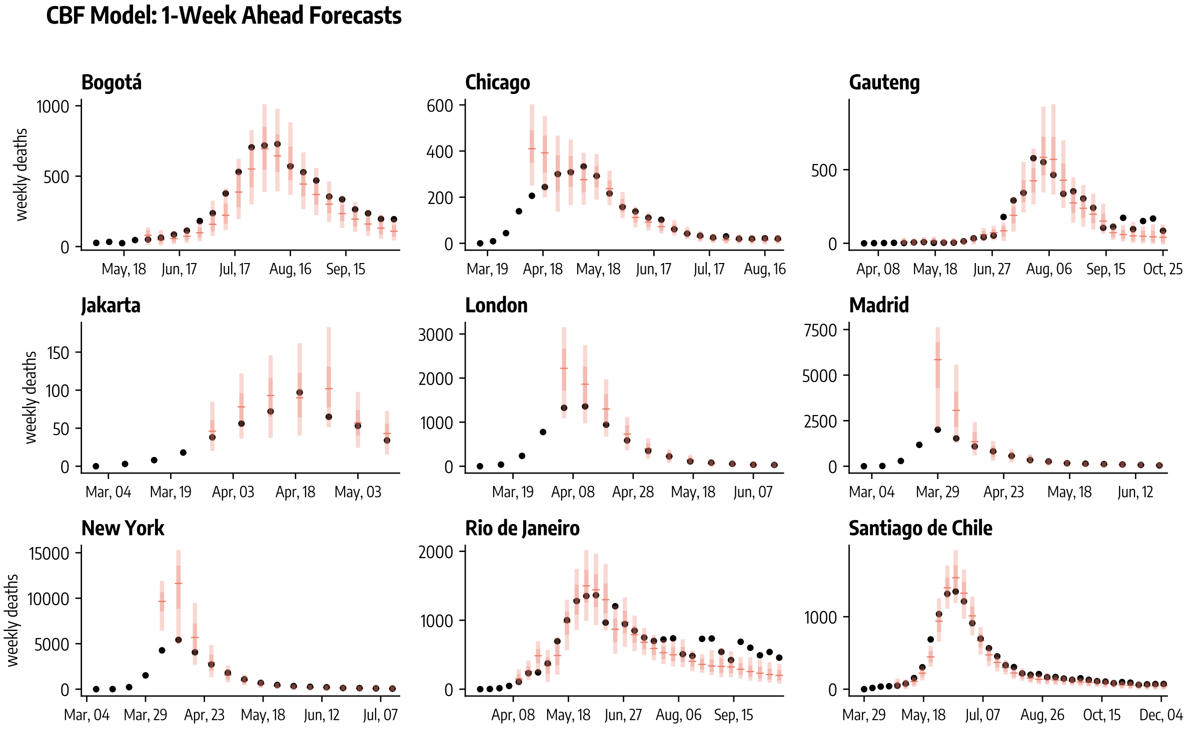

Figure 20: One-week ahead forecasts (median, 50%, and 90% predictive intervals obtained from 1,000 stochastic trajectories) of weekly deaths during the COVID-19 initial wave across nine geographies for the Compartmental Behavioral Feedback model (CBF).

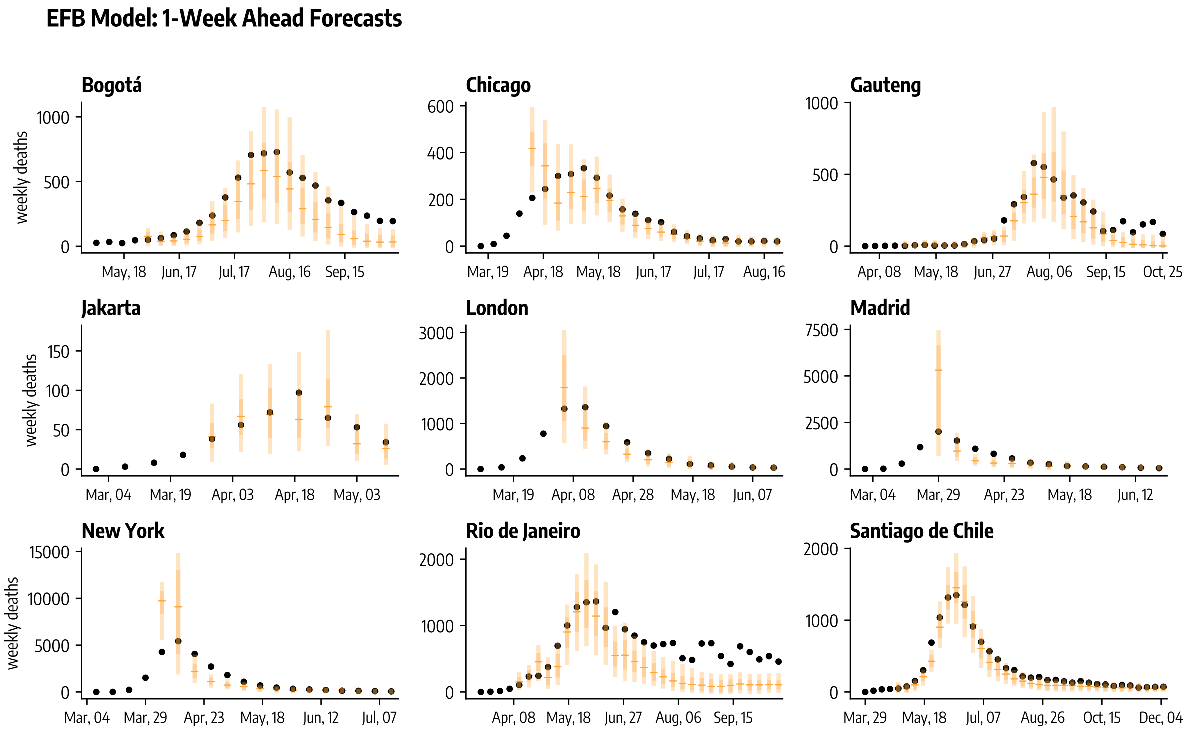

Figure 21: One-week ahead forecasts (median, 50%, and 90% predictive intervals obtained from $1,000$ stochastic trajectories) of weekly deaths during the COVID-19 initial wave across nine geographies for the Effective Force of Infection Behavioral Feedback model (EFB).

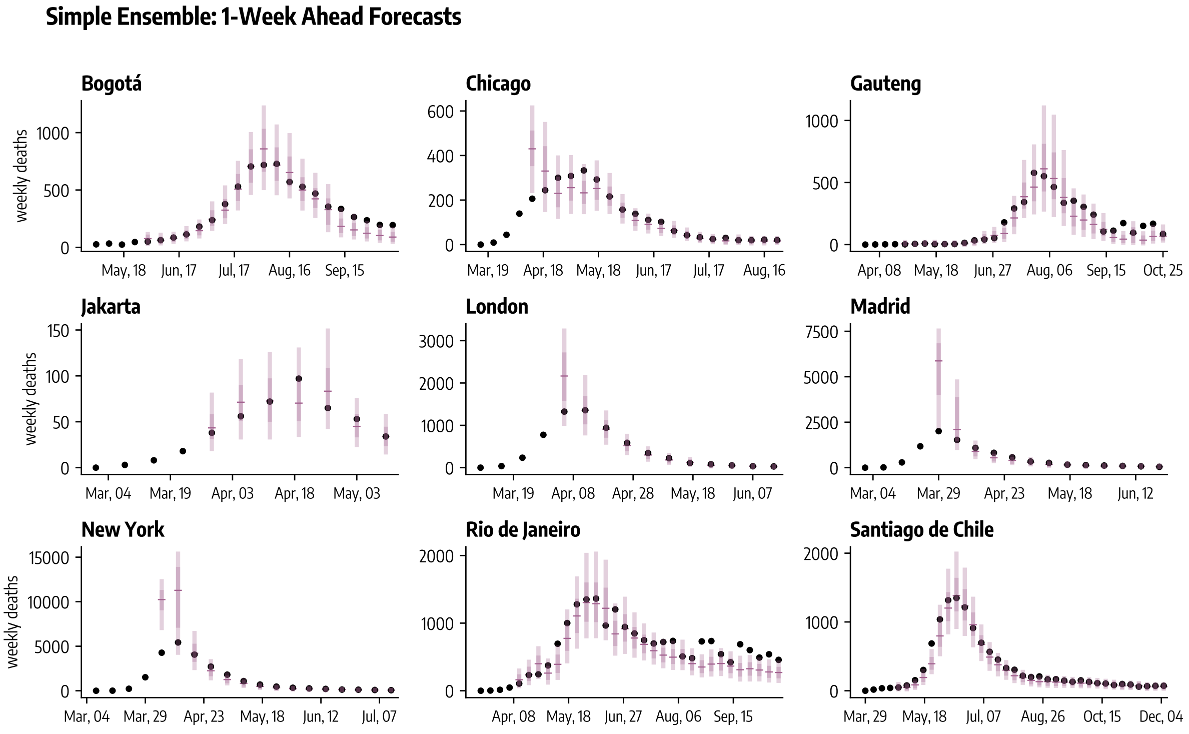

Figure 22: One-week ahead forecasts (median, 50%, and 90% predictive intervals obtained from $1,000$ stochastic trajectories) of weekly deaths during the COVID-19 initial wave across nine geographies for the simple ensemble.

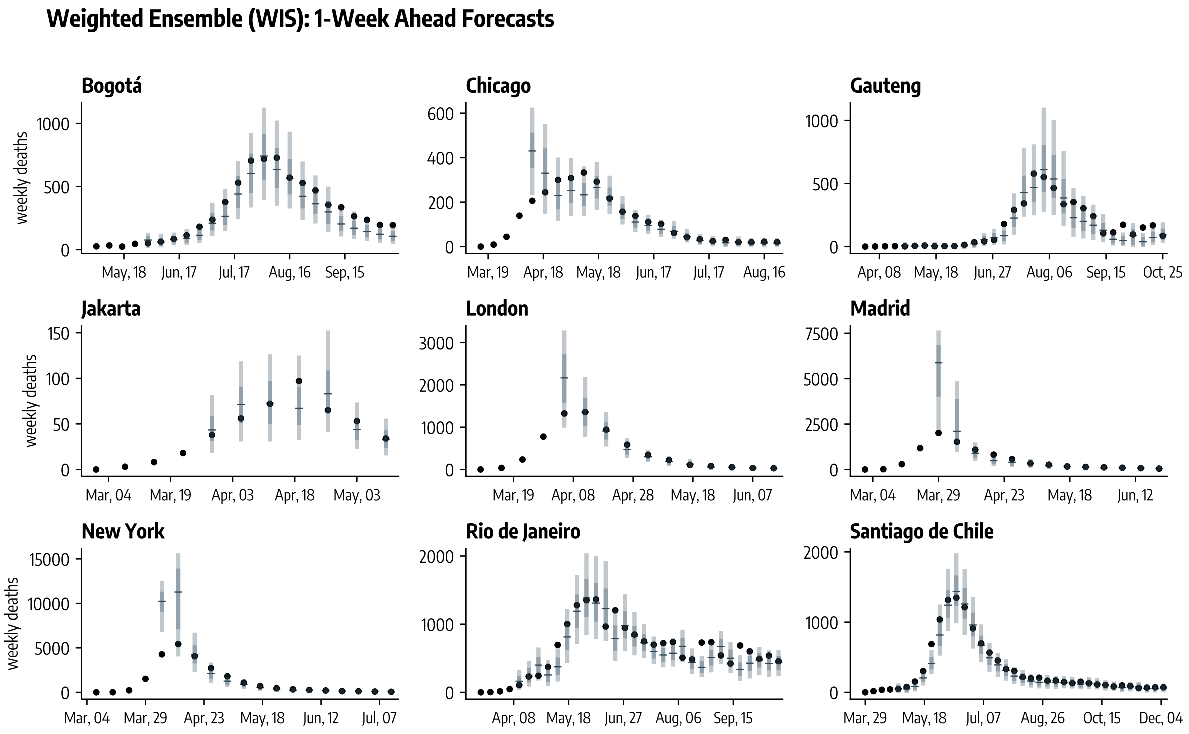

Figure 23: One-week ahead forecasts (median, 50%, and 90% predictive intervals obtained from 1,000 stochastic trajectories) of weekly deaths during the COVID-19 initial wave across nine geographies for the ensemble weighted according to past WIS performance.

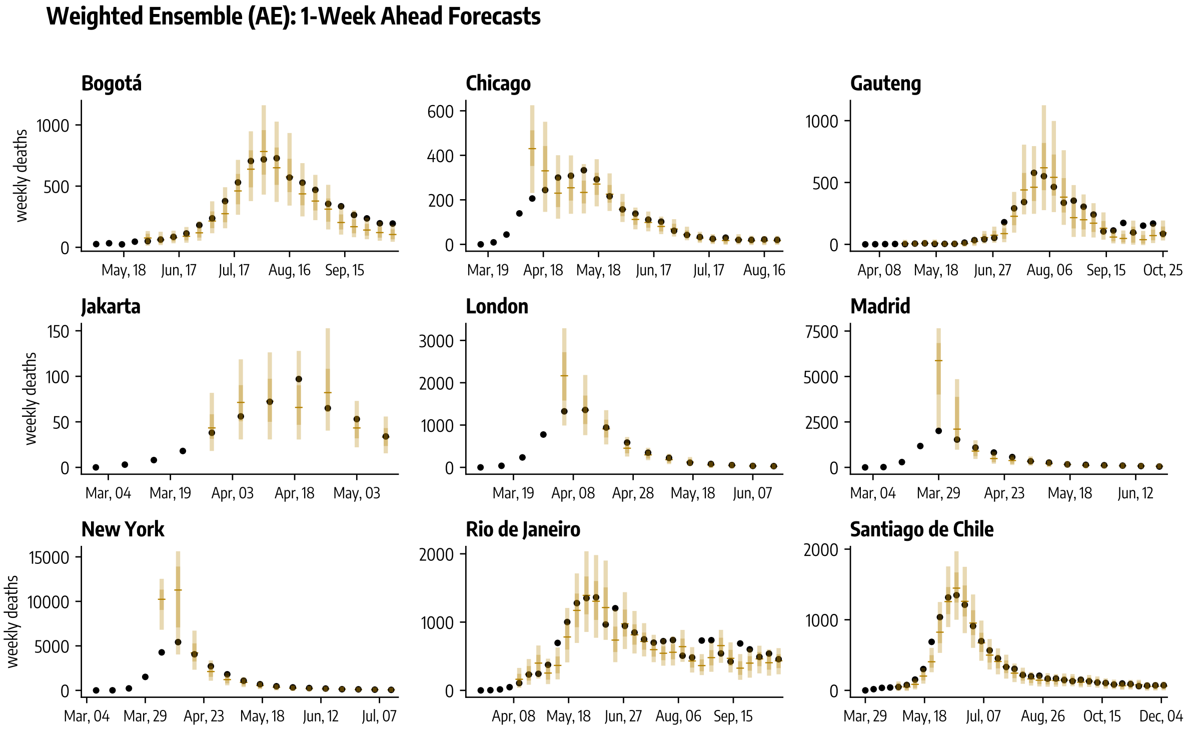

Figure 24: One-week ahead forecasts (median, 50%, and 90% predictive intervals obtained from 1,000 stochastic trajectories) of weekly deaths during the COVID-19 initial wave across nine geographies for the ensemble weighted according to past MAE performance.

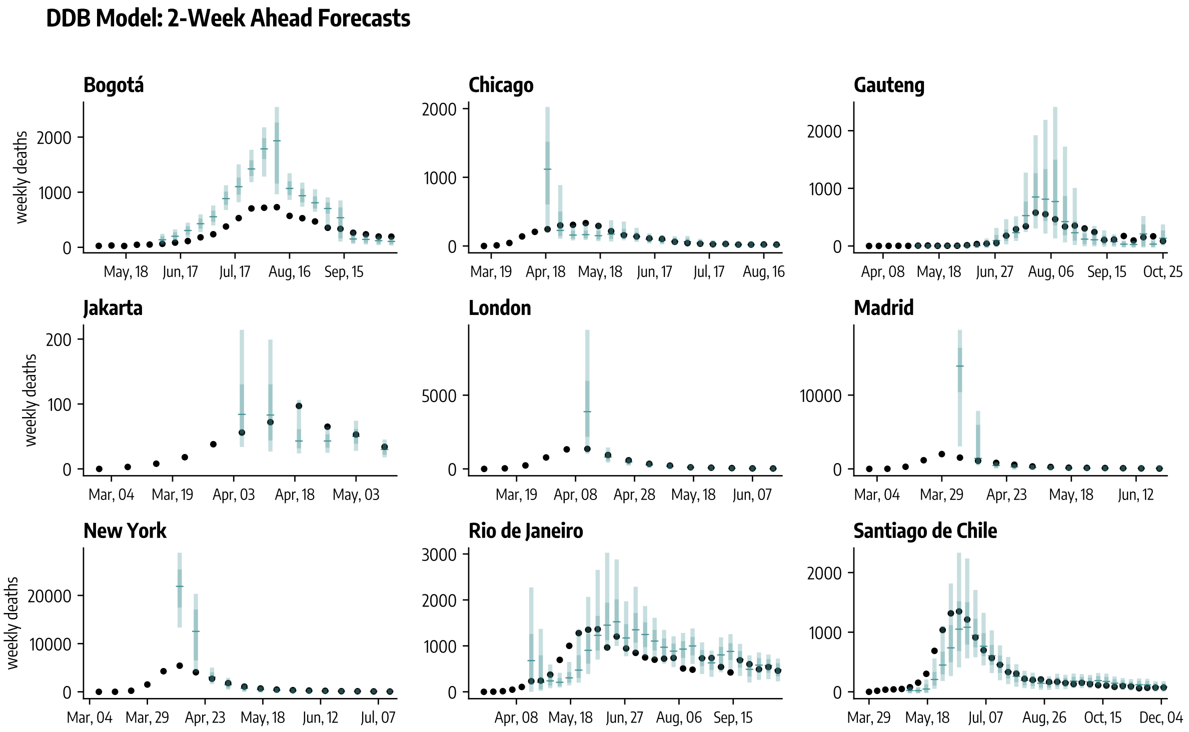

Figure 25: Two-week ahead forecasts (median, 50%, and 90% predictive intervals obtained from $1,000$ stochastic trajectories) of weekly deaths during the COVID-19 initial wave across nine geographies for the Data-Driven Behavioral model (DDB).

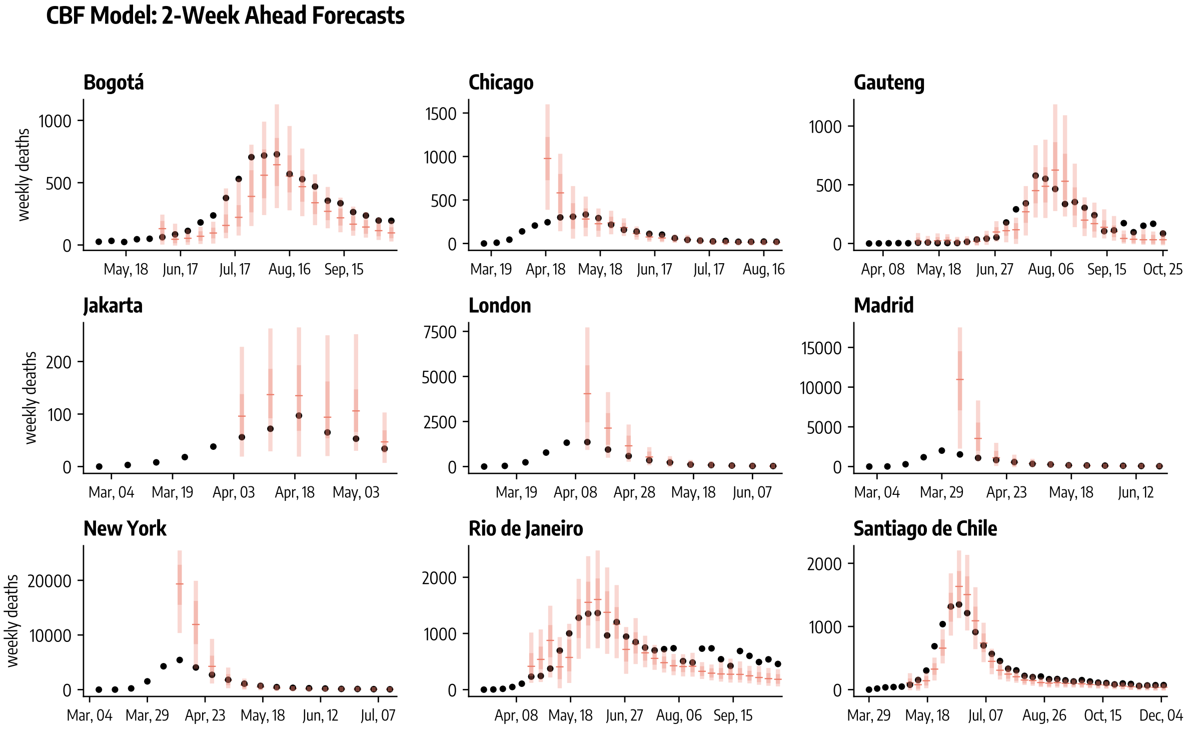

Figure 26: Two-week ahead forecasts (median, 50%, and 90% predictive intervals obtained from $1,000$ stochastic trajectories) of weekly deaths during the COVID-19 initial wave across nine geographies for the Compartmental Behavioral Feedback model (CBF).

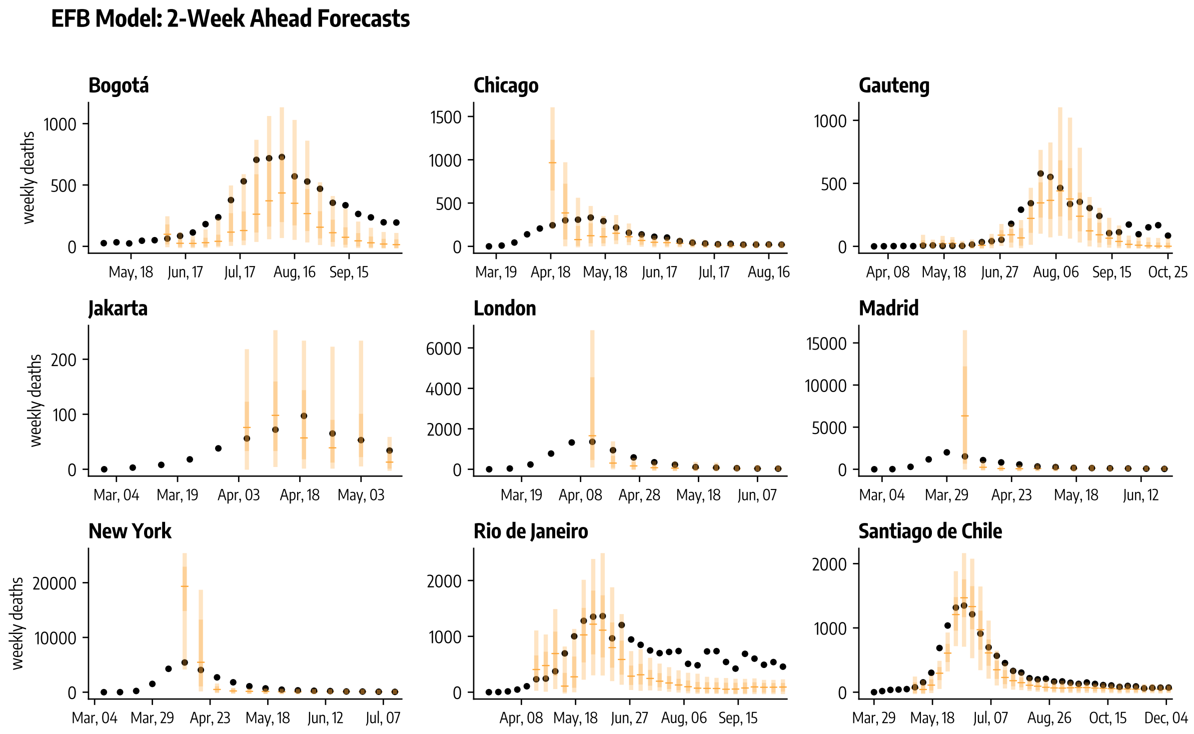

Figure 27: Two-week ahead forecasts (median, 50%, and 90% predictive intervals obtained from 1,000 stochastic trajectories) of weekly deaths during the COVID-19 initial wave across nine geographies for the Effective Force of Infection Behavioral Feedback model (EFB).

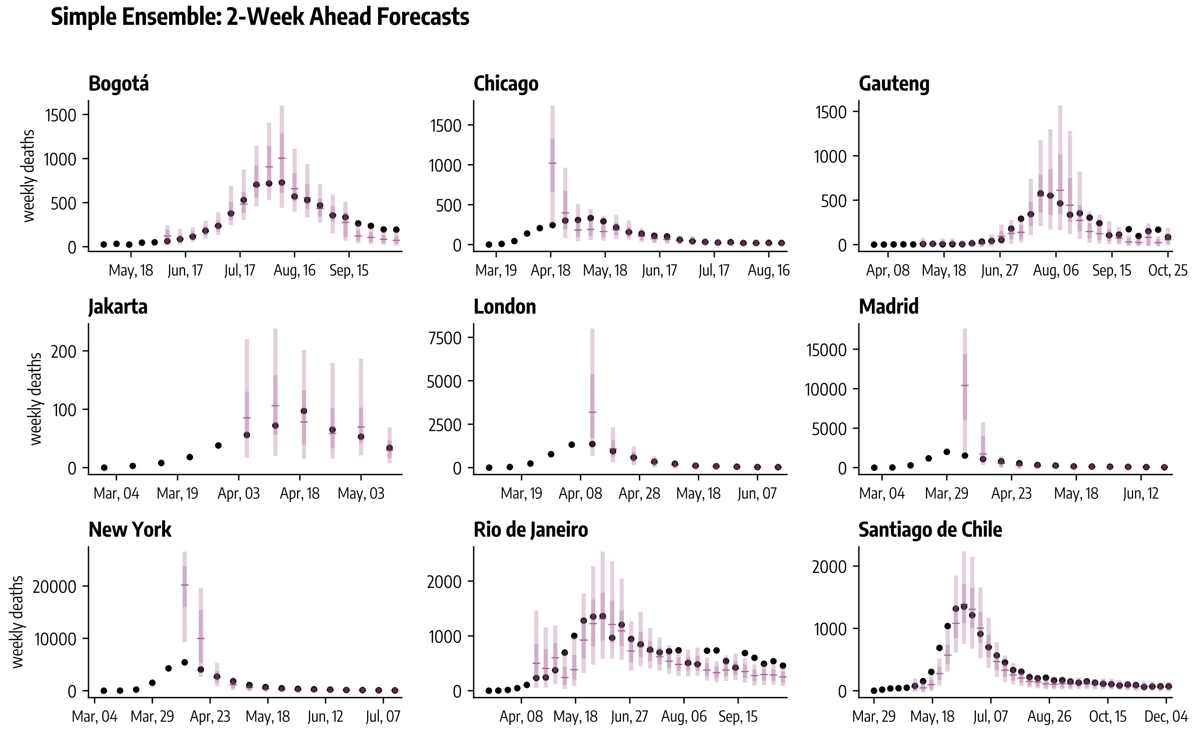

Figure 28: Two-week ahead forecasts (median, 50%, and 90% predictive intervals obtained from 1,000 stochastic trajectories) of weekly deaths during the COVID-19 initial wave across nine geographies for the simple ensemble.

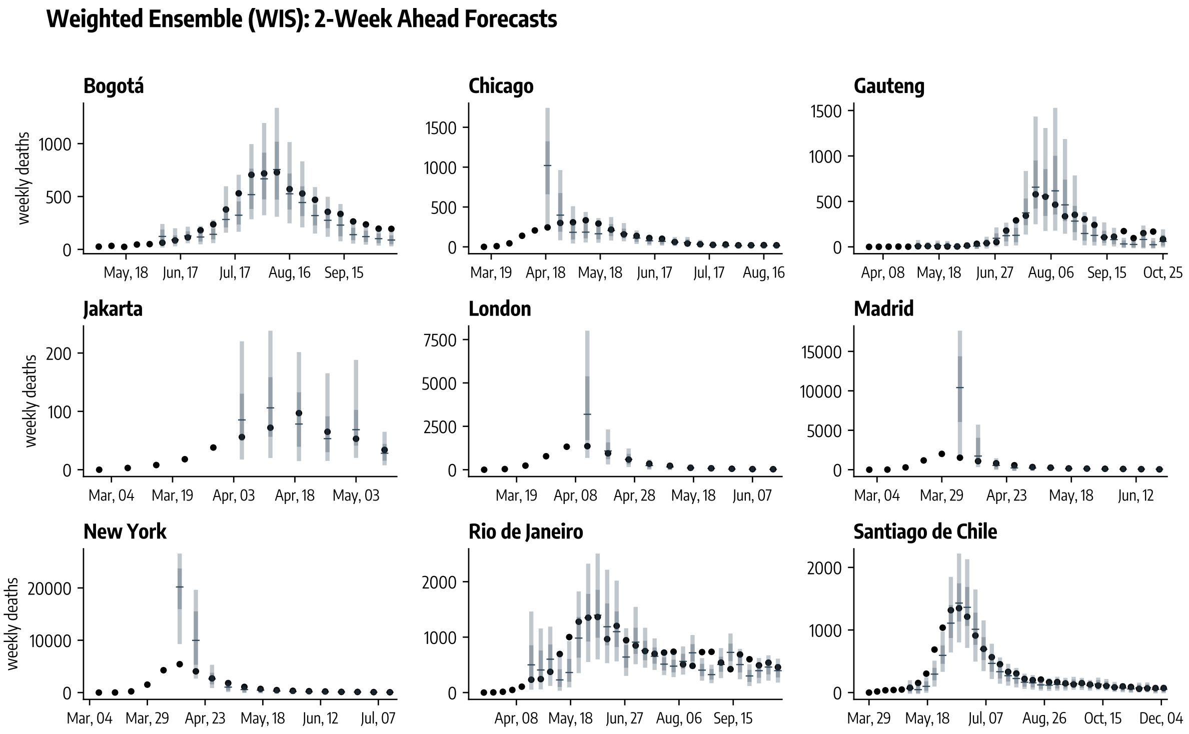

Figure 29: Two-week ahead forecasts (median, 50%, and 90% predictive intervals obtained from 1,000 stochastic trajectories) of weekly deaths during the COVID-19 initial wave across nine geographies for the ensemble weighted according to past WIS performance.

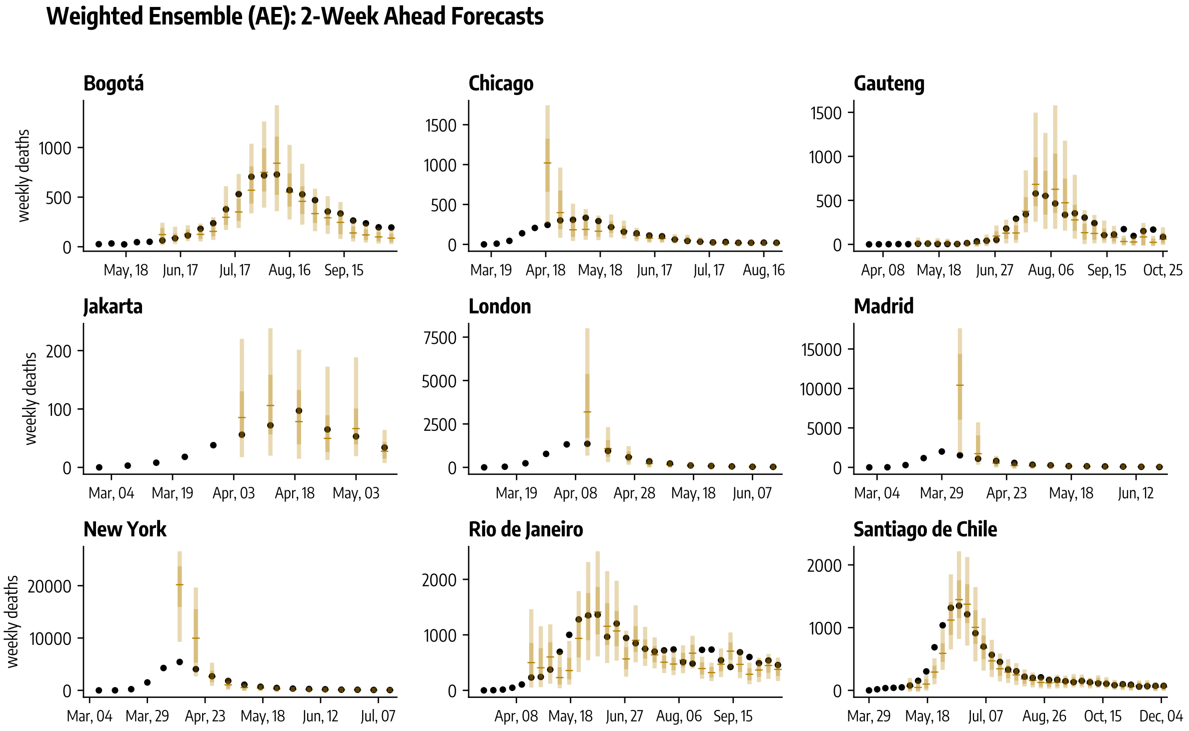

Figure 30: Two-week ahead forecasts (median, 50%, and 90% predictive intervals obtained from 1,000 stochastic trajectories) of weekly deaths during the COVID-19 initial wave across nine geographies for the ensemble weighted according to past MAE performance.

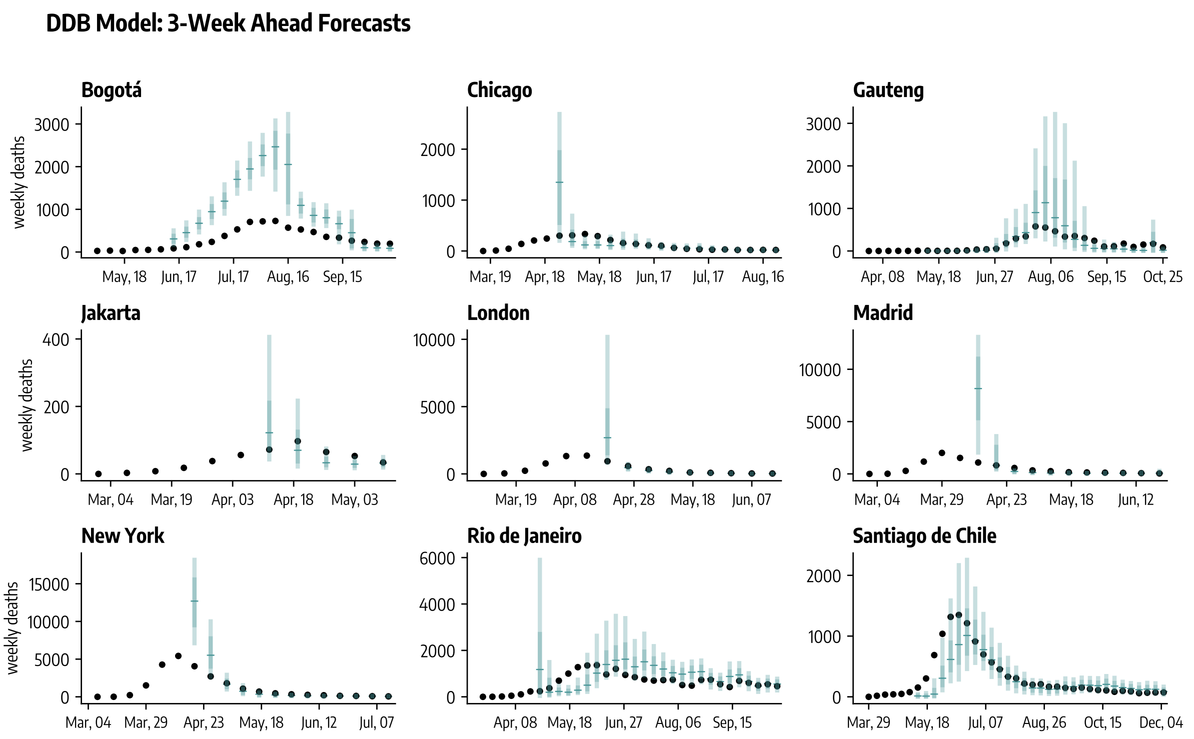

Figure 31: Three-week ahead forecasts (median, 50%, and 90% predictive intervals obtained from 1,000 stochastic trajectories) of weekly deaths during the COVID-19 initial wave across nine geographies for the Data-Driven Behavioral model (DDB).

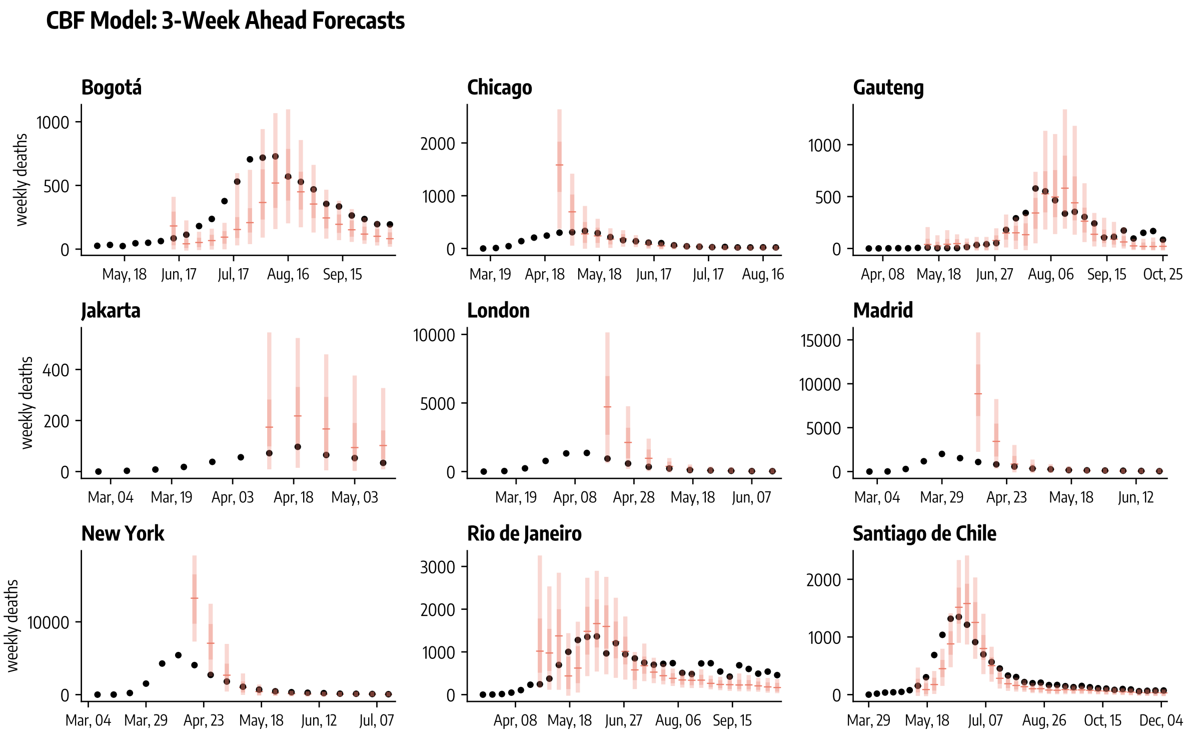

Figure 32: Three-week ahead forecasts (median, 50%, and 90% predictive intervals obtained from 1,000 stochastic trajectories) of weekly deaths during the COVID-19 initial wave across nine geographies for the Compartmental Behavioral Feedback model (CBF).

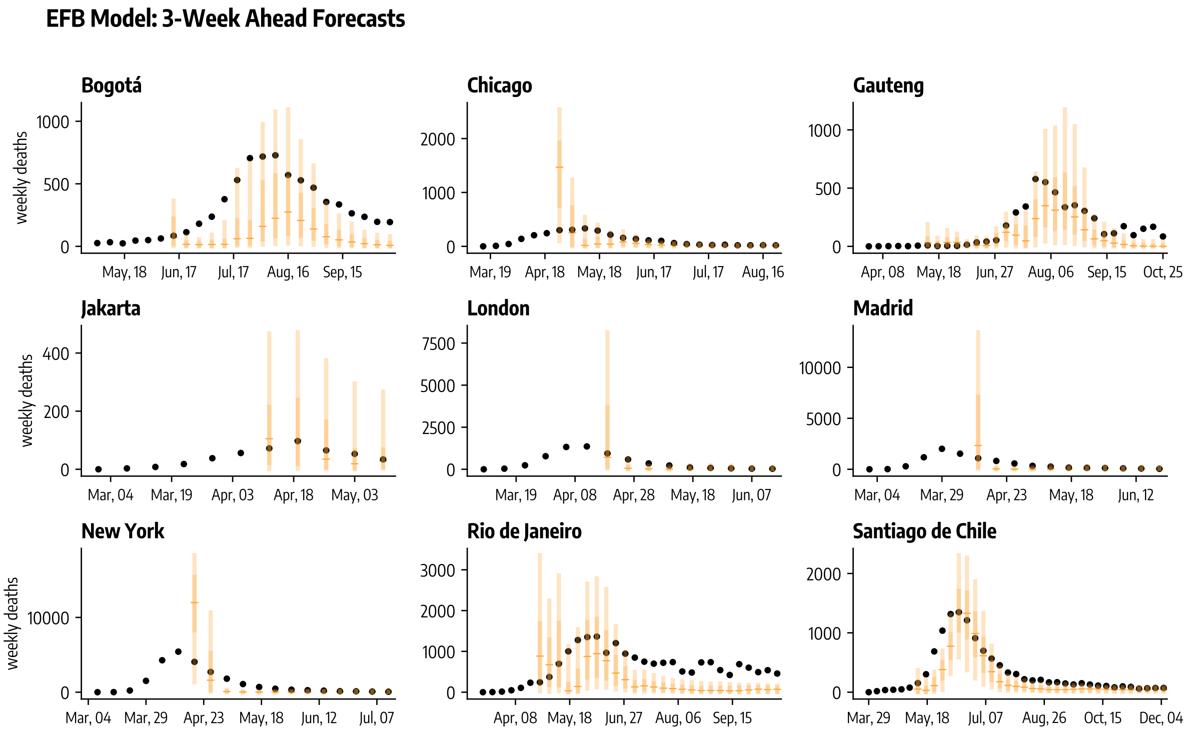

Figure 33: Three-week ahead forecasts (median, 50%, and 90% predictive intervals obtained from 1,000 stochastic trajectories) of weekly deaths during the COVID-19 initial wave across nine geographies for the Effective Force of Infection Behavioral Feedback model (EFB).

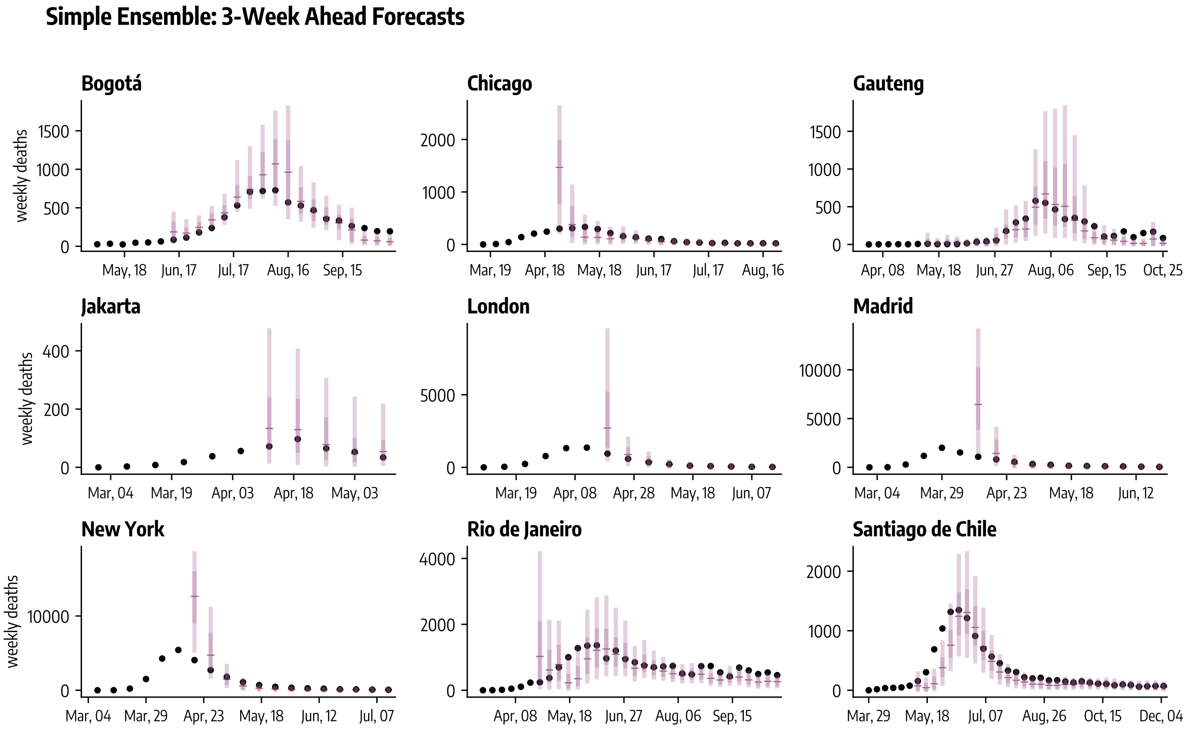

Figure 34: Three-week ahead forecasts (median, 50%, and 90% predictive intervals obtained from 1,000 stochastic trajectories) of weekly deaths during the COVID-19 initial wave across nine geographies for the simple ensemble.

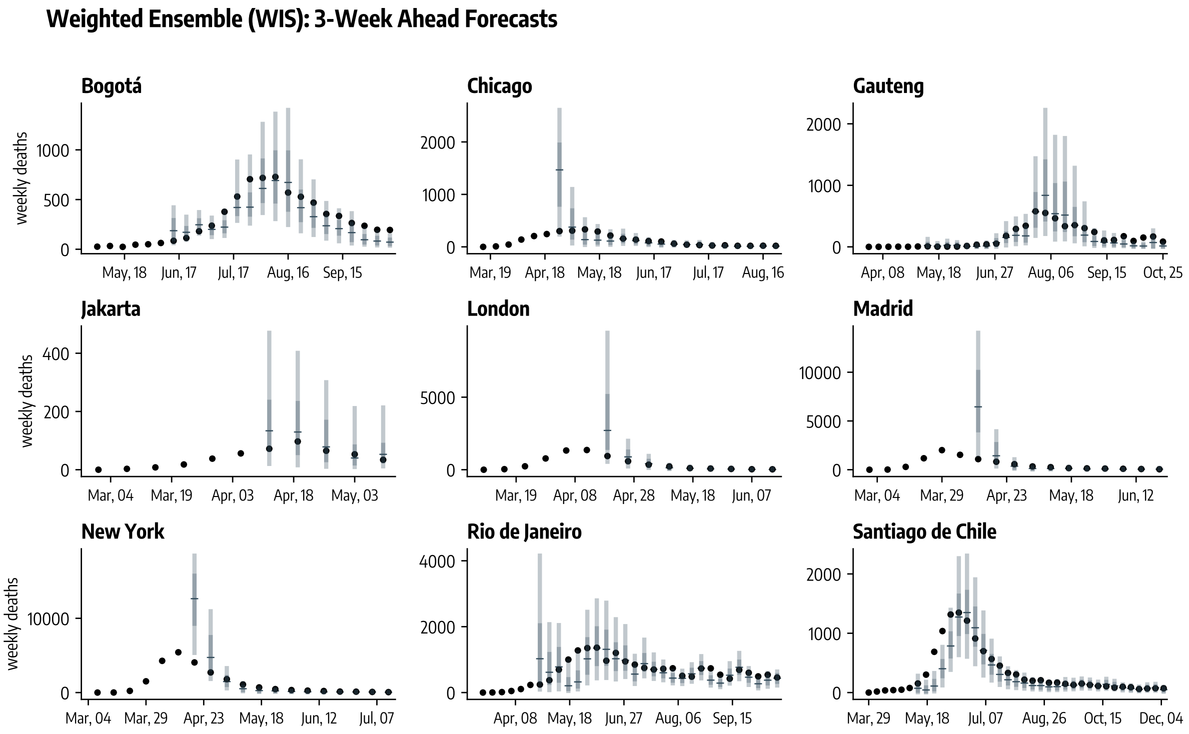

Figure 35: Three-week ahead forecasts (median, 50%, and 90% predictive intervals obtained from 1,000 stochastic trajectories) of weekly deaths during the COVID-19 initial wave across nine geographies for the ensemble weighted according to past WIS performance.

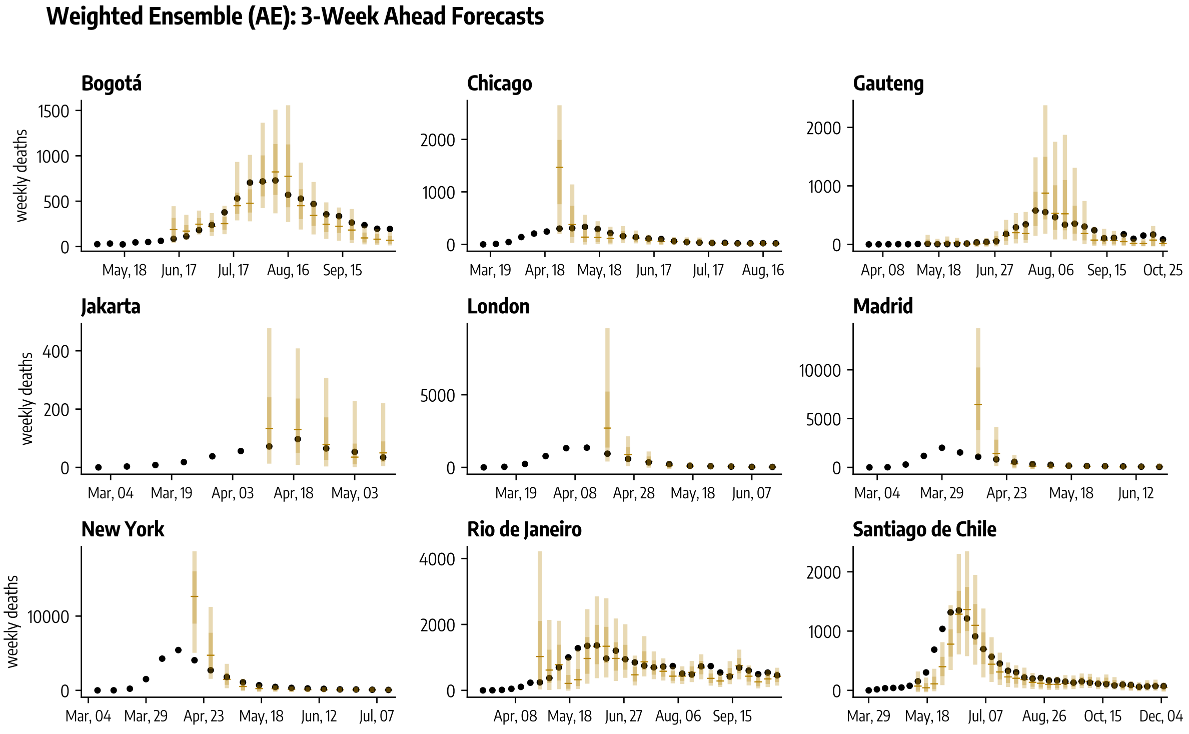

Figure 36: Three-week ahead forecasts (median, 50%, and 90% predictive intervals obtained from 1,000 stochastic trajectories) of weekly deaths during the COVID-19 initial wave across nine geographies for the ensemble weighted according to past MAE performance.

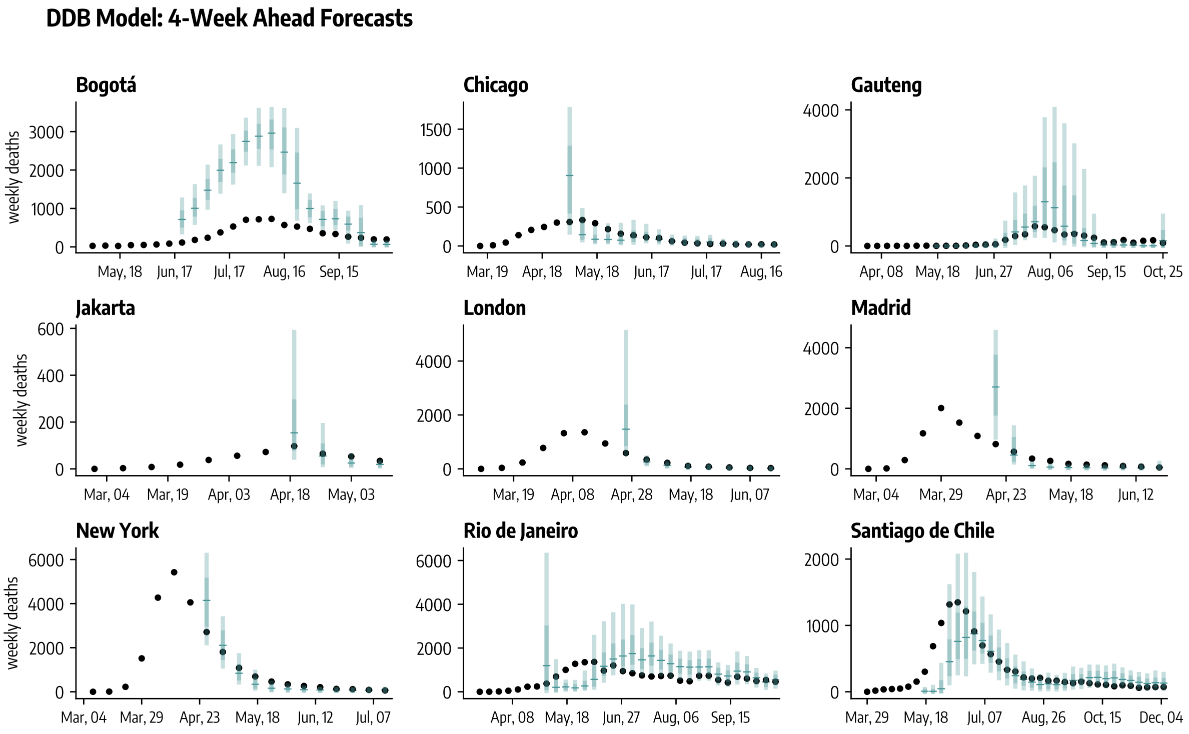

Figure 37: Four-week ahead forecasts (median, 50%, and 90% predictive intervals obtained from 1,000 stochastic trajectories) of weekly deaths during the COVID-19 initial wave across nine geographies for the Data-Driven Behavioral model (DDB).

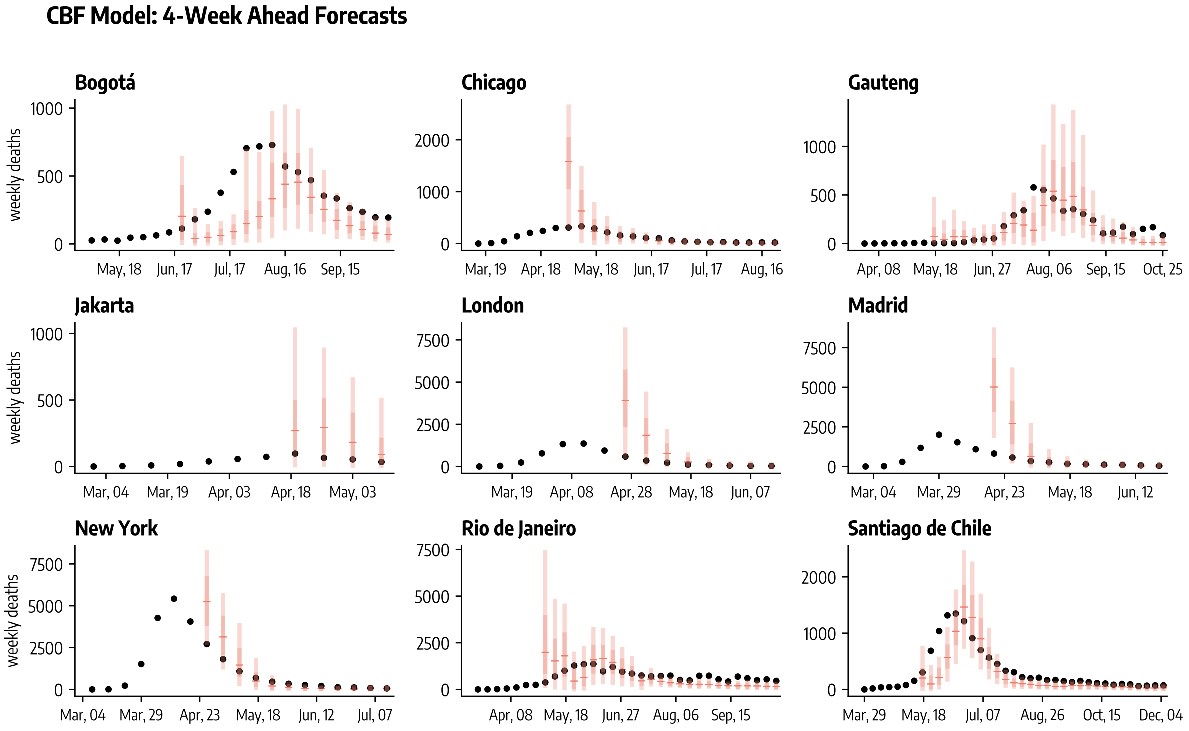

Figure 38: Four-week ahead forecasts (median, 50%, and 90% predictive intervals obtained from 1,000 stochastic trajectories) of weekly deaths during the COVID-19 initial wave across nine geographies for the Compartmental Behavioral Feedback model (CBF).

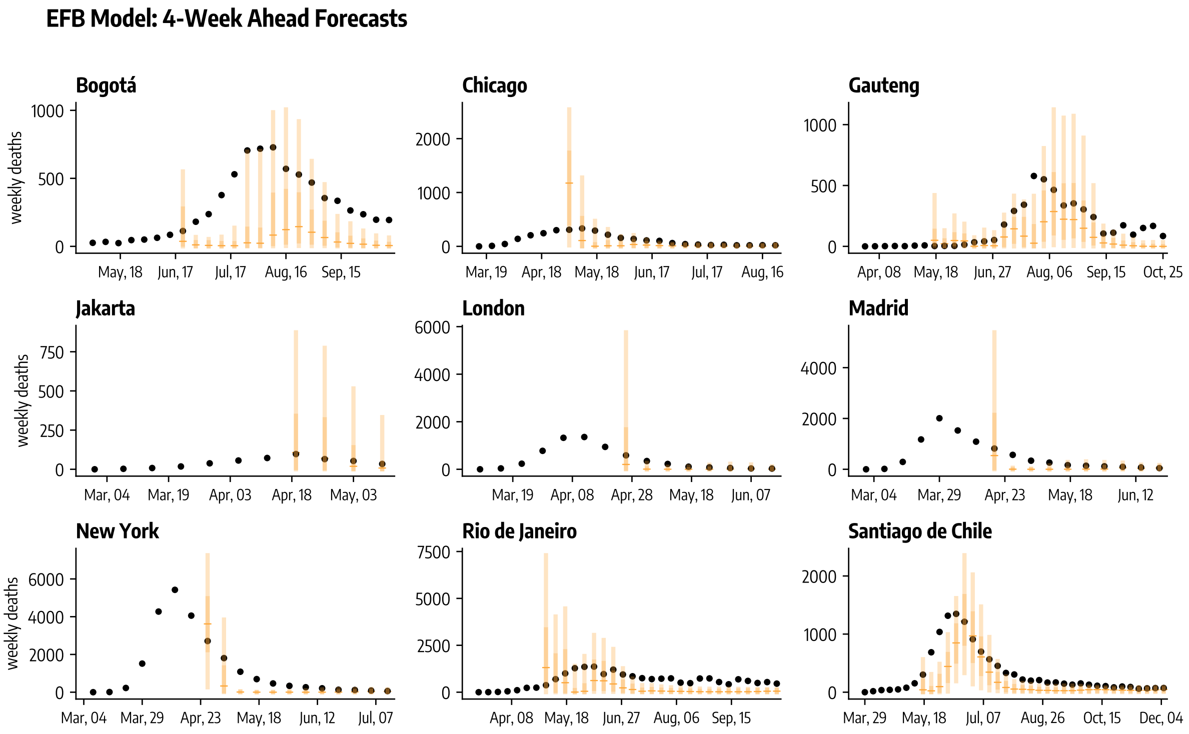

Figure 39: Four-week ahead forecasts (median, 50%, and 90% predictive intervals obtained from 1,000 stochastic trajectories) of weekly deaths during the COVID-19 initial wave across nine geographies for the Effective Force of Infection Behavioral Feedback model (EFB).

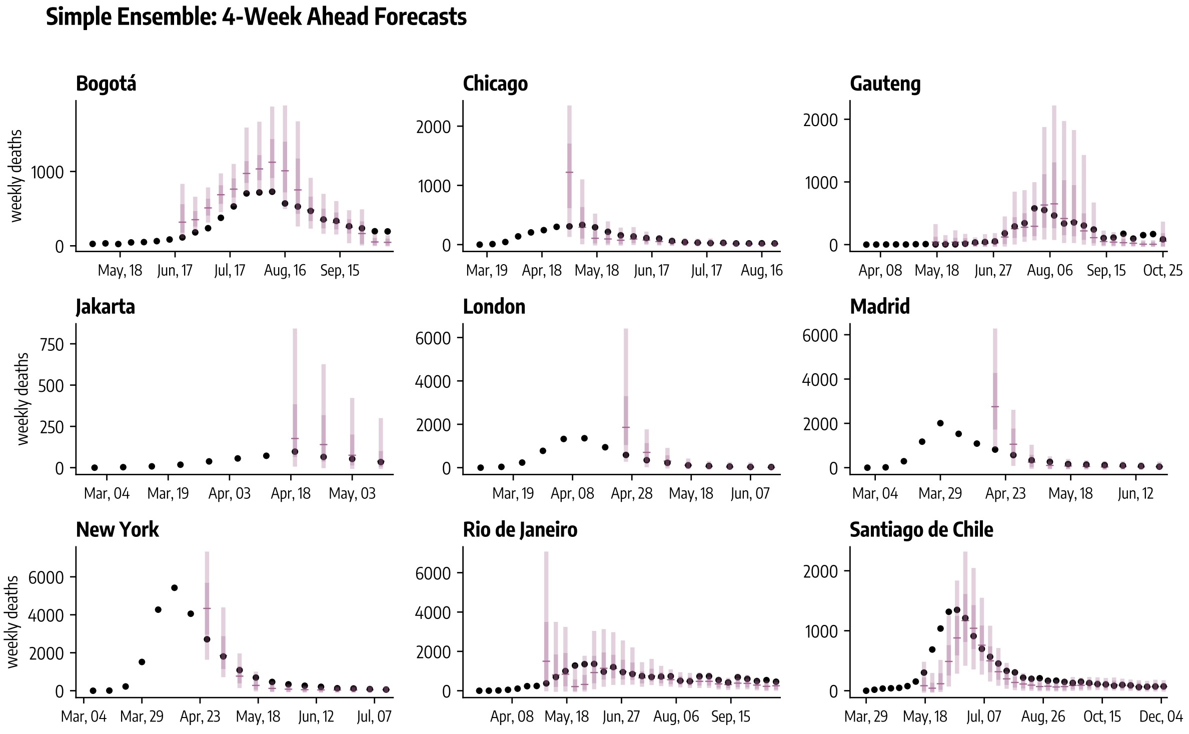

Figure 40: Four-week ahead forecasts (median, 50%, and 90% predictive intervals obtained from 1,000 stochastic trajectories) of weekly deaths during the COVID-19 initial wave across nine geographies for the simple ensemble.

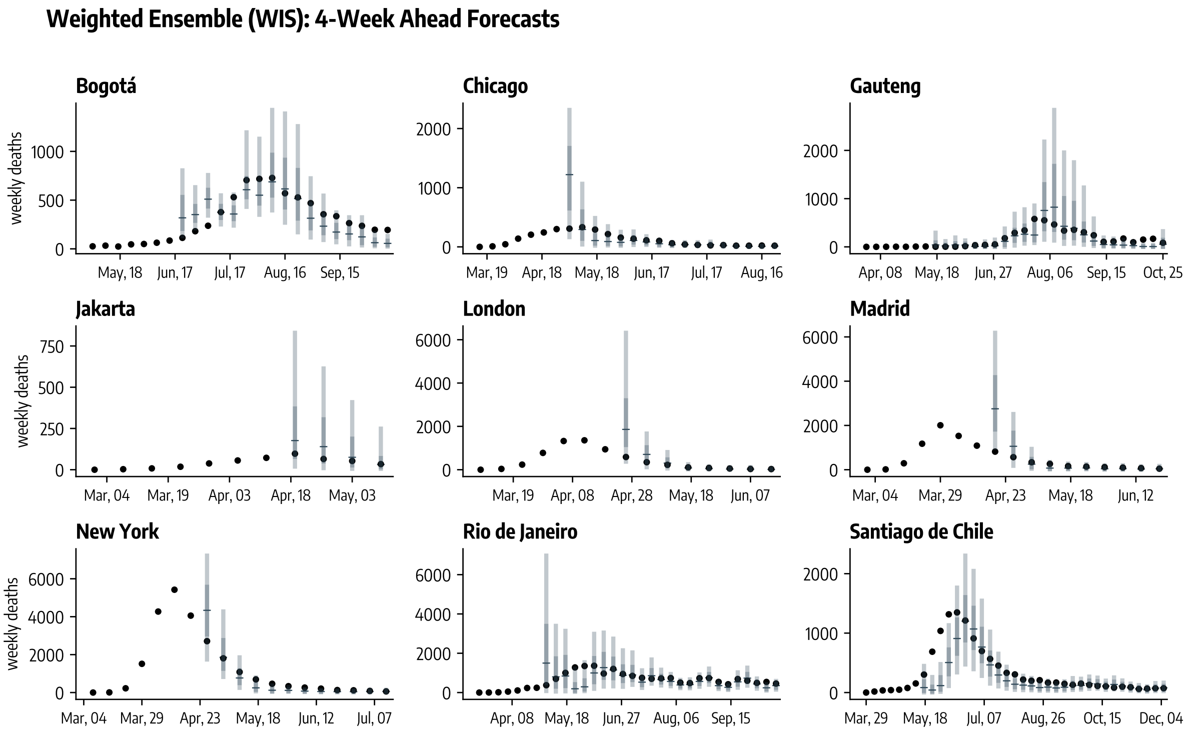

Figure 41: Four-week ahead forecasts (median, 50%, and 90% predictive intervals obtained from 1,000 stochastic trajectories) of weekly deaths during the COVID-19 initial wave across nine geographies for the ensemble weighted according to past WIS performance.

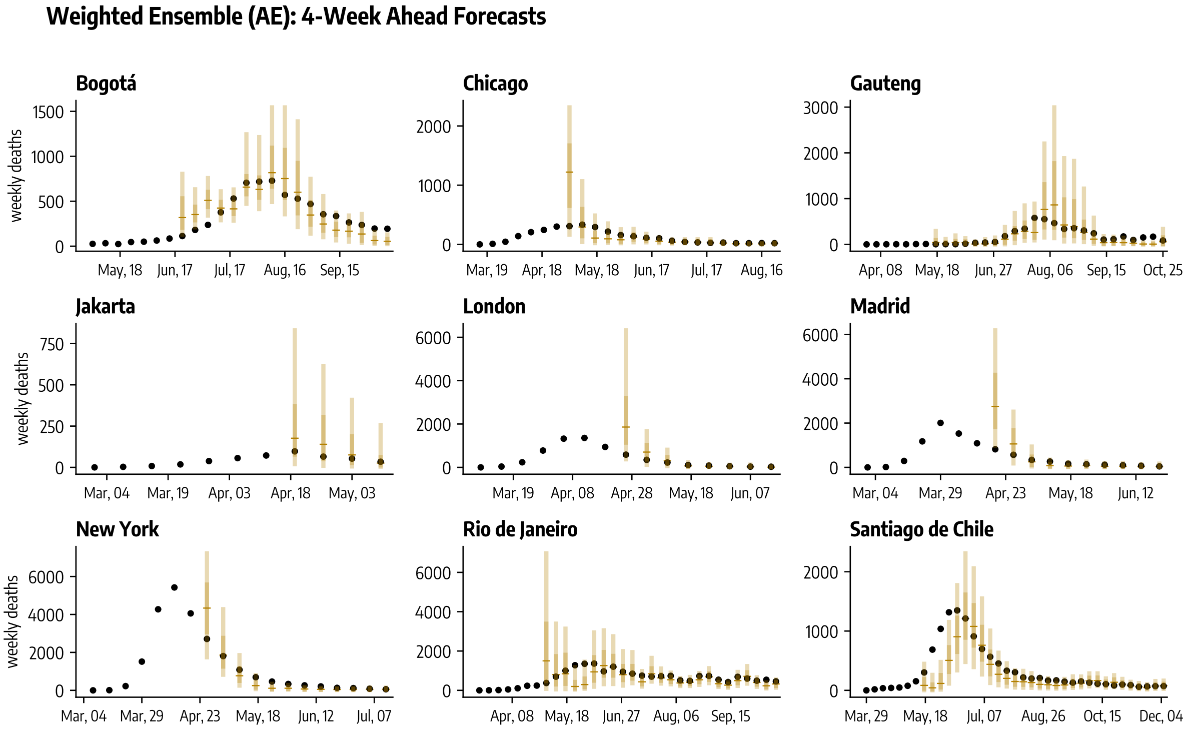

Figure 42: Four-week ahead forecasts (median, 50%, and 90% predictive intervals obtained from 1,000 stochastic trajectories) of weekly deaths during the COVID-19 initial wave across nine geographies for the ensemble weighted according to past MAE performance.
